# Quantitative evidence for relational care approaches to assessing and managing self-harm and suicide risk in inpatient mental health and emergency department settings: a scoping review

**DOI:** 10.1101/2024.08.08.24311667

**Authors:** Jessica L. Griffiths, Una Foye, Ruth Stuart, Ruby Jarvis, Beverley Chipp, Raza Griffiths, Tamar Jeynes, Lizzie Mitchell, Jennie Parker, Rachel Rowan Olive, Kieran Quirke, John Baker, Geoff Brennan, Gary Lamph, Mick McKeown, Brynmor Lloyd-Evans, Kylee Trevillion, Alan Simpson

## Abstract

There is an over-reliance on structured risk assessments and restrictive practices for managing self-harm and suicidality in inpatient mental health and emergency department (ED) settings, despite a lack of supporting evidence. Alternative ‘relational care’ approaches prioritising interpersonal relationships are needed. We present a definition of ‘relational care’, co-produced with academic and lived experience researchers and clinicians, and conducted a scoping review, following PRISMA guidelines. We aimed to examine quantitative evidence for the impact of ‘relational care’ in non-forensic inpatient mental health and ED settings on self-harm and suicide. We identified 29 relevant reviews, covering 62 relational care approaches, reported in 87 primary papers. Evidence suggests some individual-, group-, ward- and organisation-level relational care approaches can reduce self-harm and suicide in inpatient mental health and ED settings, although there is a lack of high-quality research overall. Further co-produced research is needed to clarify the meaning of ‘relational care’, its core components, and develop a clear framework for its application and evaluation. Further high-quality research is needed evaluating its effectiveness, how it is experienced by patients, carers, and staff, and exploring what works best for whom, under what circumstances, and why.

## Introduction

Suicidality and self-harm remain key reasons for inpatient admissions in both acute and mental health hospitals. Therefore, a key purpose of inpatient mental health services and emergency departments (EDs) is to provide a safe environment for people presenting with, and at-risk of, self-harm and/or suicide (1,2).

During the years 2011-21, 28% of people in the UK who died by suicide were patients in acute care settings (inpatients, under crisis resolution/home treatment teams, or recently discharged from in-patient care) (3). Rates of inpatient suicide per 10,000 admissions fell by 33% over this 11-year period. There were on average 31 deaths by suicide on UK wards annually during this period (3).

In England alone, there are approximately 220,000 self-harm presentations to EDs annually (4,5) and such individuals have a 49 times greater relative risk of suicide than that of the general population (6). Self-harm is the most frequently reported incident in mental health services and rates of self-harm have increased over time (7). Self-harm rates on inpatient mental health wards vary, with studies reporting between 4% and 70% of patients harming themselves during admission to inpatient services (8). Self-harm has been found to most often be a private act, which takes place in bedrooms, bathrooms and toilets, and during the evening hours (9).

Given the prevalence of self-harm and suicidality in inpatient mental health and ED settings, these patient groups have been identified as a priority within national suicide prevention strategies (10). Efforts to enhance their safety have been made, including the implementation of varied interventions, policies and guidelines (1,3). This includes, more recently, the use of surveillance technologies, such as Vision-Based Patient Monitoring and Management, Body Worn Cameras, and closed-circuit television (CCTV). However, there is a lack of evidence for their effectiveness in improving patient safety, and ethical concerns about their potential to negatively impact patients’ human rights, privacy, dignity and recovery (11). Inpatient and ED settings remain challenging environments in which to deliver appropriate and effective care (12–15).

Both inpatient mental health and ED settings are often fast-paced and over-stimulating environments, with high levels of distress, limited therapeutic options, lack of patient choice, inadequate involvement of families and carers, negative staff attitudes towards people who self-harm, and poor continuity of care. The consequences of this include high rates of conflict, coercion and restrictive practices (16–18). Specific challenges faced by emergency departments also include their single-visit nature, heavy footfall, long waiting times, and brief durations of each human encounter (19). In both settings, these challenges are compounded by systemic issues including rising demands on services, increasing acuity of patients’ presentations, temporary and under-staffing, and inadequate funding and resourcing (12,13). A recent independent rapid review on mental health inpatient care identified key safety issues facing inpatient settings (20).

Those who present to EDs in emotional distress and requiring interventions and treatment for self-harm injuries may be directly or indirectly excluded by services, owing to prioritisation of others with physical health conditions, public discourse about system strain, and efforts to divert mental health cases elsewhere. Although they might be seen initially within an hour, their stay in the ED, or separate decision unit, can be as long as 48-72 hours as they wait for an outcome such as hospital admission. Most ED settings have mental health liaison services attached but these are often underutilised (21,22). Frequent attendance at ED settings is likely driven by limitations within other services in the healthcare system, rejection by other services, lack of clarity of service provisions available, and in some cases convenience. For example, it is often the only local or out-of-hours service accessible to people (23).

These challenges contribute to an over-reliance in inpatient mental health and ED settings on using structured risk assessments and risk stratification to assess self-harm and suicide risk, and the use of restrictive practices, such as physical restraint, seclusion, rapid tranquilisation, and special observations to manage concerns over risk and safety (6,22–24). This is despite research consistently demonstrating the ineffectiveness of risk assessment checklists for predicting self-harm and suicide risk and the potential for restrictive practices to undermine therapeutic relationships and cause physical and psychological harm to patients and staff (9,24–26). There is, therefore, a growing need for alternative approaches in the assessment and management of self-harm and suicide risk in inpatient mental health and ED settings.

Positive relationships between staff and the people they support are fundamental to a person-centred care environment and have been identified as key to a positive culture of care in new guidance for mental health inpatient services (27). Positive therapeutic relationships between patients and clinicians are central to high-quality mental health care, and strong, consistent predictors of positive outcomes across a range of intervention types and settings (28–30).

Therapeutic relationships can underpin interventions and practices and can also be “therapy in and of itself” (29). Research indicates that patients value genuine listening, validation, warmth and curiosity within therapeutic relationships with clinicians, and that this can help build trust and facilitate disclosures about risk (23,31–34). There has, therefore, been an increasing interest in approaches to risk assessment and management which prioritise therapeutic interpersonal relationships – i.e. ‘relational’ approaches to care.

### What is ‘relational care’?

There is no widely agreed definition of ‘relational care’. It has been described across a diverse range of sectors, including health, education, criminal justice and social work (35). It also forms an integral part of practices and professional identities within professions such as nursing, psychology, social work, criminal justice and medicine, as well as in peer support work (36). Alongside the lack of an agreed consistent definition is also the challenge that across the sectors there is not a consistent descriptor or term used. Instead, there are many variations that all ultimately describe similar concepts. Furthermore, it is not a new concept – elements of it have been described for centuries. The conceptualisation of ‘relational care’ has therefore varied across time and contexts, and despite this term becoming increasingly used and topical, defining it remains a complex task, especially in the context of mental health care, where many types of relationships are involved (e.g., patient-patient, patient-staff, staff-staff and the overall ward or ED milieu).

For this project, a necessary working definition of ‘relational care’ within inpatient mental health and ED settings was coproduced by our working group, comprising academic and lived experience researchers and clinicians, as follows: “*Relational care can be practised at individual, group, organisational or systemic levels. It prioritises interpersonal relationships grounded in values such as respect, trust, humility, compassion, and shared humanity, and involves personalised and holistic care, addressing power imbalances, and promoting effective collaboration between staff, patients and their social networks.”^1^*

An organisational commitment to relational care, and reducing restrictive practices, is essential to provide the basis for developing and sustaining therapeutic relationships between staff and patients (27), from first contact (such as with paramedics and ambulance staff), in EDs and on inpatient wards.

It is important to acknowledge the tensions between practising relational care in a setting that most patients experience as initially coercive and restrictive. In inpatient mental health services, there are pronounced power imbalances between staff and patients, and patients have limited choice and agency. Democratisation of care in these services may, therefore, be considered aspirational at present. In striving for relational care, it is important to both acknowledge and take active steps towards addressing these power imbalances (26).

The environments in which relational interactions take place are important to consider as they need to be conducive to impact positively upon relational care experience, and we can conceive of configurations of space and place that are systemically more likely to support relational practice (35). For example, ward designs that maximise shared spaces, rather than demarcate space into designated staff and patient areas, or ward and ED layouts featuring outside areas and few confined spaces (45–47).

Though not their only defining characteristic, ‘relational care’ is a fundamental part of other approaches to care, such as trauma-informed, person-centred, or recovery-focused care. It is also integral to psychological therapies, encompassing the soft skills needed to foster the therapeutic relationships between staff and patients that are fundamental to effective therapy. In this paper, psychological therapies are therefore included as relational care.

### Review objective

This scoping review aimed to answer the following research question: What is the quantitative evidence for the impact of ‘relational care’ in non-forensic inpatient mental health and ED settings on self-harm and suicide-related outcomes?

A scoping review methodology was deemed most appropriate due to the lack of a consistent definition of ‘relational care’, its conceptual complexity, and the limited research on this emerging topic. This approach allows us to broadly and systematically map relevant existing literature, and to identify gaps, key issues and themes.

## Materials and methods

This scoping review was conducted in accordance with the Preferred Reporting Items for Systematic Reviews and Meta-Analyses Extension for Scoping Reviews (PRISMA-ScR) (48). The PRISMA-ScR checklist can be seen in Appendix C. The review was conducted by the National Institute for Health and Care Research (NIHR) Policy Research Unit in Mental Health (MHPRU) based at King’s College London and University College London. The MHPRU conducts research in response to policymaker need (e.g., in the Department for Health and Social Care or NHS England). A working group comprising academic and lived experience researchers, and clinicians met regularly throughout the course of the project.

## Eligibility criteria

Our review’s inclusion and exclusion criteria are described below. A table summary is available in Appendix D.

### Population

Patients of any age, gender and ethnicity were included. Staff, family members/carers or non-mental health patients were excluded.

### Setting

We included reviews that focused on care delivered within non-forensic inpatient mental health settings, including acute and longer-term inpatient services, and emergency departments. We excluded reviews focused on forensic inpatient mental health services, non-psychiatric medical inpatient services, services specifically for people with intellectual disabilities or autistic people, neurorehabilitation services, services specifically for people living with dementia, and community-based services.

### Intervention

We included relational care approaches to assessing and managing self-harm and suicide risk in inpatient mental health and emergency department settings. These approaches must include a focus on interpersonal relationships and involve at least some of the values and/or principles described in our co-produced definition of ‘relational care’, provided above. We excluded pharmacological interventions, surveillance technologies, restrictive interventions (e.g., physical restraint, seclusion room use, rapid tranquilisation), structured risk assessment checklists and risk stratification, approaches focused only on the physical design of the environment, and standard aspects of inpatient mental health and ED care (e.g., psychosocial assessments, ward rounds).

### Outcomes

We included reviews that examined self-harm and/or suicide-related outcomes, such as measures of suicidal ideation, frequency of self-harm or suicide attempts, time to next self-harm or suicide attempt, and rates of completed suicides. We excluded reviews that focused solely on risks to or from others, other patient outcomes, or staff or carer outcomes.

### Types of studies

We opted to scope published reviews rather than primary research studies, due to preliminary literature searches revealing numerous existing reviews on the effectiveness of interventions for assessing and managing self-harm and suicide in inpatient mental health and ED settings.

Quantitative and mixed-methods reviews were eligible for inclusion, including systematic, scoping, integrative, rapid, and narrative reviews. Both peer-reviewed and non-peer-reviewed sources were eligible for inclusion. We excluded primary research studies, books, commentaries, editorials, PhD/MSc/BSc theses, opinion pieces, blog posts and social media content. We applied no date restrictions but only included studies published in English. These restrictions were applied to narrow our scope, ensuring this review could be completed within the required timescales.

## Literature searching

We searched three academic databases (Medline, PsycINFO and CINAHL) for reviews examining the impact of relational care approaches on self-harm and suicide-related outcomes in inpatient mental health and ED settings. Database searches were conducted on 11/06/24 and were limited to review articles. No date or language search restrictions were applied.

Our search strategy included key terms relating to ‘relational care’ and ‘relational practice’ as well as terms to search more generally for approaches to assessing and managing self-harm or suicide risk in inpatient mental health and ED settings. The search terms were drafted by JG and further refined through consultation with the working group. The full search terms used can be seen in Appendix E. The results of the database searches were exported into Endnote and duplicates were removed.

Additional relevant literature was also identified through searching Google Scholar, the National Institute for Health and Care Excellence (NICE) website, reference and citation lists of included reviews, and recommendations from members of our working group.

## Selection of sources of evidence

All studies identified through database searches were independently double screened at title and abstract (JG, UF, RS). 10% of full texts were independently double screened (JG, UF). Any disagreements during screening were resolved through discussion. Screening was conducted in Rayyan (49). Studies identified through searching Google Scholar, the NICE website, expert recommendations and forwards and backwards citation searching were screened by JG and RS.

## Data charting and data items

Two data extraction forms were developed in Microsoft Word and collaboratively revised with the working group. The first form summarised the eligible reviews, including their design, aims, search strategies, eligibility criteria, identified relational care approaches, and overall conclusions. The second form summarised each of the relevant primary studies in these reviews, including information about their designs, locations, samples, interventions, any control/comparison groups, and reported quantitative evidence for the impact of the relational care intervention on self-harm and suicide-related outcomes. Data were extracted into these forms by two researchers (JG, RS), and all entries were double-checked for accuracy. Disagreements were resolved through discussion. No systematic quality appraisal of the included reviews or primary studies was conducted.

## Synthesis

Synthesis was led by two researchers (JG, RS), with input from the working group. The characteristics and findings of the included reviews were tabulated (Appendix G) and summarised narratively.

Similarly, the characteristics and results of relevant primary studies within these reviews were tabulated and narratively described, grouped by setting and relational care approach. Only quantitative evidence for the impact of relational care approaches on self-harm or suicide-related outcomes was synthesised.

More detailed tables and narrative descriptions summarising evidence from primary studies are provided in the appendices (see Supplementary File 1 for relational care approaches in inpatient mental health settings, and Supplementary File 2 for ED settings).

## Results

Database searches returned 2,424 studies. After removing duplicates, 2,118 records remained for title and abstract screening. 2,064 studies were excluded, leaving 52 studies for full-text screening. Additional search methods identified 18 studies. Overall, 29 reviews met our inclusion criteria and were included in this scoping review. A list of studies excluded at full-text screening, with reasons for their exclusion, are provided in Appendix F. Figure 1 presents the PRISMA flow diagram (50). A table of included review characteristics is available in Appendix G.

**Figure 1.**
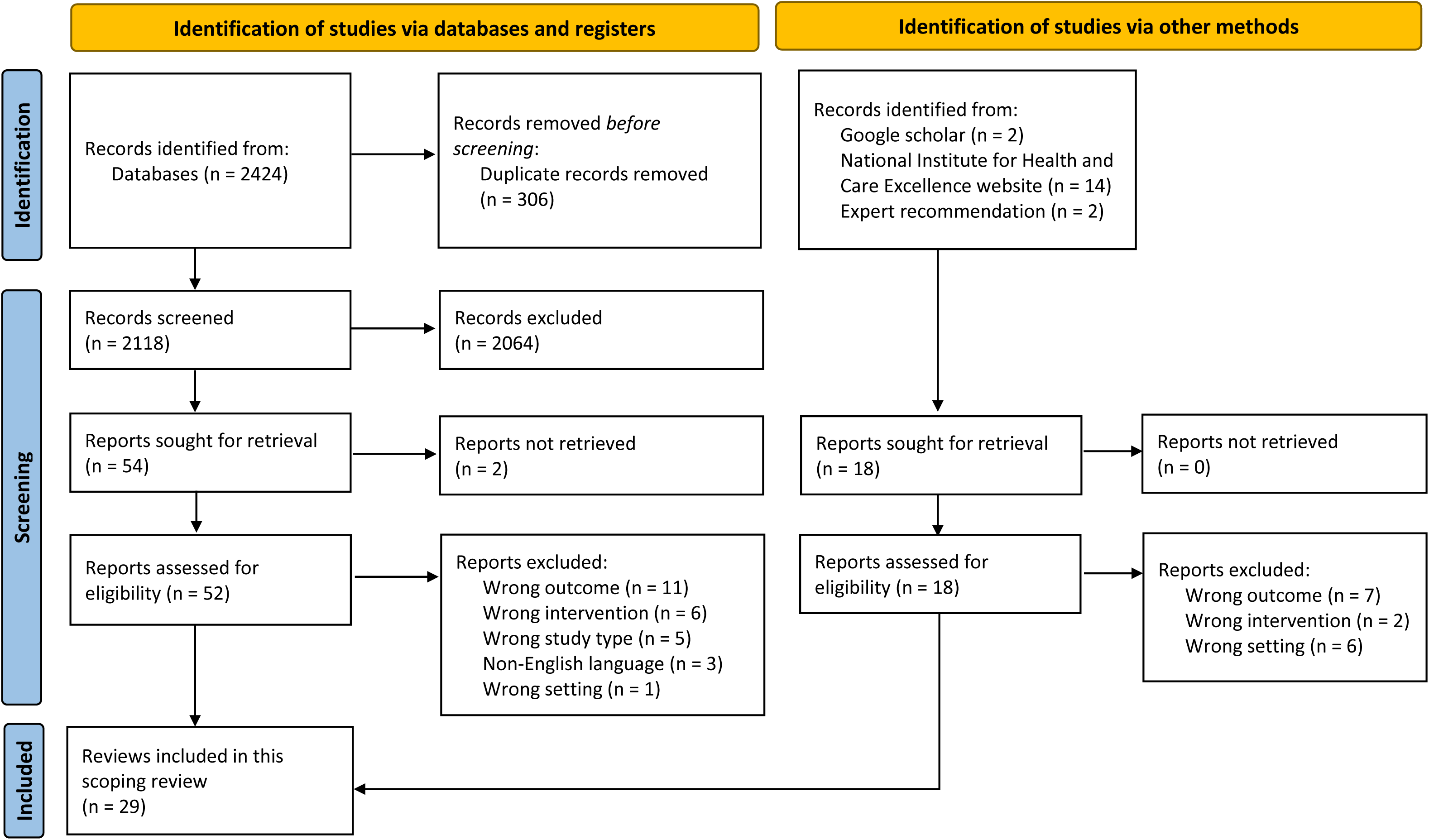
PRISMA flow diagram

### Characteristics of included reviews

All reviews identified studies by searching academic databases. Thirteen reviews also searched grey literature sources (e.g., clinical trial registries, Google Scholar, ResearchGate, relevant governmental and non-governmental websites, contacted authors for unpublished research) (51–63). Search strategies and eligibility criteria were not clearly stated in one review (64).

Out of the 29 included reviews, there was one systematic review with meta-analysis (51), 14 systematic reviews without meta-analyses (52–57,65–72), two rapid reviews (58,73), one integrative review (74), two scoping reviews (59,60), and nine non-systematic narrative reviews (61–64,75–79). Eighteen of the reviews focused on inpatient mental health settings only (51,52,55–58,62–68,74–77,79), six focused on emergency department settings only (53,59,60,69,71,73), and five included inpatient and ED settings (54,61,70,72,78). Eighteen reviews included self-harm and suicide as outcomes of interest (51–56,60–62,65,67,68,70,71,74–76,80), three reviews included self-harm only (59,66,77), and eight reviews included suicide only (58,63,64,69,73,78,79).

None of the included reviews used the term ‘relational’ to describe the interventions they examined. They captured ‘relational’ approaches by either searching broadly for any intervention for assessing and/or managing self-harm or suicide risk, or by specifically investigating ‘non-pharmacological’, ‘non-restrictive’, ‘psychological’, or ‘psychosocial’ interventions. There was considerable overlap in the primary studies included in the reviews.

### Characteristics of primary papers

In the 29 included reviews, 87 relevant primary papers were identified, reporting on 82 primary studies. 32 (39.0%) primary studies were conducted in the USA (81–114), 22 (26.8%) in the UK (80,115–136), 4 (4.9%) in Ireland (137–140), 4 (4.9%) in Germany (141–144), 4 (4.9%) in France (145–148), 3 (3.7%) in Canada (149–151), 3 (3.7%) in Switzerland (152–154), 2 (2.4%) in Australia (155,156), and 1 (1.2%) each in New Zealand (157), French Polynesia (158), Japan (159), Taiwan (160), South Korea (161), Spain (162,163), and Italy (164). One study (1.2%) had sites in Brazil, India, Sri Lanka, Iran and China (165,166). This shows that most of the included primary studies on relational care approaches were conducted in high-income countries, the majority in the USA and UK.

Overall, 49 primary papers reported on adult samples, 20 on CYP samples, 12 on adult and CYP samples, and six did not specify the age of participants. More detailed breakdowns of sample ages by primary study are provided in Table 1 and Table 2.

**Table 1.** Overview of relational care approaches identified and their impact on self-harm and suicide-related outcomes in non-forensic <u>inpatient mental health settings</u>.

| Primary study | Design | Intervention | Age group | Effect on self-harm | Effect on suicide |
| --- | --- | --- | --- | --- | --- |
| Dialectical behaviour therapy-based approaches |  |  |  |  |  |
| Springer 1996* (81) | RCT | Adapted inpatient DBT | Adults | No significance testing | Positive |
| Bohus 2000 (141) | Prospective pilot without control | Adapted inpatient DBT | Adults & CYP | Positive | Not measured |
| Kleindienst 2008 (143) | Naturalistic follow up without control | Adapted inpatient DBT | Adults | Positive | Not significant |
| Bohus 2004 (142) | Non-randomised trial | Adapted inpatient DBT | Adults | Positive | Not measured |
| Gibson 2014 (137) | Non-randomised trial | Adapted inpatient DBT | Adults | Positive | Not measured |
| Katz 2004 (149) | Non-randomised trial | Adapted inpatient DBT | CYP | Positive | Positive |
| Edel 2017 (144) | Pilot study with control | Adapted inpatient DBT | Adults | Not significant | Not measured |
| Barley 1993 (82) | Pre-post with control | Adapted inpatient DBT | Adults & CYP | Positive | Not measured |
| McDonell 2010 (83) | Pre-post with historic control | Adapted inpatient DBT | CYP | Positive | Not measured |
| Tebbett-Mock 2020 (84) | Pre-post with historic control | Adapted inpatient DBT | CYP | Positive | Positive |
| Booth 2014 (138) | Pre-post without control | Adapted inpatient DBT | Adults | Positive |  |
| Cognitive behaviour therapy-based approaches |  |  |  |  |  |
| Bentley 2017* (85) | Proof of concept RCT | Unified Protocol for the Transdiagnostic Treatment of Emotional Disorders | Adults | Not measured | Not significant |
| LaCroix 2018* (86) | Pilot RCT | Post-Admission Cognitive Therapy (PACT) | Adults | Not measured | Negative |
| Ghahramanlou-Holloway 2020* (87) | Pilot RCT | Post-Admission Cognitive Therapy (PACT) | Adults | Not measured | Not significant |
| Haddock 2019* (115) | RCT | Cognitive-behavioural suicide prevention (CBSP) therapy | Adults | Not measured | Not significant |
| Liberman & Eckman, 1981* (88) | RCT | Behavioural therapy | Adults | Not measured | Positive |
| Patsiokas & Clum, 1985* (89) | RCT | Cognitive restructuring | Not reported | Not measured | Not significant |
|  |  | Problem-solving therapy (PST) |  | Not measured | Not significant |
| Alesiani 2014 (164) | Pre-post without controls | Systems Training for Emotional Predictability and Problem Solving (STEPPS) therapy | Adults | Positive | Positive |
| Other psychological approaches |  |  |  |  |  |
| Celano 2017 (90) | RCT | Phone-based positive psychology | Adults | No significance testing | No significance testing |
|  |  | Cognition-focused intervention |  | No significance testing | No significance testing |
| Lieberman & Eckman, 1981 (88) | RCT | Insight-oriented psychotherapy | Adults | Not measured | Positive |
| Katz 2004 (149) | Non-randomised trial | Psychodynamic-oriented crisis assessment and treatment | CYP | Positive | Positive |
| Edel 2017 (144) | Pilot study with controls | Combined DBT and MBT group therapies | Adults | Positive | Not measured |
| Ellis 2012 (91) | Open trial, case-focused design without control | Collaborative Assessment and Management of Suicidality (CAMS) | Adults | Not measured | Positive |
| Ellis 2015 (92) | Naturalistic non-randomised comparison trial with control | Collaborative Assessment and Management of Suicidality (CAMS) | Adults | Not measured | Positive |
| Berrino 2011 (152) | Cohort study with control | Brief admission crisis intervention program | Adults | No significance testing | Positive |
| Yen 2019 (93) | Pre-post without control | Steps to Enhance Positivity (STEPs) therapy | CYP | Not measured | Positive |
| **Staff training** |  |  |  |  |  |
| Bowers 2006 (116) | Before-and-after trial without controls | City nurses | Adults | Positive | Not significant |
| Bowers 2008 (134) | RCT | City nurses | Not reported | Not significant |  |
| Ercole-Fricke 2016 (94) | Quasi-experimental without controls | Collaborative problem-solving training for nurses | CYP | Positive | Not measured |
| **Observations** |  |  |  |  |  |
| Stewart 2009 (117) | Longitudinal analysis without controls | Constant observations | Adults | Not significant | Not measured |
| Bowers 2003 (118) | Cross-sectional | Constant observations | Adults | Negative | Not measured |
| Bowers 2008 (119) | Cross-sectional | Constant observations | Adults | Not significant | Not measured |
|  |  | Intermittent observation |  | Positive | Not measured |
| Bowers 2011 (121) | Cross-sectional | Intermittent observation | Adults and CYP | Not measured | Positive |
| Stewart & Bowers 2012 (120) | Cross-sectional | Intermittent observation | Adults | Positive | Not measured |
| Stewart, Bowers & Ross 2012 (122) | Cross-sectional | Constant observations | Adults | No significance testing | No significance testing |
| **Ward- and organisational-level approaches** |  |  |  |  |  |
| Bowers 2015 (123) | Pragmatic cluster RCT | Safewards | Adults | Positive |  |
| Dickens 2020 (156) | Longitudinal pre-post without controls | Safewards | Adults | Positive |  |
| Reen 2020 (80) | Interrupted time series without controls | Twilight shifts and evening activities programme | CYP | Positive | Not measured |
| Gordon 2004 (124) | Pre-post with controls | Tidal model | Adults | No significance testing | Not measured |
| Fletcher & Stevenson 2001 (125) | Pre-post without controls | Tidal model | Adults | No significance testing | Not measured |
| Stevenson, Barker & Fletcher 2002 (126) | Pre-post without controls | Tidal model | Adults | No significance testing | No significance testing |
| Dodds & Bowles 2002 (127) | Pre-post without controls | Bradford Refocusing model | Adults | No significance testing | No significance testing |
| **Mixed interventions** |  |  |  |  |  |
| Pfeiffer 2019* (95) | RCT | Peer support and DBT strategies | Adults | Not measured | No significance testing |
| Berntsen 2011 (155) | Quantitative descriptive without controls | Staff training in DBT and seclusion and restraint, programme to reward patient behaviour, five patient exercise sessions per week | CYP | No significance testing | Not measured |
| **Other approaches** |  |  |  |  |  |
| Motto 1976 (96); Motto & Bostrom 2001 (97) | RCT | Caring letters | Adults | Not measured | Positive |
| Bennewith 2014 (135) | Pilot study without controls | Caring letters | Adults | No significance testing | No significance testing |
| Springer 1996 (81) | RCT | Wellness and lifestyle discussion group | Adults | Positive | Positive |
| Drew 2001 (98) | Retrospective correlational design with control | No-suicide contracts | Adults | Negative | Negative |
| Potter 2005 (99) | Pre-post without controls | Safety agreement tool/contract | Adults | Not significant |  |

The 'self-harm' column summarises the effect of the relational care approach in each primary study on self-harm outcomes, including self-harm frequency, severity, and frequency of presentations to services for self-harm. The 'suicide' column similarly summarises the effect of the relational care approach in each primary study on suicide-related outcomes, such as completed suicides, suicide attempts, suicidal ideation, and presentations to services for suicidality. In both columns, 'positive' and green shading indicates significant improvement in the outcome, 'negative' and red shading significant negative impact, and 'not significant' and yellow shading no significant effect. 'No significance testing' and grey shading indicates a lack of statistical analysis, and 'not measured' and grey shading shows that the outcome was not measured in the primary study. The 'effect on self-harm' and 'effect on suicide' columns are merged in studies where no distinction was made between suicidal and non-suicidal self-injury. \* Indicates that the study was included in Yiu et al.'s (2021) (51) systematic review and meta-analysis of psychosocial interventions in inpatient settings, which included 10 RCTs (examining DBT interventions, CBT interventions, and gratitude journalling), and concluded that psychosocial interventions were not any more effective than control interventions. RCT = Randomised Controlled Trial.

**Table 2.**
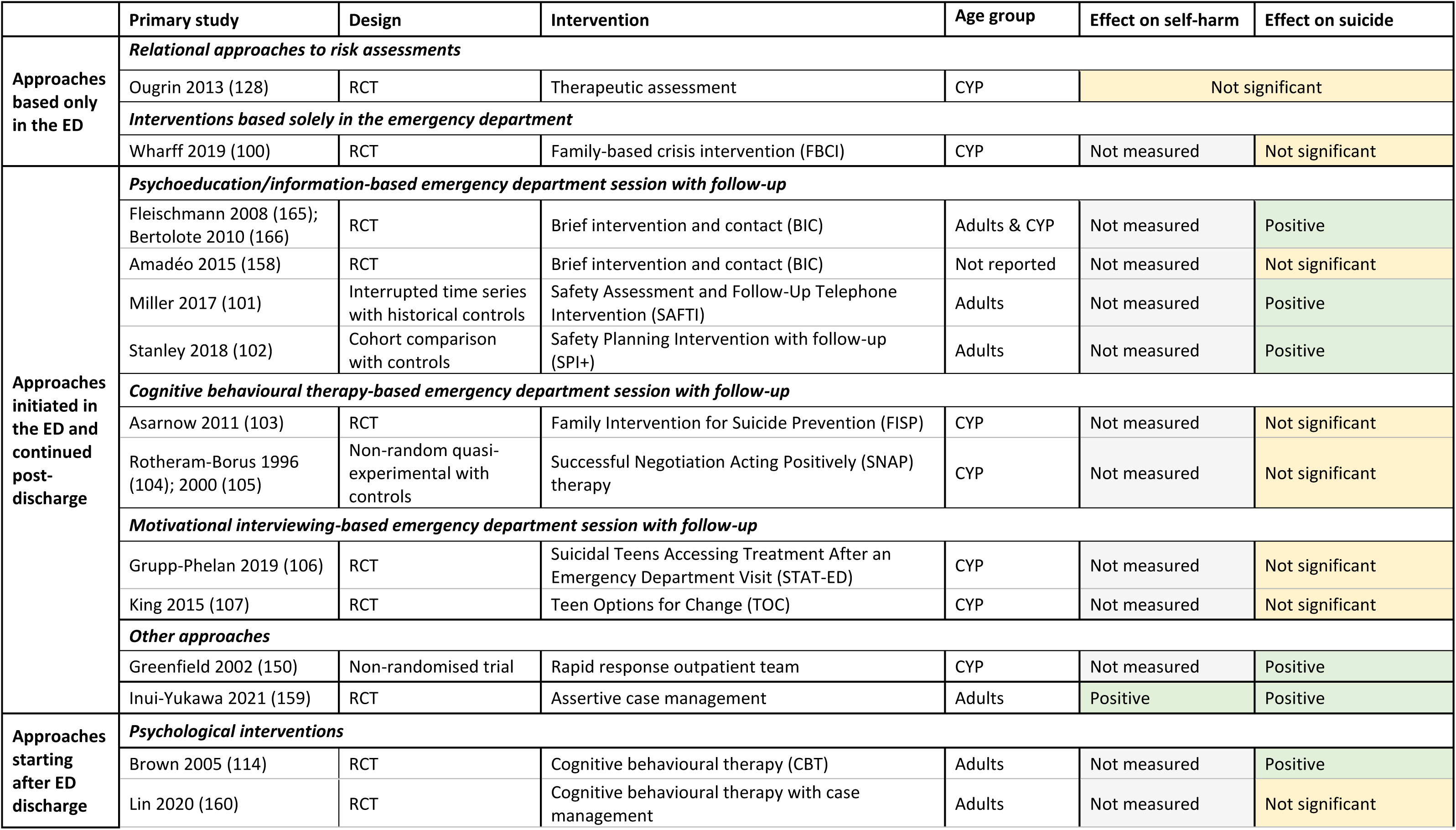
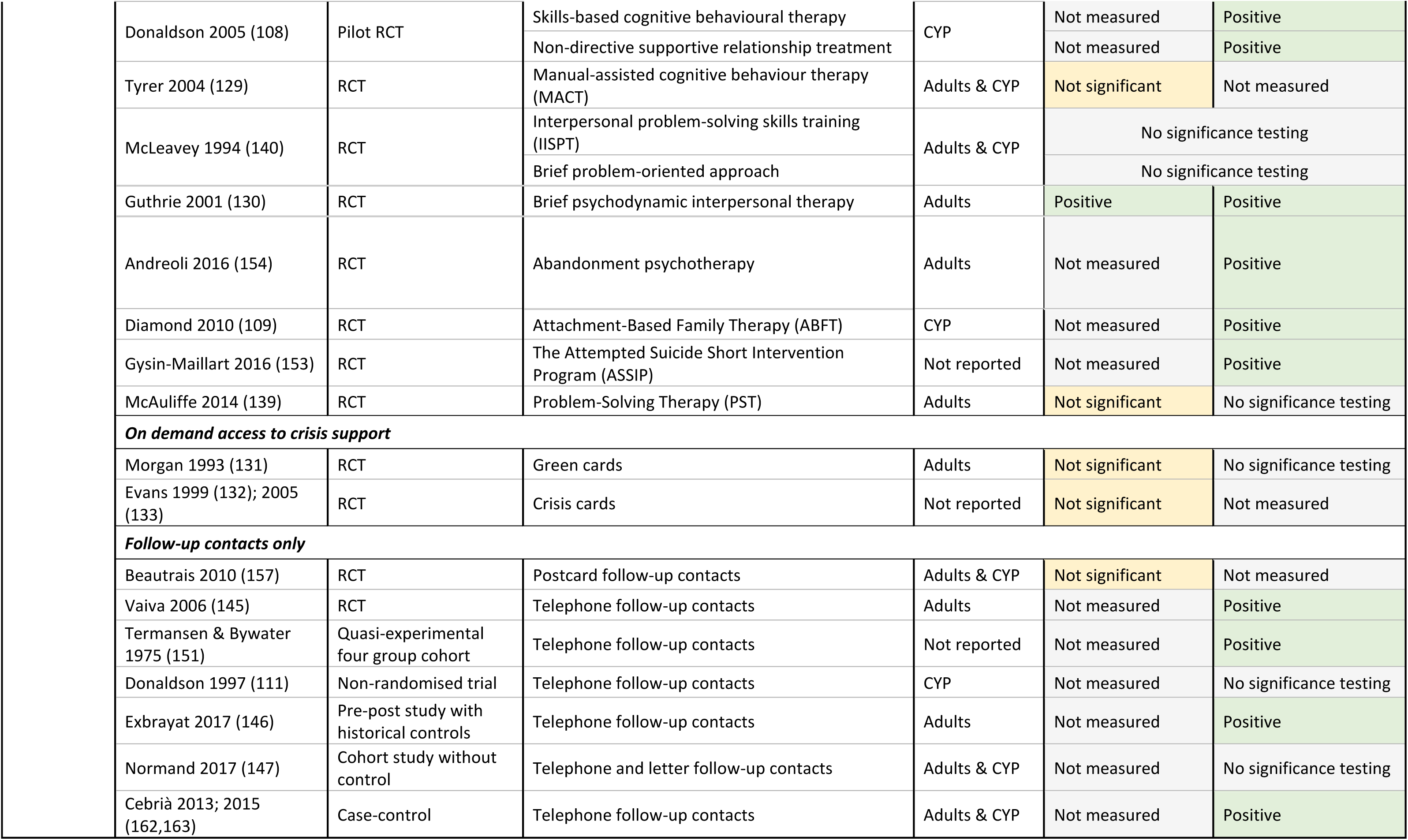
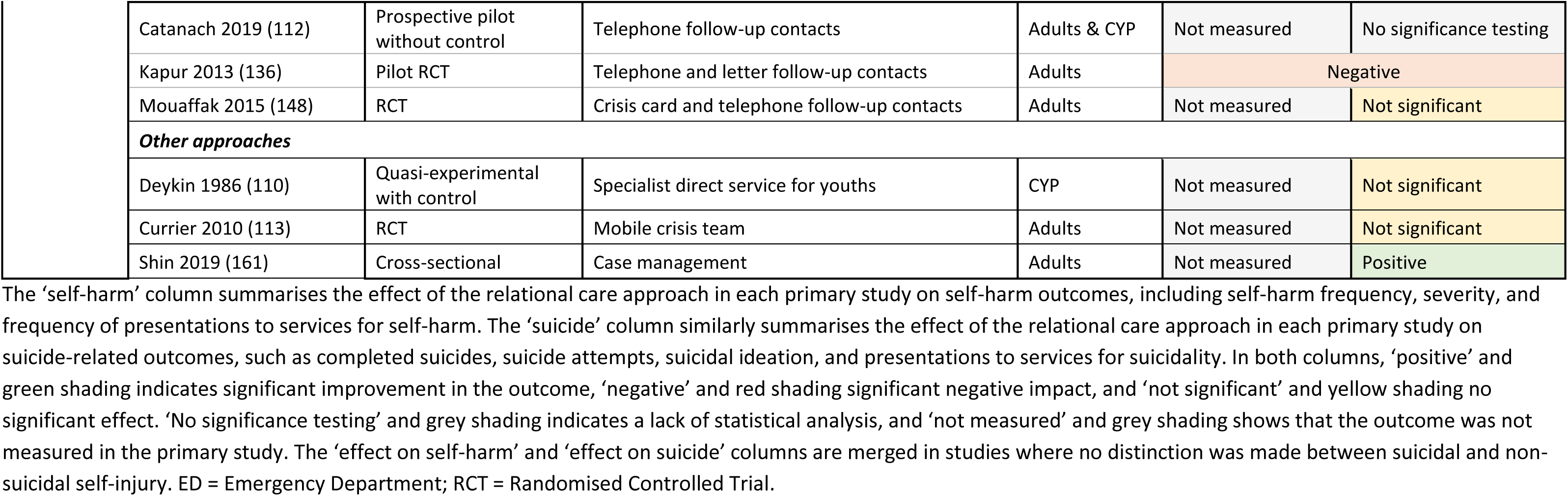
Overview of relational care approaches identified and their impact on self-harm and suicide-related outcomes in <u>emergency department settings</u>.

|  | Primary study | Design | Intervention | Age group | Effect on self-harm | Effect on suicide |
| --- | --- | --- | --- | --- | --- | --- |
| Approaches based only in the ED | Relational approaches to risk assessments |  |  |  |  |  |
|  | Ougrin 2013 (128) | RCT | Therapeutic assessment | CYP | Not significant |  |
|  | Interventions based solely in the emergency department |  |  |  |  |  |
|  | Wharff 2019 (100) | RCT | Family-based crisis intervention (FBCI) | CYP | Not measured | Not significant |
| Approaches initiated in the ED and continued post-discharge | Psychoeducation/information-based emergency department session with follow-up |  |  |  |  |  |
|  | Fleischmann 2008 (165); Bertolote 2010 (166) | RCT | Brief intervention and contact (BIC) | Adults & CYP | Not measured | Positive |
|  | Amadéo 2015 (158) | RCT | Brief intervention and contact (BIC) | Not reported | Not measured | Not significant |
|  | Miller 2017 (101) | Interrupted time series with historical controls | Safety Assessment and Follow-Up Telephone Intervention (SAFTI) | Adults | Not measured | Positive |
|  | Stanley 2018 (102) | Cohort comparison with controls | Safety Planning Intervention with follow-up (SPI+) | Adults | Not measured | Positive |
|  | Cognitive behavioural therapy-based emergency department session with follow-up |  |  |  |  |  |
|  | Asarnow 2011 (103) | RCT | Family Intervention for Suicide Prevention (FISP) | CYP | Not measured | Not significant |
|  | Rotheram-Borus 1996 (104); 2000 (105) | Non-random quasi-experimental with controls | Successful Negotiation Acting Positively (SNAP) therapy | CYP | Not measured | Not significant |
|  | Motivational interviewing-based emergency department session with follow-up |  |  |  |  |  |
|  | Grupp-Phelan 2019 (106) | RCT | Suicidal Teens Accessing Treatment After an Emergency Department Visit (STAT-ED) | CYP | Not measured | Not significant |
|  | King 2015 (107) | RCT | Teen Options for Change (TOC) | CYP | Not measured | Not significant |
|  | Other approaches |  |  |  |  |  |
|  | Greenfield 2002 (150) | Non-randomised trial | Rapid response outpatient team | CYP | Not measured | Positive |
|  | Inui-Yukawa 2021 (159) | RCT | Assertive case management | Adults | Positive | Positive |
| Approaches starting after ED discharge | Psychological interventions |  |  |  |  |  |
|  | Brown 2005 (114) | RCT | Cognitive behavioural therapy (CBT) | Adults | Not measured | Positive |
|  | Lin 2020 (160) | RCT | Cognitive behavioural therapy with case management | Adults | Not measured | Not significant |

|  |  |  |  |  |  |
| --- | --- | --- | --- | --- | --- |
| Donaldson 2005 (108) | Pilot RCT | Skills-based cognitive behavioural therapy | CYP | Not measured | Positive |
|  |  | Non-directive supportive relationship treatment |  | Not measured | Positive |
| Tyrer 2004 (129) | RCT | Manual-assisted cognitive behaviour therapy (MACT) | Adults & CYP | Not significant | Not measured |
| McLeavey 1994 (140) | RCT | Interpersonal problem-solving skills training (IISPT) | Adults & CYP | No significance testing |  |
|  |  | Brief problem-oriented approach |  | No significance testing |  |
| Guthrie 2001 (130) | RCT | Brief psychodynamic interpersonal therapy | Adults | Positive | Positive |
| Andreoli 2016 (154) | RCT | Abandonment psychotherapy | Adults | Not measured | Positive |
| Diamond 2010 (109) | RCT | Attachment-Based Family Therapy (ABFT) | CYP | Not measured | Positive |
| Gysin-Maillart 2016 (153) | RCT | The Attempted Suicide Short Intervention Program (ASSIP) | Not reported | Not measured | Positive |
| McAuliffe 2014 (139) | RCT | Problem-Solving Therapy (PST) | Adults | Not significant | No significance testing |
| ***On demand access to crisis support*** |  |  |  |  |  |
| Morgan 1993 (131) | RCT | Green cards | Adults | Not significant | No significance testing |
| Evans 1999 (132); 2005 (133) | RCT | Crisis cards | Not reported | Not significant | Not measured |
| ***Follow-up contacts only*** |  |  |  |  |  |
| Beautrais 2010 (157) | RCT | Postcard follow-up contacts | Adults & CYP | Not significant | Not measured |
| Vaiva 2006 (145) | RCT | Telephone follow-up contacts | Adults | Not measured | Positive |
| Termansen & Bywater 1975 (151) | Quasi-experimental four group cohort | Telephone follow-up contacts | Not reported | Not measured | Positive |
| Donaldson 1997 (111) | Non-randomised trial | Telephone follow-up contacts | CYP | Not measured | No significance testing |
| Exbrayat 2017 (146) | Pre-post study with historical controls | Telephone follow-up contacts | Adults | Not measured | Positive |
| Normand 2017 (147) | Cohort study without control | Telephone and letter follow-up contacts | Adults & CYP | Not measured | No significance testing |
| Cebrià 2013; 2015 (162,163) | Case-control | Telephone follow-up contacts | Adults & CYP | Not measured | Positive |
| Catanach 2019 (112) | Prospective pilot without control | Telephone follow-up contacts | Adults & CYP | Not measured | No significance testing |
| Kapur 2013 (136) | Pilot RCT | Telephone and letter follow-up contacts | Adults | Negative |  |
| Mouaffak 2015 (148) | RCT | Crisis card and telephone follow-up contacts | Adults | Not measured | Not significant |
| **Other approaches** |  |  |  |  |  |
| Deykin 1986 (110) | Quasi-experimental with control | Specialist direct service for youths | CYP | Not measured | Not significant |
| Currier 2010 (113) | RCT | Mobile crisis team | Adults | Not measured | Not significant |
| Shin 2019 (161) | Cross-sectional | Case management | Adults | Not measured | Positive |

The 'self-harm' column summarises the effect of the relational care approach in each primary study on self-harm outcomes, including self-harm frequency, severity, and frequency of presentations to services for self-harm. The 'suicide' column similarly summarises the effect of the relational care approach in each primary study on suicide-related outcomes, such as completed suicides, suicide attempts, suicidal ideation, and presentations to services for suicidality. In both columns, 'positive' and green shading indicates significant improvement in the outcome, 'negative' and red shading significant negative impact, and 'not significant' and yellow shading no significant effect. 'No significance testing' and grey shading indicates a lack of statistical analysis, and 'not measured' and grey shading shows that the outcome was not measured in the primary study. The 'effect on self-harm' and 'effect on suicide' columns are merged in studies where no distinction was made between suicidal and non-suicidal self-injury. ED = Emergency Department; RCT = Randomised Controlled Trial.

Sixty-two relevant relational care approaches were identified which had been evaluated in terms of their impact on self-harm and/or suicide risk in inpatient or ED settings, across the 87 primary papers. Many of these were psychological interventions delivered at individual or group levels.

However, some ward- and organisation-level approaches were also identified. The primary studies reporting on them varied in design, from RCTs and controlled studies, to pre-post and cross-sectional studies.

Thirty different relational care approaches were identified from the included reviews which had been quantitatively examined in terms of their impact on self-harm and/or suicide-related outcomes in <u>inpatient mental health settings</u>, in 46 primary papers (see Table 1 for an overview).

Thirty-two different relational care approaches were identified from the included reviews which had been quantitatively examined in terms of their impact on self-harm and/or suicide-related outcomes in <u>ED settings</u>, in 41 primary papers (see Table 2 for an overview).

## Overall conclusions of the reviews

Overall, recurrent themes in the conclusions of the reviews included: a lack of high-quality evidence for the impact of these interventions on self-harm and suicide in inpatient mental health and ED settings; some interventions and their underlying theoretical assumptions and mechanisms of change are poorly described; a lack of consistency in methods and outcomes measured across studies; and a lack of lived experience involvement in the research. None of the reviews addressed how good relational care may be provided for neurodivergent individuals. This is important given that they often face barriers in accessing and benefiting from mental health care which can be mitigated with simple, reasonable adjustments (167,168). Nevertheless, the reviews did highlight some approaches with some supporting evidence for a positive change in key outcomes, summarised below.

### Inpatient settings

We identified a systematic review and meta-analysis by Yiu et al. (2021) which included 10 RCTs evaluating psychosocial interventions in inpatient settings (including CBT, DBT and gratitude journalling) (51). It concluded that psychosocial interventions did not significantly reduce suicidal ideation or suicide attempts compared to controls post-intervention (95% CI = -0.38 to 0.10; p = 0.26) or at follow-up (95% CI = -0.15 to 0.59; p = 0.24) (51). However, it only included some of the primary studies identified in this scoping review, in part due to only including RCTs, whereas we included primary studies of any quantitative design.

Other reviews we identified in this setting provide some evidence suggesting that the following approaches can have a significant positive effect on self-harm: adapted inpatient DBT (in 9/11 studies) (82–84,137,138,141–143,149), combined DBT and MBT (in 1/1 studies) (144), STEPPS therapy (in 1/1 studies) (164), psychodynamic-oriented crisis assessment and treatment (in 1/1 studies) (149), city nurses (in 1/2 studies) (116,134), collaborative problem-solving training for nurses (in 1/1 studies) (94), intermittent observation (in 2/2 studies) (119,120), and twilight nursing shifts with an evening activities programme (in 1/1 studies) (80). Evidence also suggests that Safewards significantly reduces ‘conflict’ events (including self-harm and suicide attempts amongst other conflict events) (in 2/2 studies) (123,156).

There was also some evidence for a significant positive effect on suicide-related outcomes for adapted inpatient DBT (in 4/5 studies) (81,84,138,149), CAMS (in 2/2 studies) (91,92), STEPs (in 1/1 studies) (93), psychodynamic-oriented crisis assessment and treatment (in 1/1 studies) (149), insight-oriented psychotherapy (in 1/1 studies) (88), a wellness and lifestyle discussion group (in 1/1 studies) (81), a brief admission crisis program (in 1/1 studies) (152), intermittent observation (in 1/2 studies) (121), and post-discharge caring letters (in 1/2 studies) (96). Only 2/7 studies of CBT-based approaches in inpatient settings, investigating STEPPS (164) and behavioural therapy (88), showed a significant positive impact on suicide-related outcomes, the remaining studies either found no significant effect (85,87,89,115) or a significant negative effect (86).

There was some evidence that no-suicide contracts (in 1/1 studies) (98), constant observation (in 1/4 studies) (118), and post-admission cognitive therapy (in 1/2 studies) (86) can have a significant negative impact on self-harm and/or suicide-related outcomes in inpatient settings.

### Emergency department settings

In ED settings, there was some evidence that brief psychodynamic interpersonal therapy initiated after ED discharge (in 1/1 studies) (130) and assertive case management initiated in the ED and continued post-ED discharge (in 1/1 studies) (159) significantly reduced self-harm. Other relational care approaches either had no significant impact on self-harm (128,129,131–133,139,157) or their impact on self-harm was not investigated or not significance tested.

There was evidence that some approaches initiated in the ED and continued post-ED discharge can significantly improve suicide-related outcomes, including: Safety Assessment and Follow-up Telephone Intervention (SAFTI) (in 1/1 studies) (101), Safety Panning Intervention with follow-up (SPI+) (in 1/1 studies) (102), brief intervention and contact (BIC) (in 1/2 studies) (165,166), a rapid response outpatient team (in 1/1 studies) (150) and assertive case management (in 1/1 studies) (159).

There was also some evidence suggesting that the following relational care approaches initiated post-ED discharge significantly improve suicide-related outcomes: CBT-based interventions (in 2/5 studies) (108,114), non-directive supportive relationship treatment (in 1/1 studies) (108), brief psychodynamic interpersonal therapy (in 1/1 studies) (130), abandonment psychotherapy (in 1/1 studies) (154), Attachment-Based Family Therapy (ABFT) (in 1/1 studies) (109), the Attempted Suicide Short Intervention Program (ASSIP) (in 1/1 studies) (153), case management (in 1/1 studies) (161), and telephone follow-up contacts (in 4/6 studies) (145,146,151,162,163).

The remaining relational care approaches were either found to have no significant effect on suicide-related outcomes (100,103–107,110,113,128,148) or their impact on them was not investigated or not significance tested. One study found that combined letter and telephone follow-up contacts were associated with significantly worse suicide-related outcomes compared to usual care (136).

For a more detailed breakdown of primary study results for each relational care approach in inpatient mental health settings, see Supplementary File 1. For a more detailed breakdown of primary study results for each relational care approach in ED settings, see Supplementary File 2.

## Discussion

### Key findings

Our scoping review outlines a proposed universal definition of ‘relational care’ and synthesises quantitative evidence for relational care approaches to assessing and managing self-harm and suicide risk in non-forensic inpatient mental health and ED settings. Twenty-nine relevant reviews were identified reporting on 62 relevant relational care approaches. Many of these were psychological interventions delivered at individual or group levels. However, some ward- and organisation-level approaches were also identified. For most of the relational care approaches included, only one primary study was identified assessing its impact on self-harm and/or suicide in inpatient or ED settings.

It is important to acknowledge that none of the included reviews’ research questions explicitly used the term ‘relational care’ and they, therefore, included some primary studies that did and did not meet our eligibility criteria for relational care. Instead, the reviews within this report constructed research questions which used the terms ‘psychosocial’, ‘psychological’, ‘non-restrictive’, and ‘non-pharmacological’ approaches, with the ‘relational care’ aspects of these interventions being implicit, rather than the stated primary focus of the studies.

In inpatient settings, supporting evidence was identified from controlled studies for some psychological interventions, including adapted inpatient DBT, combined DBT and MBT, CAMS, psychodynamic-oriented crisis assessment and treatment, behavioural therapy, insight-oriented psychotherapy, a wellness and lifestyle discussion group, and a brief admission crisis program. Additionally, controlled studies suggest that Safewards and post-discharge ‘caring letters’ can reduce self-harm and/or suicide. Uncontrolled studies provide some evidence for STEPPs therapy, STEPs, intermittent observation, twilight nursing shifts with evening activities, and certain staff training approaches such as ‘city nurses’ and ‘collaborative problem-solving training for nurses’. There was a lack of evidence, or mixed evidence, regarding the impact of other relational care interventions on self-harm and suicide-related outcomes in inpatient settings. Evidence from a controlled study of no-suicide contracts and an uncontrolled study of constant observation suggests that they can have a significant negative impact on self-harm and/or suicide related outcomes.

In EDs, relational care approaches demonstrated mixed effectiveness. Evidence was identified from controlled studies suggesting that some psychological approaches (e.g., brief psychodynamic interpersonal therapy, abandonment psychotherapy, SAFTI, SPI+, BIC, ABFT, ASSIP, some CBT-based approaches, and non-directive supportive relationship treatment), rapid response outpatient teams, assertive case management, and post-discharge telephone contacts can have a significant positive impact on self-harm and/or suicide-related outcomes. An uncontrolled cross-sectional study provided evidence supporting a post-discharge case management intervention. Evidence from controlled studies indicated that therapeutic assessments, other psychological approaches, on-demand crisis support (e.g., crisis cards, green cards), a specialist direct service for youths, mobile crisis teams, postcard follow-up contacts, and combined crisis card and telephone follow-up contacts, do not have a significant effect on self-harm or suicide-related outcomes. Evidence from one controlled study suggested that combined telephone and letter follow-up contacts can significantly worsen self-harm and suicide-related outcomes.

Overall, the identified reviews highlighted a lack of high-quality research in this area, noting poorly described interventions and mechanisms of change and inconsistent methodologies and outcome measures in primary studies. However, it is essential to consider that absence of evidence is not evidence of a lack of value in these approaches. It may instead reflect some of the challenges in researching ‘relational care’ and its impact on self-harm and suicide in inpatient and ED settings, explored below.

## Challenges defining ‘relational care’

As identified earlier in this report, the term ‘relational care’ is not widely used within inpatient mental health academic research. This is despite the concept having a longstanding history and underpinning many clinical approaches in mental health, including in inpatient and ED settings (29,169,170). Reviews on ‘relational care’ in a mental health context are only just beginning to emerge. For example, Lamph et al. (in prep) are conducting a conceptual analysis of ‘relational practice’, drawing upon global, cross-sector papers to report some of its key components.

The concept of ‘relational care’ also extends beyond mental healthcare; it has been described and applied across a range of other contexts, including education, criminal justice, and social work. For example, in social work, ‘relational-based practice’ is seen as core to social workers’ interactions and roles, and it is also cited within a variety of mental health nursing education texts (171–173). Whilst the concept of relational care exists across different sectors, there is variation in how it is defined and understood by clinicians and service users. For example, different professions have different perspectives on what ‘relational care’ means and how it can be applied in their work, shaped by their professional identities and philosophical and training backgrounds. ‘Relational care’ can be understood and applied differently depending on cultural, contextual, and individual factors. This variability makes it difficult to define, operationalise and research.

## Challenges in defining and assessing fidelity to relational care values and principles

Another challenge is to evaluate fidelity to ‘relational care’. Some fundamental components such as respect, authenticity, and shared humanity, can be difficult to measure and depend on the personal qualities of individual health professionals. It is possible that a ‘relational care’ intervention could be delivered in a way that is perfunctory and inconsistent with the values and principles that underpin it. For example, verbal de-escalation encourages staff to validate patients’ emotional responses while empathising calmly and is a part of some relational care approaches. While intended to be supportive and comforting, there is a risk that it could be experienced as invalidating or a means of “providing a kinder façade to oppressive practice” (26). This complexity can make it difficult to operationalise and evaluate adherence to relational care approaches in research.

## Difficulties in measuring self-harm and suicide outcomes

Evaluating the impact of any intervention on self-harm and suicide rates in inpatient and ED settings is a challenge. While highly important, it must be considered that the numbers of suicides on inpatient wards remains, thankfully, a relatively rare occurrence (3). As a result, it is difficult to evaluate the impact of any intervention on preventing suicides without conducting large-scale studies on multiple wards (e.g., Bowers et al. (2008) (119)). Furthermore, the nature of suicidality and reasons people may engage in self-harming behaviours, as well as self-harm methods, are vast, variable, and may change drastically over time, making them difficult to measure. It can also be challenging to distinguish suicidal and non-suicidal self-injury (174). Whilst frequency of self-injury is a crude outcome measure, accounting for self-injury severity risks creating a problematic and potentially invalidating hierarchy of methods. The private nature of self-harm also means it is unlikely to be accurately measured. More restrictive approaches may keep people safer in the short term but cause long-term harm, such as physical and psychological injury, dehumanisation, erosion of trust between patients and staff, and (re)traumatisation (25,175). There is a need to be person-centred when approaching these topics, as what works to help keep some patients safe may be problematic for others. There is no standard ‘one size fits all’ approach for everyone and all services.

## The impact of many relational care approaches on self-harm and suicide has not been researched

There are many other relational care approaches used in inpatient and ED settings which were not captured by these reviews, and thus within this report, because they were not quantitatively evaluated in the academic literature in terms of their impact on self-harm or suicide. There is likely a bias in the research towards approaches such as DBT which were developed with an explicit and direct focus on reducing self-harm and suicide. It is notable that this review identified evidence supporting relational care interventions which take a less behavioural approach, for example, brief psychodynamic interpersonal therapy (130). Other therapies and approaches that also have positive effects in the long- or short-term are likely to exist, though their direct impact on self-harm and suicide may not have been evaluated in research and so they will not have been identified in this scoping paper.

Approaches that have an indirect impact on self-harm and suicide, including interventions aimed at changing ward cultures and environment may, therefore, be overlooked within these reviews. Such approaches include evidence-based approaches such as Safewards (56,156,176) and the Assured intervention (33). Other approaches include Open Dialogue (177,178), therapeutic communities (179,180), and Enabling Environments (181). These examples offer valuable insights into the potential benefits of relational care interventions, values, and practices which address systemic and cultural factors affecting self-harm and suicide risk management.

## Barriers and facilitators to implementing relational care approaches in these settings

While this scoping review found evidence for the use of some relational care approaches within inpatient and ED settings to reduce suicide and self-harm, it is important to acknowledge that consistently and effectively implementing relational care in these contexts is difficult. Whilst implementing complex interventions in any real-world setting is inherently challenging and requires careful consideration of active and dynamic factors that either facilitate or hinder implementation (182,183), these specialist settings introduce additional unique barriers.

Firstly, inpatient mental health and ED environments are dynamic with a diverse mix of different staff, patients, and visitors, each with their unique backgrounds and personalities. There are therefore many different relationships at play, between patients, between staff and patients, and between different staff. There may naturally be variability in the provision of relational care between services, wards, staff teams, and people on different shifts. Individuals with certain personal qualities (e.g., people who are caring, kind and empathetic) may provide relational care more naturally, whereas others may struggle to engage relationally. Furthermore, an individual’s capacity to provide relational care may vary over time, for example, depending on their personal circumstances and other factors such as stress levels, burnout, and other stressors (184). Navigating the boundary between demonstrating these qualities and maintaining safe boundaries and professional limitations also needs to be considered.

Secondly, providing relational care consistently in an inpatient or ED context is further complicated by the changing composition of staff and patients in these settings. Inconsistent shift patterns, high levels of unfilled vacancies (especially for registered nurses), reliance on bank and agency staff, and utilisation of more peripheral team members introduces variability. Patients themselves often have transient experiences in EDs and short stays in inpatient settings, and the NHS Mental Health Implementation Plan is aiming to reduce the length of inpatient psychiatric stays further, to a maximum of 32 days (185). These factors require careful consideration as they will impact both implementation of relational care at a personal level and influence the broader ward milieu and culture at a more ecological level.

Thirdly, inpatient mental health and ED settings are complex and coercive environments. Many patients – often the majority – are compulsorily detained and may experience interventions and restrictive practices against their will, leading to diminished autonomy and limited choices. There are therefore significant power imbalances between patients and staff, which no doubt create considerable barriers to implementing an intervention based on relationship equality, particularly within a hierarchical, authoritarian system (26).

Finally, it is crucial to remember that these are contexts where there are significant risks. Getting things wrong can have severe consequences, including physical and psychological harm to patients, devastation to families, and severe distress to staff. In ED settings, there is often a disproportionate focus on mental health presentations as the cause of violence and aggression. This can contribute to staff difficulty distinguishing clinical distress and agitation from actual violence and aggression, increasing staff anxiety and leading to a reliance on restrictive interventions to manage risk, thereby hindering the implementation of relational care. Front-facing staff in ED and inpatient settings who spend the most time with patients often receive the least training, are the lowest paid, and receive the least supervisory support (e.g., supervision and reflective practice). This can result in high levels of burnout and moral injury amongst staff (186). Furthermore, staff face pressure from hospital management, external regulatory agencies, and coroners to document risk assessments. This is in addition to the already substantial burden of administrative tasks, monitoring and reporting required of staff, which reduces time available for direct clinical care. These pressures faced by staff can hinder their ability to effectively implement person-centred, relational care and drive an over-reliance on risk assessment tools and restrictive practices, despite their ineffectiveness in managing risk (187).

## Strengths and limitations

This paper offers a broad overview of the quantitative evidence for relational care approaches to assessing and managing self-harm and suicide risk in inpatient mental health and ED settings. We have presented a coproduced comprehensive definition of ‘relational care’, laying the groundwork for future research in this area. This review is the result of a collaboration of academic and lived experience researchers and clinicians with expertise in the topic of relational care, ensuring representation of diverse expert perspectives.

However, this report also has some limitations. Firstly, we did not register a protocol a priori for this review. Future studies should consider protocol registration to enhance transparency and reproducibility. Secondly, due to time constraints, we did not systematically search grey literature. This may have limited the scope of the literature identified. However, many of the reviews that we identified did search grey literature (e.g., pre-print servers, Google Scholar, relevant websites, policy documents) more comprehensively. Thirdly, in line with PRISMA guidelines (48), we did not conduct any formal quality appraisal, limiting the certainty of conclusions about the strength of the evidence identified. Fourthly, although we conducted independent double screening of all sources at title/abstract and a subsample of full-texts, we did not perform formal double independent data extraction. However, all extracted data were double-checked for accuracy. Finally, qualitative evidence was not included in our synthesis due to time limitations. Further research incorporating it could provide insight into patient, staff and family/carer experiences and views of relational care approaches and, subjectively, what makes a positive difference (188,189).

## Implications for research, policy and practice

The current lack of a consistent definition of ‘relational care’ poses a significant challenge for both research and practice. Future research could aim to clarify the meaning of ‘relational care’, its core components, and develop a clear framework for its consistent application and evaluation.

Conceptualisations of ‘relational care’ should consider the influence of culture and context, including how it intersects with the needs of marginalised groups, such Black and ethnic minority groups, those facing language barriers, autistic individuals, and people with intellectual disabilities. This is crucial given the inequities that these groups experience in terms of access, experiences, and outcomes in acute mental healthcare (30,190–196). However, the consideration of culture and context should not be limited to marginalised groups; it should be a universal consideration for all patients, staff, services, and healthcare systems.

Further research is needed to evaluate the impact of relational care approaches on quality and safety in inpatient mental health and ED settings, including more large-scale RCTs and studies evaluating long-term outcomes (30). This includes research examining the impact of relational care on self-harm and suicide, as well as on other important outcomes such as psychological safety, self-neglect, physical health, iatrogenic harms, staff safety and wellbeing, therapeutic alliance, engagement with services (e.g., length of stay, readmission rates, other service use), and treatment satisfaction. Economic evaluations taking these broader outcomes into account are also needed; cost-effectiveness evidence is important for shaping policy and practice. Further research co-produced with patients, families/carers, staff, policymakers, and commissioners is needed to ensure research addresses the priorities of these key stakeholders.

Future research should also focus on understanding the barriers and facilitators of successfully implementing relational care approaches to assessing and managing self-harm and suicide risk in these settings, including consideration of training and support needs for staff. Furthermore, realist approaches could help to determine what works for whom, in what circumstances, and why (197). This could enable relational care approaches to be more effectively adapted and tailored to different contexts and populations, including those underrepresented in research studies (30).

Given the complexity of research in this area there is a considerable need for qualitative studies to explore patient, staff, and family/carer experiences of relational care approaches. Personal stories from qualitative studies could help to understand how relational care can be provided authentically, rather than performatively. Whilst some primary qualitative studies were identified in this scoping exercise, synthesising their findings was beyond our scope. Synthesis of this qualitative literature, and further qualitative research, would help to understand the nuances in both the delivery and experience of these interventions.

While this scoping exercise highlighted a general lack of high-quality evidence for relational care approaches, research has shown that many common practices in inpatient mental health and ED settings are not supported by the evidence, for example, structured risk assessments, no-suicide contracts, and constant observations. It can be argued that it is preferable to implement approaches based on the principles of relational care whilst continuing to develop its evidence base than continue to use approaches with evidence of harm.

## Conclusion

This scoping review proposes a co-produced definition of ‘relational care’ and identifies supporting evidence for some relational care approaches to assessing and managing self-harm and suicide risk in inpatient mental health and ED settings, including a variety of individual-, group-, and organisation-level approaches. However, further high-quality research, including larger-scale RCTs, is required to evaluate their effectiveness and long-term impact. Co-produced research is needed to clarify the definition, core components, and develop a framework for applying and evaluating ‘relational care’. Future studies should also focus on understanding barriers and facilitators to implementing relational care and incorporate qualitative methods to capture the perspectives of patients, staff, and carers.

## Lived experience commentary by Raza Griffiths, Tamar Jeynes and Lizzie Mitchell

This Lived Experience Commentary comes from the perspective of wanting to strengthen lived experience voices in policy research and positively impacting practice, by ensuring that research reflects the priorities service users themselves have highlighted. In this regard we would like to highlight the following points about this paper.

The paper concentrates on developing the idea of ‘relational care’ and using it to assess and manage suicidality and self-harm. But the impetus for developing the idea of “relational care” does not seem to have come from people with lived experience. The idea itself is innocuous, encapsulating standard tropes about how workers should ideally relate to service users. This semantic repackaging suggests some exciting new developments, whereas in all probability, it may simply become another ‘buzzword’ to mask a lack of real change, as happened with earlier concepts like “recovery” and “trauma informed”.

On a practical level, there were difficulties in reviewing literature defining ‘relational care’ differently, and using various methods of measuring, recording and evaluating services. How are staff and services meant to adhere to a standard where there isn’t a set definition?

Moreover, the studies reviewed self-defined how ‘relational’ their services were, based on their own definition of services, rather than asking how *we* as service users rated them in terms of relational care.

Even more than this: shouldn’t we as service users, be defining what the ideal characteristics of the way staff relate to us should be, rather than using a rubric on what is important which has been developed by someone else? Reviews should not be reinforcing knowledge from research studies which exclude Lived Experience voices.

In its definition of relational care, the paper foregrounds interpersonal relationships, which are crucial and can be therapeutic in themselves. However, relationships exist within powerful political, systemic and cultural constraints and unequal power dynamics, which the paper does not focus on. The bigger picture needs to be addressed, including the impact of severe understaffing and long waiting lists.

A key cultural challenge to relational ways of working, is the reliance on coercive practices, which sits diametrically opposite relational ways of working. Widespread and controversial use of control and restraint in inpatient services is a point of ongoing debate and campaigning within mental health, with the United Nations Convention on the Rights of Persons with Disabilities being an important rallying point for us and our allies. It argues for a move away from biomedical coercive approaches to ones which could be broadly defined as ‘relational’. But will it be possible to mainstream a relational approach in the current system, or can it only ever be tokenistic, given the nature of the mental health system?

Finally, the review highlights a reduction in suicides in inpatient care between 2010 – 2020. The broader context outside wards, however, was of a steep rise in suicide, which was correlated with the financial squeeze, a more onerous benefits regime and cutbacks to mental health services. This highlights the need to focus on the wider social context, entailing joined up action from diverse organisations and central government addressing wider social determinants of self-harm and suicide.

## Data availability statement

All data used is publicly available in the published papers included in this study.

## Ethics approval and consent to participate

Not applicable.

## Consent for publication

Not applicable.

## Acronyms

A&E: Accident and Emergency
ABFT: Attachment-Based Family Therapy
ASSIP: The Attempted Suicide Short Intervention Program
BPD: Borderline Personality Disorder
BIC: Brief Intervention and Contact
CAMS: Collaborative Assessment and Management of Suicidality
CBSP: Cognitive-Behavioural Suicide Prevention Therapy
CBT: Cognitive Behaviour Therapy
CCTV: Closed-Circuit Television
CYP: Children and Young People
DBT: Dialectical Behaviour Therapy
ED: Emergency Department
FBCI: Family-Based Crisis Intervention
FISP: Family Intervention for Suicide Prevention
HCP: Healthcare Professional
IISPT: Interpersonal Problem-Solving Skills Training
ISRCTN: International Standard Randomised Controlled Trial Number
LGBTIQ: Lesbian, Gay, Bisexual, Transgender, Intersex, Queer or Questioning
MACT: Manual-Assisted Cognitive Behaviour Therapy
MBT: Mentalisation-Based Therapy
MHPRU: Policy Research Unit in Mental Health
NHS: National Health Service
NIHR: National Institute for Health and Care Research
NICE: National Institute for Health and Care Excellence
NSSI: Non-Suicidal Self Injury
RCT: Randomised Controlled Trial
SAFTI: Safety Assessment and Follow-Up Telephone Intervention
SNAP: Successful Negotiation Acting Positively therapy
SPI+: Safety Planning Intervention with follow-up
STAT-ED: Suicidal Teens Accessing Treatment After an Emergency Department Visit
STEPPS: Systems Training for Emotional Predictability and Problem Solving therapy
STEPS: Steps to Enhance Positivity therapy
TOC: Teen Options for Change
UK: United Kingdom
USA: United States of America

## Footnotes

1 When referencing this definition, please cite this paper as follows: [add citation]. Our definition draws upon existing definitions and descriptions of ‘relational care’ in the literature (35,37–44) (see Appendix A). An expanded definition is provided in Appendix B.

## Appendices

## Appendix A: Definitions drawn upon in coproducing a working definition of ‘relational care’

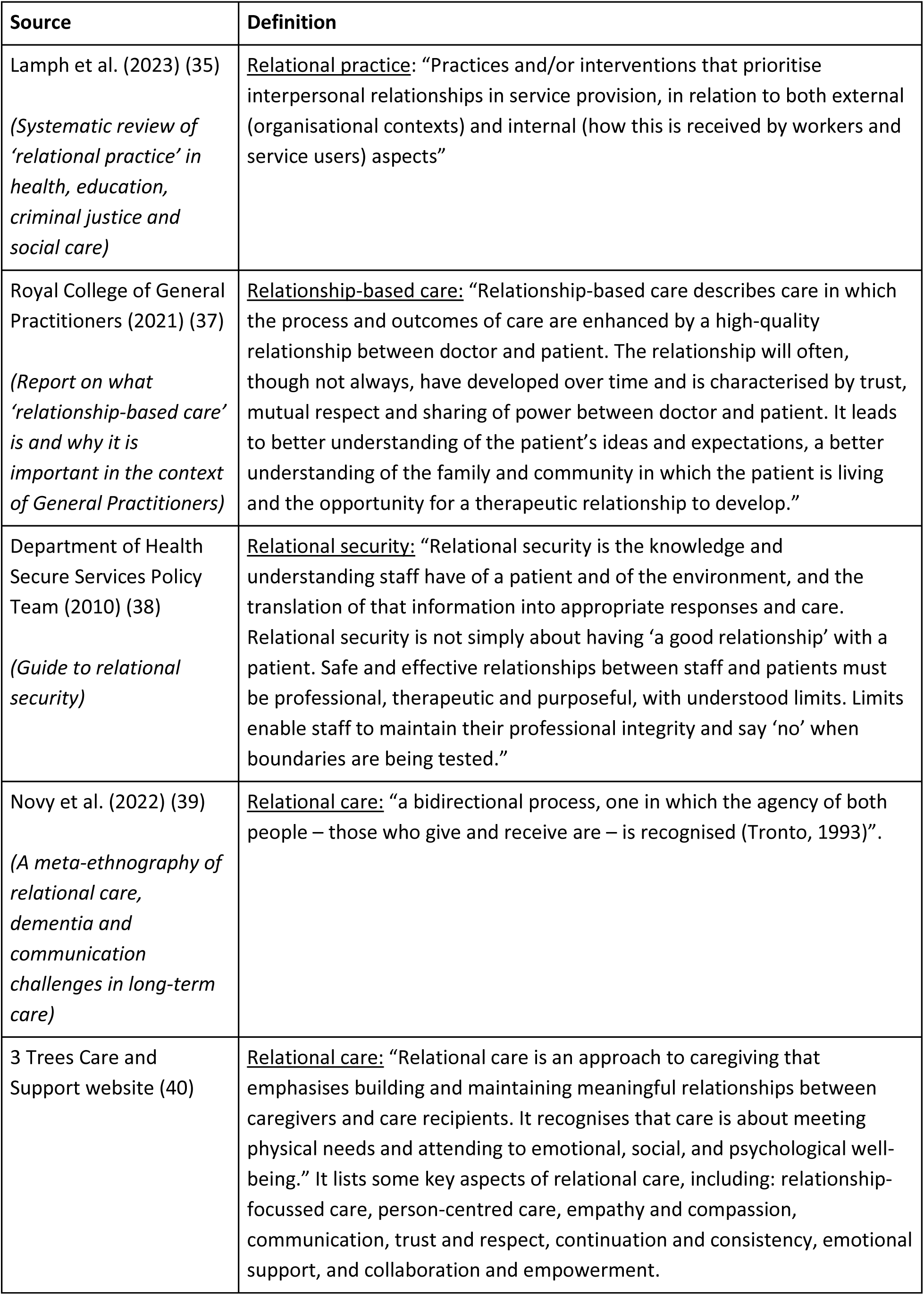
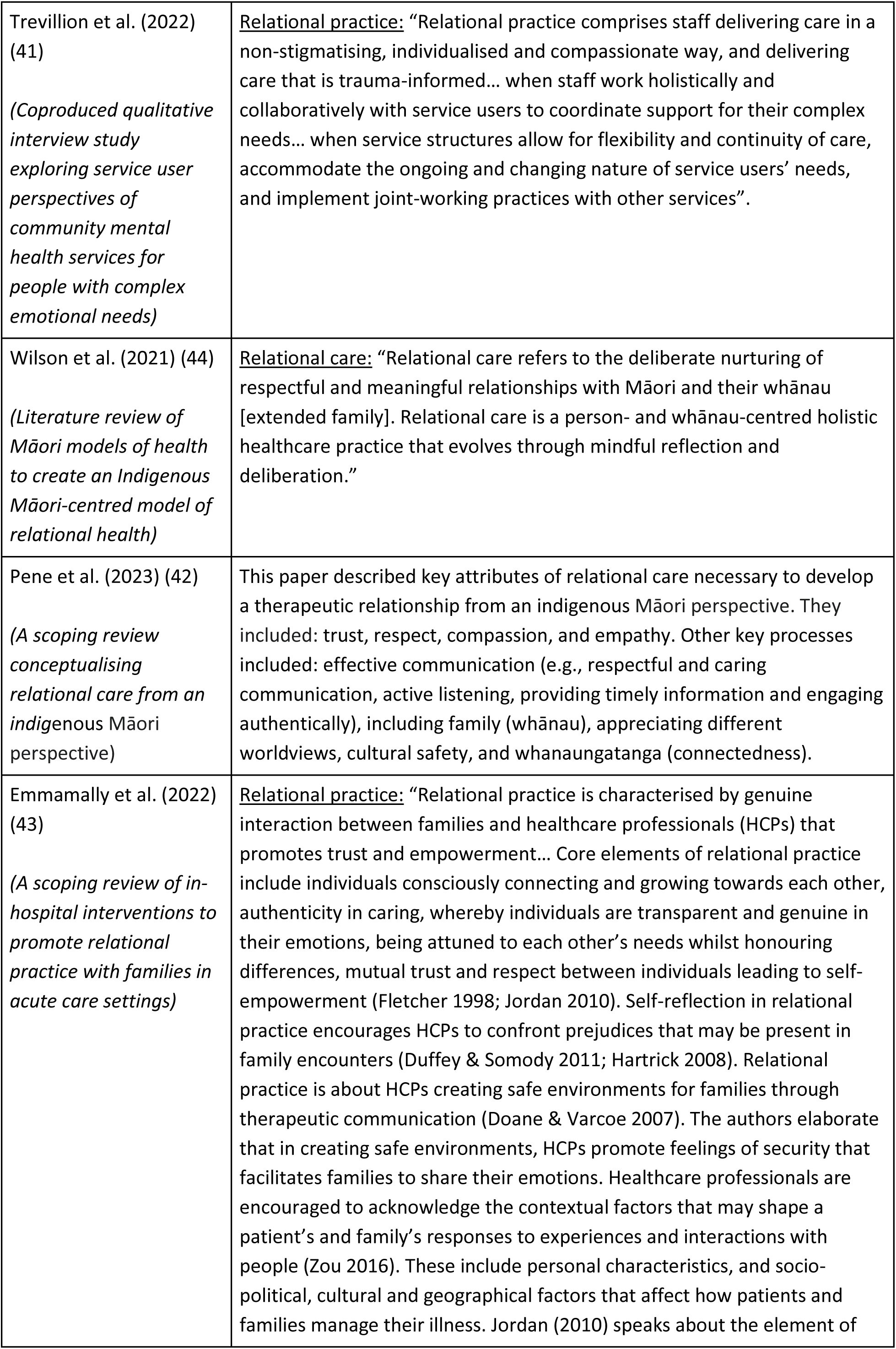
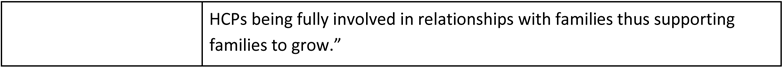

Appendix A: Definitions drawn upon in coproducing a working definition of 'relational care'
| Source | Definition |
| --- | --- |
| Lamph et al. (2023) (35)<br>*(Systematic review of 'relational practice' in health, education, criminal justice and social care)* | <u>Relational practice</u>: "Practices and/or interventions that prioritise interpersonal relationships in service provision, in relation to both external (organisational contexts) and internal (how this is received by workers and service users) aspects" |
| Royal College of General Practitioners (2021) (37)<br>*(Report on what 'relationship-based care' is and why it is important in the context of General Practitioners)* | <u>Relationship-based care</u>: "Relationship-based care describes care in which the process and outcomes of care are enhanced by a high-quality relationship between doctor and patient. The relationship will often, though not always, have developed over time and is characterised by trust, mutual respect and sharing of power between doctor and patient. It leads to better understanding of the patient's ideas and expectations, a better understanding of the family and community in which the patient is living and the opportunity for a therapeutic relationship to develop." |
| Department of Health Secure Services Policy Team (2010) (38)<br>*(Guide to relational security)* | <u>Relational security</u>: "Relational security is the knowledge and understanding staff have of a patient and of the environment, and the translation of that information into appropriate responses and care. Relational security is not simply about having 'a good relationship' with a patient. Safe and effective relationships between staff and patients must be professional, therapeutic and purposeful, with understood limits. Limits enable staff to maintain their professional integrity and say 'no' when boundaries are being tested." |
| Novy et al. (2022) (39)<br>*(A meta-ethnography of relational care, dementia and communication challenges in long-term care)* | <u>Relational care</u>: "a bidirectional process, one in which the agency of both people – those who give and receive are – is recognised (Tronto, 1993)". |
| 3 Trees Care and Support website (40) | <u>Relational care</u>: "Relational care is an approach to caregiving that emphasises building and maintaining meaningful relationships between caregivers and care recipients. It recognises that care is about meeting physical needs and attending to emotional, social, and psychological well-being." It lists some key aspects of relational care, including: relationship-focussed care, person-centred care, empathy and compassion, communication, trust and respect, continuation and consistency, emotional support, and collaboration and empowerment. |
| Trevillion et al. (2022) (41)<br>*(Coproducted qualitative interview study exploring service user perspectives of community mental health services for people with complex emotional needs)* | <u>Relational practice</u>: “Relational practice comprises staff delivering care in a non-stigmatising, individualised and compassionate way, and delivering care that is trauma-informed... when staff work holistically and collaboratively with service users to coordinate support for their complex needs... when service structures allow for flexibility and continuity of care, accommodate the ongoing and changing nature of service users’ needs, and implement joint-working practices with other services”. |
| Wilson et al. (2021) (44)<br>*(Literature review of Māori models of health to create an Indigenous Māori-centred model of relational health)* | <u>Relational care</u>: “Relational care refers to the deliberate nurturing of respectful and meaningful relationships with Māori and their whānau [extended family]. Relational care is a person- and whānau-centred holistic healthcare practice that evolves through mindful reflection and deliberation.” |
| Pene et al. (2023) (42)<br>*(A scoping review conceptualising relational care from an indigenous Māori perspective)* | This paper described key attributes of relational care necessary to develop a therapeutic relationship from an indigenous Māori perspective. They included: trust, respect, compassion, and empathy. Other key processes included: effective communication (e.g., respectful and caring communication, active listening, providing timely information and engaging authentically), including family (whānau), appreciating different worldviews, cultural safety, and whanaungatanga (connectedness). |
| Emmamally et al. (2022) (43)<br>*(A scoping review of in-hospital interventions to promote relational practice with families in acute care settings)* | <u>Relational practice</u>: “Relational practice is characterised by genuine interaction between families and healthcare professionals (HCPs) that promotes trust and empowerment... Core elements of relational practice include individuals consciously connecting and growing towards each other, authenticity in caring, whereby individuals are transparent and genuine in their emotions, being attuned to each other’s needs whilst honouring differences, mutual trust and respect between individuals leading to self-empowerment (Fletcher 1998; Jordan 2010). Self-reflection in relational practice encourages HCPs to confront prejudices that may be present in family encounters (Duffey & Somody 2011; Hartrick 2008). Relational practice is about HCPs creating safe environments for families through therapeutic communication (Doane & Varcoe 2007). The authors elaborate that in creating safe environments, HCPs promote feelings of security that facilitates families to share their emotions. Healthcare professionals are encouraged to acknowledge the contextual factors that may shape a patient’s and family’s responses to experiences and interactions with people (Zou 2016). These include personal characteristics, and socio-political, cultural and geographical factors that affect how patients and families manage their illness. Jordan (2010) speaks about the element of |
|  | HCPs being fully involved in relationships with families thus supporting families to grow.” |

## Appendix B: Expanded definition of ‘relational care’ co-produced by our working group of academic and lived experience researchers and clinicians

Relational care can be practised at individual, group, organisational or systemic levels. It relates to <u>how</u> care is delivered, rather than the specific content or format of interventions. Relational care prioritises interpersonal relationships, acknowledging their central role in effective treatment and recovery. It is grounded in values such as respect, dignity, empathy, humility, authenticity, compassion, empowerment, trust, and shared humanity. Relational care is guided by principles that include: understanding individuals within the context of their lives, providing personalised and holistic care, promoting cultural safety, fostering effective communication, believing in patients and inspiring hope. It is also guided by the principle of democratisation – actively involving patients and the people close to them (e.g., family, friends, partners) in decisions about their care and the functioning of the care environment. This requires power imbalances to be acknowledged and addressed.

## Appendix C: Preferred Reporting Items for Systematic reviews and Meta-Analyses extension for Scoping Reviews (PRISMA-ScR) Checklist

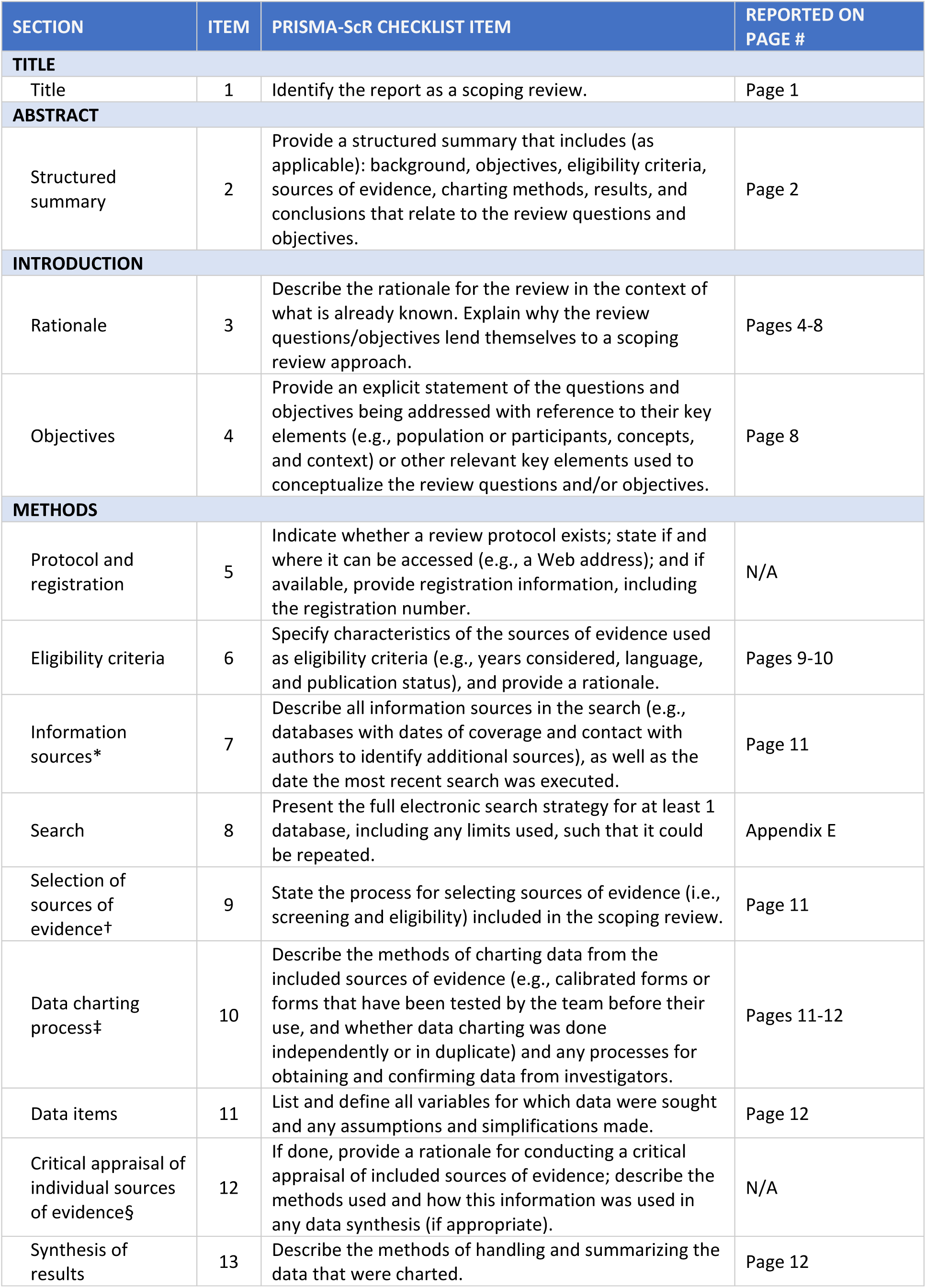
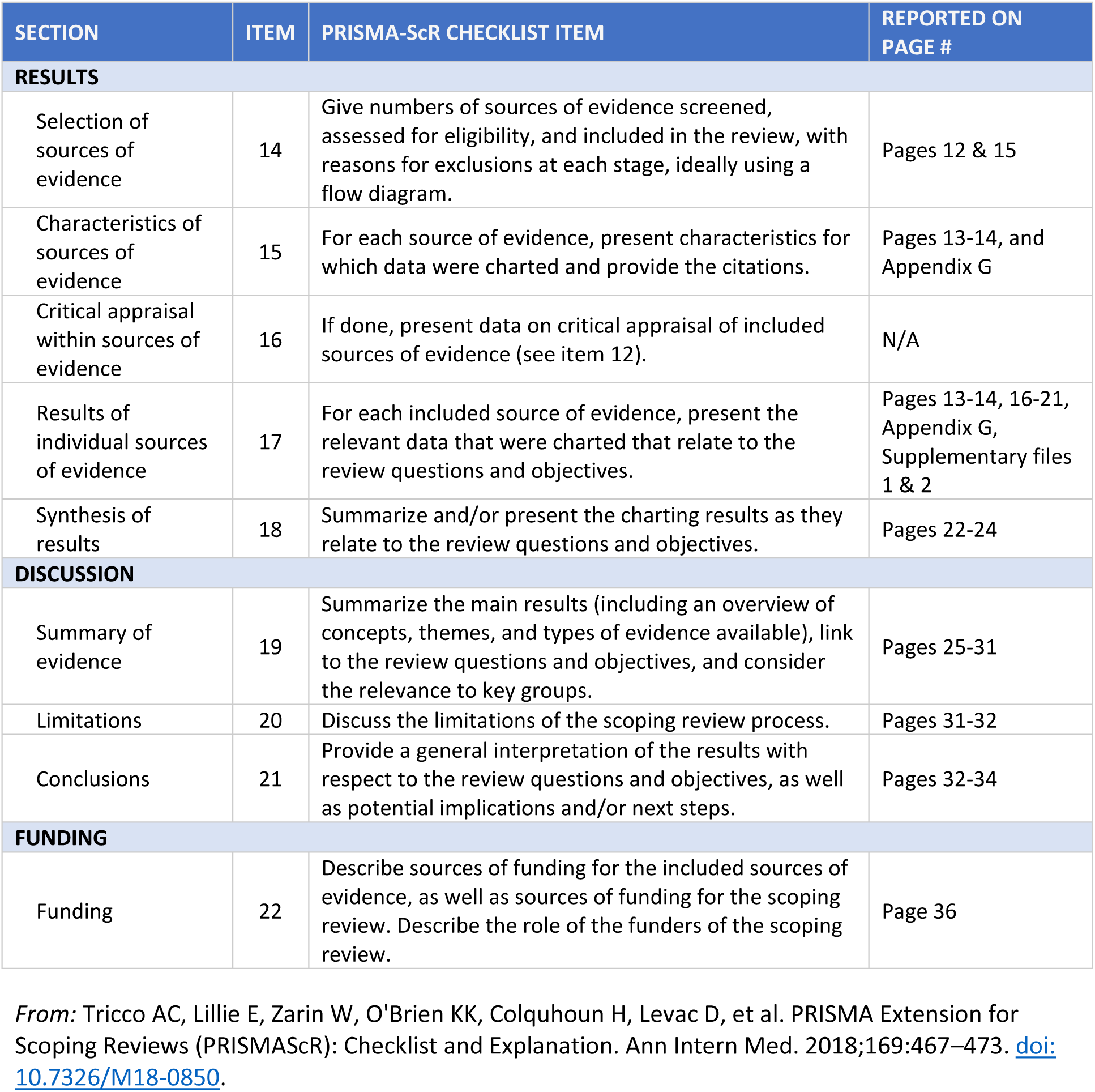

## Appendix D: Inclusion and exclusion criteria for reviews in this report

|  | Included | Excluded |
| --- | --- | --- |
| **Population** | Mental health patients (of any age, ethnicity, sex, or gender) | Reviews only including staff or family/carers, or non-mental health patients |
| **Intervention/ approach** | Relational care approaches to assessing and managing self-harm and suicide risk in inpatient mental health and emergency department settings. These approaches must include a focus on interpersonal relationships and involve at least some of the values and/or principles outlined in the definition of 'relational care' (see above). | Pharmacological interventions<br>Surveillance technologies<br>Restrictive interventions (e.g., seclusion room use, rapid tranquilisation, physical restraint)<br>Structured risk assessment checklists and risk stratification<br>Standard aspects of inpatient mental health or emergency department care (e.g., ward rounds, psychosocial assessments)<br>Approaches focusing only on the physical design of the environment |
| **Comparators/ controls** | Reviews examining any comparator/control groups were eligible to be included<br>Reviews of studies with no comparator/control groups | None |
| **Outcomes** | Self-harm (e.g., frequency, severity)<br>Suicide (e.g., suicidal ideation, suicide attempt frequency, time to suicide attempts, completed suicides) | Risk to others<br>Risk from others<br>Other patient outcomes<br>Staff outcomes<br>Carer outcomes |
| **Setting** | Non-forensic inpatient mental health settings (including acute and longer-term inpatient services)<br>Emergency departments | Forensic inpatient mental health services<br>Services specifically for people with an intellectual disability<br>Services specifically for autistic people<br>Non-psychiatric medical inpatient services<br>Services specifically for people living with dementia<br>Neurorehabilitation wards<br>Community-based services |
| **Study type** | Reviews (e.g., systematic reviews, scoping reviews, rapid reviews, narrative reviews)<br>Peer-reviewed and non-peer reviewed reviews<br>Reviews published any date<br>Reviews published in English<br>Studies conducted in any country | Primary research studies<br>Books<br>Commentaries<br>Editorials<br>PhD/MSc/BSc theses<br>Opinion pieces<br>Blog posts<br>Social media content<br>Non-English language papers |

## Appendix E: Search strings

1. (Psychiatri* or “mental health”).mp.
2. (inpatient or hospital* or ward* or facility* or unit* or PICU or “136-suite” or “136 suite” or “place* of safety” or emergency department* or A&E).mp.
3. (Intervention* or approach* or strateg* or program* or manag* or protocol* or therap* or initiative* or mileu* or environment* or anti* or prevent* or improv* or trauma-informed or trauma informed or safeguard* or protect* or precaution* or reduc* or mitigat* or secur* or risk assessment* or model* or train* or policy* or policies* or leadership* or activit* or group* or session* or practice* or treatment* or QI or project* or peer or counselling* or de-escalat* or skill* or technique* or implement* or meeting* or communit* or scheme*).mp.
4. (Suicid* or ligature* or ligation or hang* or strangle* or strangulation* or asphyxi* or parasuicid* or self-harm* or self harm* or self-injur* or self injur* or self-mutilat* or self mutilat* or DSH or NSSI or self-poison* or self poison* or incident* or safety).mp.
5. 1 and 2 and 3 and 4
6. limit 5 to “review articles”

## Appendix F: Excluded full texts and reasons for exclusion

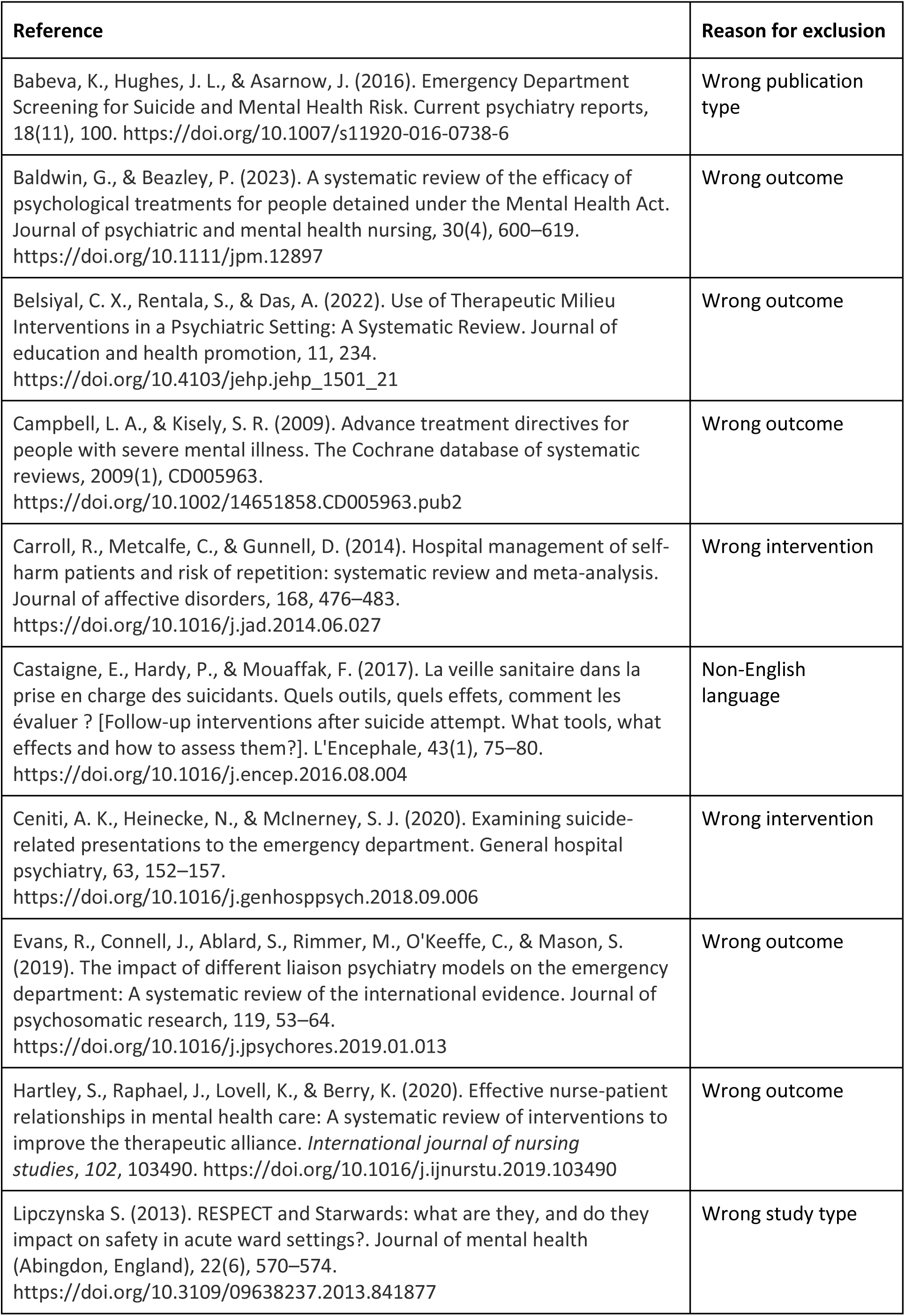
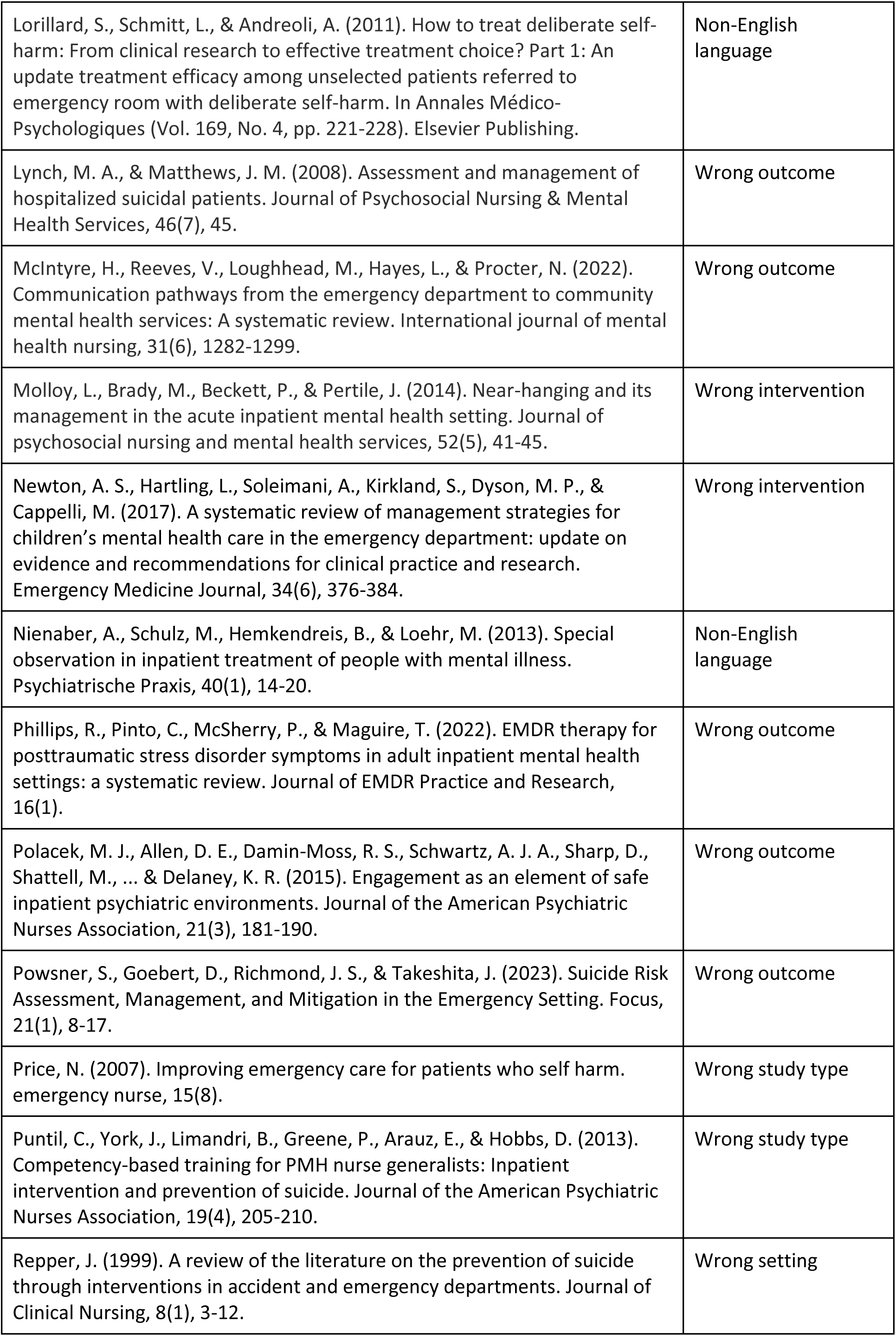
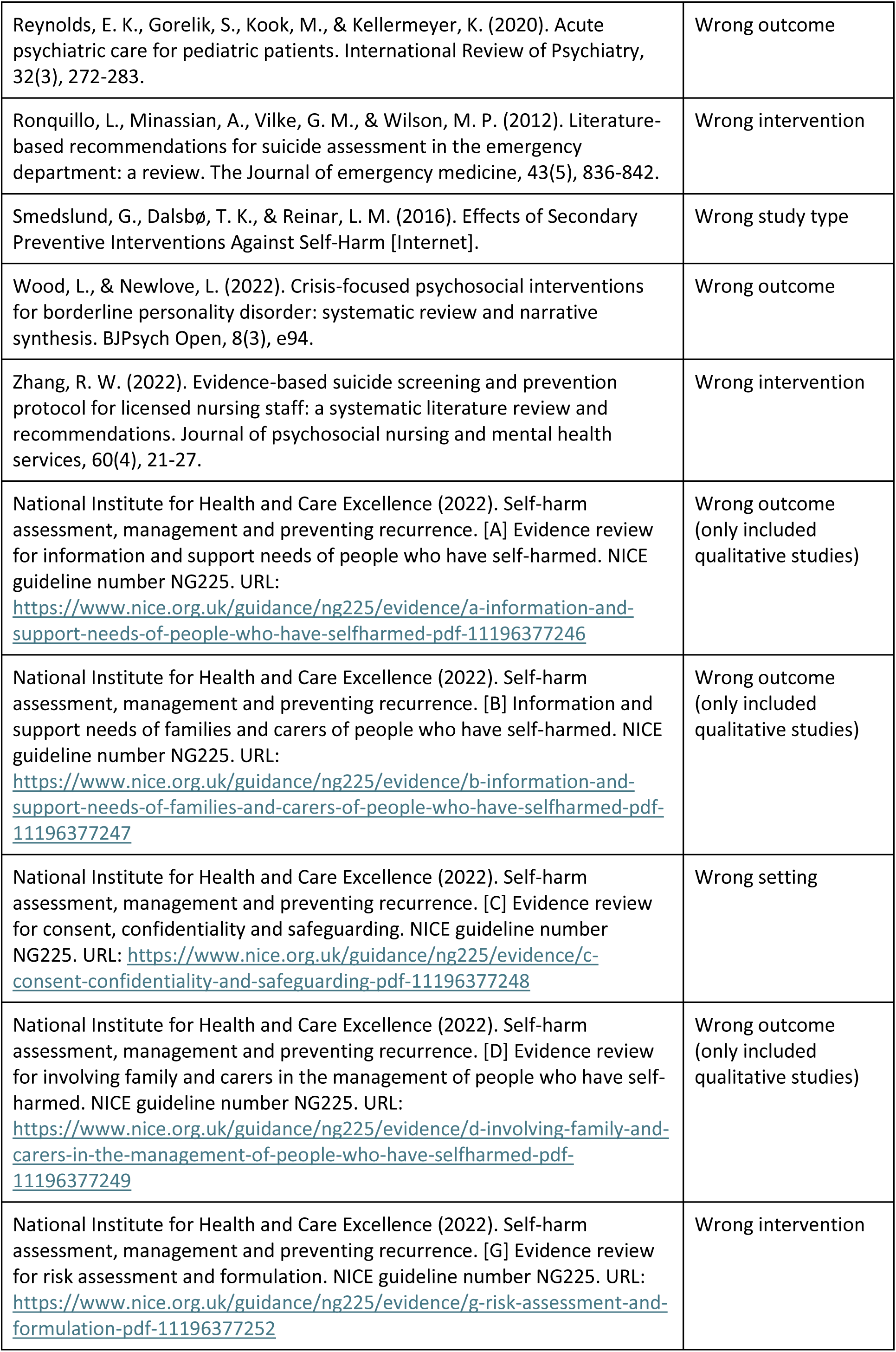
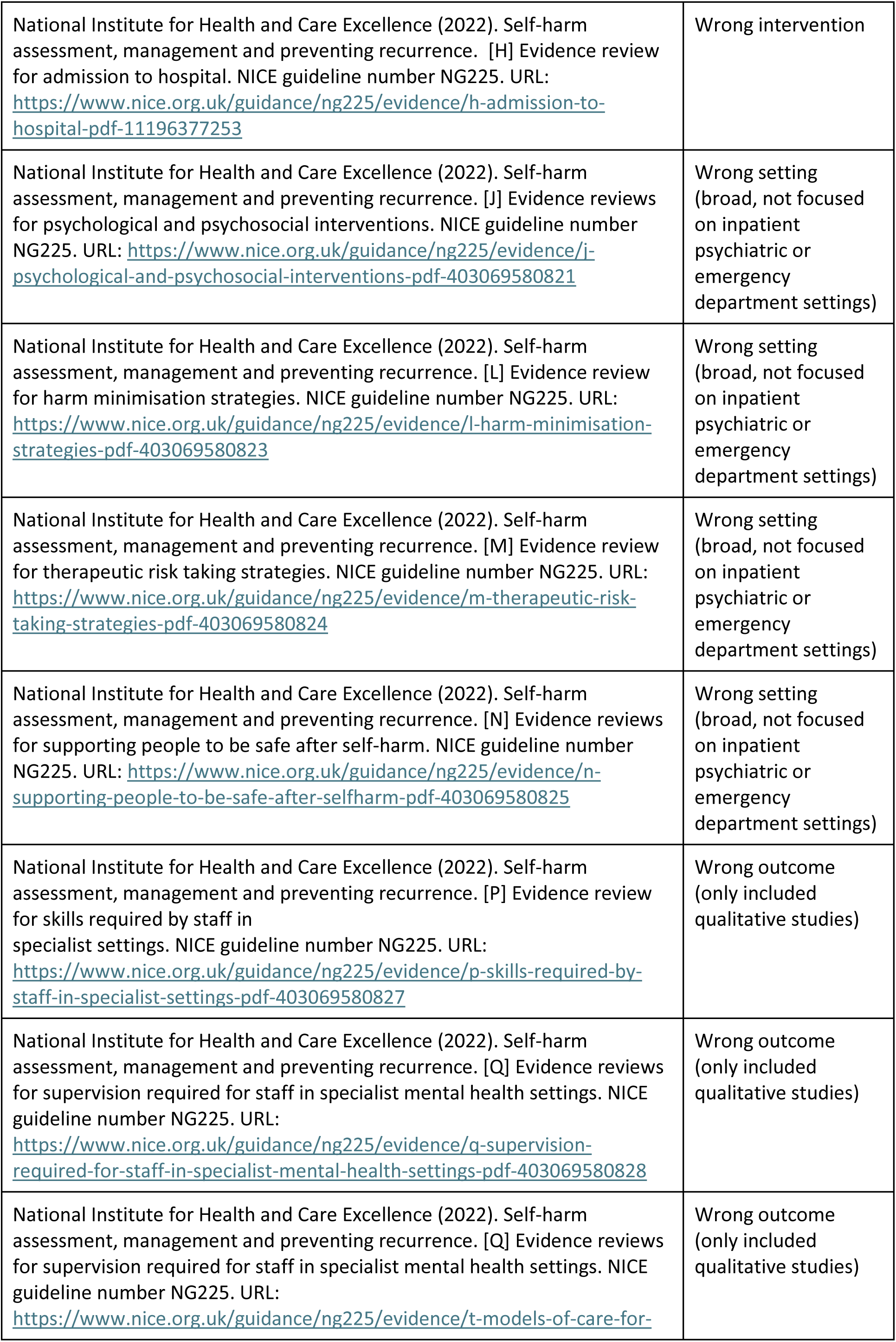
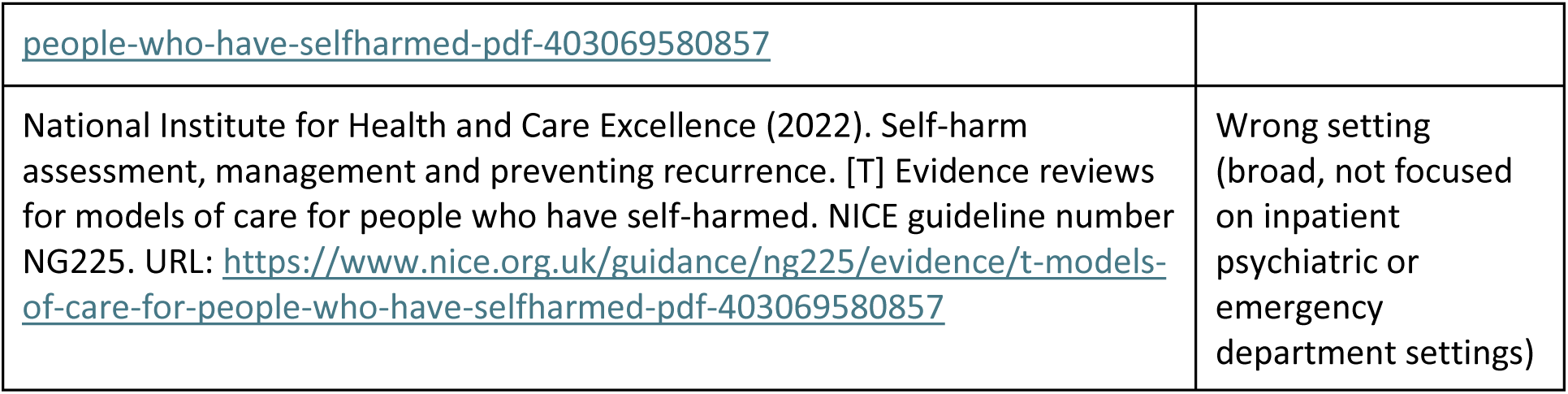

## Appendix G: Table of review characteristics

| Author, date, title, review type | Review aim | Setting (inpatient/emergency department) | Review scope | Relational interventions identified | Review conclusions |
| --- | --- | --- | --- | --- | --- |
| Nawaz et al. (2021) (65)<br><u>Title:</u> Interventions to reduce self-harm on in-patient wards: systematic review<br>Systematic review<br>(23 included papers) | Assess the efficacy of interventions that may be used to reduce the incidence and severity of self-harm and suicide attempts in adolescent and adult psychiatric inpatient settings. | Inpatient | <u>Searched:</u> Academic databases only<br><u>Designs:</u> Any study with a quantitative component. Excluded qualitative studies, commentaries and reviews.<br><u>Populations:</u> Included inpatients of all ages. Excluded people with intellectual disabilities.<br><u>Settings:</u> Included all mental health ward types (e.g., acute, adolescent, PICU, forensic). Excluded A&E, community settings, other general hospital settings.<br><u>Outcomes:</u> Self-harm and suicide<br><u>Interventions:</u> Interventions with any aim if impact on self-harm was a reported outcome | DBT | Identified some evidence of some interventions reducing self-harm but noted variable study quality and some interventions being poorly described. |
|  |  |  |  | Problem-solving therapy |  |
|  |  |  |  | Steps to Enhance Positivity (STEPS) therapy |  |
|  |  |  |  | Systems Training for Emotional Predictability and Problem Solving (STEPPS) therapy |  |
|  |  |  |  | Unified Protocol for the Transdiagnostic Treatment of Emotional Disorders' |  |
|  |  |  |  | Phone-based positive psychology |  |
|  |  |  |  | Post-admission cognitive therapy |  |
|  |  |  |  | Safewards |  |
|  |  |  |  | City nurses |  |
|  |  |  |  | Collaborative problem-solving training for nurses |  |
|  |  |  |  | Twilight nursing shift and structured evening activities programme |  |
|  |  |  |  | Bradford Refocusing model |  |
| Yiu et al. (2021) (51) | To examine the | Inpatient | <u>Searched:</u> Academic databases and ISRCTN | CBT | The results of this meta- |
| <u>Title:</u> A systematic review and meta-analysis of psychosocial interventions aiming to reduce risks of suicide and self-harm in psychiatric inpatients<br>Systematic review and meta-analysis<br>(10 included papers) | effectiveness of psychosocial interventions for suicide or self-harm in acute mental health inpatient settings on suicidality, self-harm (primary outcomes), depression, hopelessness, and suicide attempts (secondary outcomes). |  | Registry (trial registry)<br><u>Designs:</u> Only included RCTs<br><u>Population:</u> Included adult inpatients<br><u>Settings:</u> Inpatient mental health settings<br><u>Outcomes:</u> Self-harm and suicide were primary outcomes.<br><u>Interventions:</u> Included psychosocial interventions (non-pharmacological intervention targeting psychological or social factors that can reduce self-harm and suicide in people with mental health problems) | DBT | analysis suggest that psychosocial interventions are not any more effective in reducing suicidality or suicide attempts post-therapy and at follow-up in acute mental health inpatient settings than control interventions. However, large-scale RCTs are needed to confirm. No RCTs were identified for self-harm interventions, highlighting a need for more research. |
|  |  |  |  | Peer support and DBT strategies |  |
| Griffiths et al. (2022) (66)<br><u>Title:</u> Non-restrictive interventions to reduce self-harm amongst children in mental health inpatient settings: Systematic review and narrative synthesis<br>Systematic review<br>(7 included papers) | To identify interventions to reduce self-harm amongst children in mental health inpatient settings that do not rely on using restrictive practices, and evidence of their effectiveness. | Inpatient | <u>Searched:</u> Academic databases only<br><u>Designs:</u> Included quantitative, qualitative and mixed methods primary research. Excluded reviews, case studies, single case designs, conference papers, unpublished theses.<br><u>Population:</u> Included CYP inpatients. Excluded studies where >50% of the population were over 18 years old.<br><u>Settings:</u> Included CYP inpatient mental health settings.<br><u>Outcomes:</u> Self-harm<br><u>Interventions:</u> Non-restrictive interventions designed to reduce self-harm | DBT-informed interventions | Few studies found, and all were quantitative. The interventions identified comprised multiple elements making it difficult to discern which were essential to their effectiveness. The underlying theoretical assumptions and mechanisms of change were also unclear. Future research should involve young people with lived experience, use standard outcome measures, and investigate acceptability for children and young people as well as effectiveness. |
|  |  |  |  | Nursing twilight shift and evening activities programme |  |
|  |  |  |  | Staff training in DBT and seclusion and restraint, programme to reward patient behaviour, 5 patient exercise sessions per week |  |
| Thibaut et al. (2019) (52)<br><u>Title:</u> Patient safety in inpatient mental health settings: a systematic review<br>Systematic review<br>(364 included studies) | Identify and synthesise the literature on patient safety within inpatient mental health settings. | Inpatient | <u>Searched:</u> Academic databases, Google Scholar<br><u>Designs:</u> Empirical peer-reviewed studies with a clear aim or research question, that used primary data, written in English, published between 1<sup>st</sup> Jan 1999 to 27<sup>th</sup> June 2019. Excluded secondary data, protocols, editorials, commentaries/clinical case reviews/'snapshot' studies of a patient group, book chapters, conference abstracts, audits, dissertations, epidemiological studies and reviews. No restrictions on comparators.<br><u>Population:</u> Included mental health inpatients. Excluded centres on physical healthcare patients.<br><u>Settings:</u> Inpatient settings. Excluded amalgamation of data from inpatient and outpatient settings (where inpatient sample cannot be separated out), primary care, outpatient mental health services, community or social care<br><u>Outcomes:</u> Patient safety outcomes. Excluded studies where patient safety was not the central research question or outcome.<br><u>Interventions:</u> Excluded interventions where patient safety was not the central aim | DBT informed skills training for self-harm – 'Living through distress' | Highlighted a lack of research into interventions for managing risk to self in inpatient settings, particularly a lack of RCTs. |
|  |  |  |  | Peer support and DBT strategies |  |
|  |  |  |  | Special observations |  |
|  |  |  |  | Collaborative Management and Assessment of Suicide |  |
| Evans et al. (2022) (58) | Identify the barriers and facilitators to | Inpatient | <u>Searched:</u> Academic databases for English language citations between 2009-2019 and | Special observations | Evidence suggests that monitoring and checking the |
|  |  |  |  | No-suicide contracts |  |
| <u>Title:</u> Managing suicidality in inpatient care: a rapid review. The Journal of Mental Health Training, Education and Practice<br>Rapid review | implementing relational and environmental risk management approaches that address suicidality in inpatient mental health and learning disability services. |  | Google searching to identify relevant policy and guideline documents.<br><u>Designs:</u> Included quantitative and qualitative research, and policies, guidance and reports<br><u>Population:</u> Inpatients in mental health and learning disability services<br><u>Settings:</u> Inpatient mental health and learning disability services<br><u>Outcomes:</u> Suicidality<br><u>Interventions:</u> Relational and environmental risk management approaches that address suicidality | Tidal model | environment regularly is important, along with closer engagement and observation of patients to an agreed protocol by informed nursing staff. Standardisation, staff training, and individual patient risk formulations are important. Increased engagement at admission, and when reducing observation levels, is important. Most research focused on locking wards, observation levels and care planning specifically for going on leave from the ward. There is a need for more research investigating 'engagement activities', exploring what they are and their effectiveness. There is a need for new innovative ways of managing suicide risk in inpatient settings combining meaningful engagement and patient safety. |
| Bloom et al. (2012) (67)<br><u>Title:</u> Use of Dialectical Behavior Therapy in Inpatient Treatment of Borderline Personality Disorder: A Systematic Review<br>Systematic review<br>(11 included papers) | To characterise different modifications of standard DBT that have been delivered in inpatient settings and to report on the effectiveness of the DBT treatment strategies implemented in such settings to reduce target symptoms associated with the disorder. | Inpatient | <u>Searched:</u> Academic databases only<br><u>Designs:</u> Included published, peer-reviewed empirical studies.<br><u>Population:</u> Patients with a diagnosis of BPD or self-reported recent suicidal or out-of-control behaviours.<br><u>Settings:</u> Inpatient settings<br><u>Outcomes:</u> Looked at a range<br><u>Interventions:</u> Any form of DBT. Treatment had to aim to address BPD symptoms (including but not limited to self-harm, suicidal behaviour or overtly aggressive behaviour) as well as other psychiatric symptoms (e.g., symptoms of depression and anxiety). Excluded DBT addressing symptoms not related to BPD, DBT not adapted from Linehan's published DBT text, or not administered in an inpatient mental health setting. | DBT | There is considerable variation in the configuration and duration of DBT implementation for inpatients with borderline personality disorder. However, findings suggest that DBT may be effective in reducing symptoms related to borderline personality disorder in inpatient settings. Future research should standardise and systematically test inpatient DBT. |
| Timberlake et al. (2020) (77)<br><u>Title:</u> Nonsuicidal Self-Injury: Management on the Inpatient | To review the latest research on treatment and management of non-suicidal self-injury specific for the acute inpatient psychiatric population. | Inpatient | <u>Searched:</u> Academic databases only<br><u>Designs:</u> Included peer-reviewed articles. Excluded abstract only/poster presentations<br><u>Population:</u> Adolescent, young adult and adult populations. Excluded studies only focusing on | Special observation<br>Safety contracts<br>Combined DBT and mentalisation-based group therapies<br>Safewards | Concludes that there is a lack of empirical research investigating interventions for reducing self-harm in inpatient settings. States that research studies with controlled designs are |
| Psychiatric Unit<br>Non-systematic review<br>(9 included papers) |  |  | CYP or older adult populations, developmentally delayed populations, psychotic disorders and traumatic brain injury populations.<br><u>Settings:</u> Included inpatient settings.<br><u>Outcomes:</u> Deliberate self-harm. Excluded studies not focusing on self-harm or that did not distinguish between non-suicidal self-harm or suicidal acts.<br><u>Interventions:</u> Any | Collaborative problem-solving nursing approach | needed. |
| Cox et al. (2010) (76) | To critically review the empirical evidence base for alternative | Inpatient | <u>Searched:</u> Academic databases only<br><u>Designs:</u> Included empirical papers. Excluded | Bradford Refocusing model<br>City nurses | This review emphasised how enhanced observations and the development of |
| <u>Title:</u> Alternative approaches to 'enhanced observations' in acute inpatient mental health care: a review of the literature<br>Non-systematic review<br>(5 included papers) | approaches to 'enhanced observations' from those proposed in the Standing Nursing and Midwifery Advisory Committee guidelines (SNMAC DoH 1999) on individuals receiving care on open acute inpatient mental health wards. |  | non-empirical papers.<br><u>Populations:</u> Not specified<br><u>Settings:</u> Included acute inpatient mental health settings. Excluded prisons, forensic mental health settings, or any other permanently locked inpatient mental healthcare setting.<br><u>Outcomes:</u> Not specified<br><u>Interventions:</u> Alternative approaches to 'observations', structured programmes of change to nurses' beliefs, attitudes and practice or changes to policy or changes in therapeutic functions of the ward environment with direct relevance to managing individuals at risk and reducing 'observations' | Special observations | alternative approaches are a complex activity requiring planning and consideration. It highlights a lack of empirical evidence for alternatives, and a need to revise current practices due to dissatisfaction amongst both patients and staff. |
| James et al. (2012) (75) | To examine the | Inpatient | <u>Searched:</u> Academic databases only | Special observations | Wards attempted to manage |
| <u>Title:</u> Self-harm and attempted suicide within inpatient psychiatric services: a review of the literature<br>Non-systematic review<br>(88 included studies) | prevalence, characteristics, and antecedents of self-harm incidents on psychiatric wards, the measures used by wards to manage self-harm, and the experiences of psychiatric nurses. |  | <u>Designs:</u> Included empirical studies of self-harm and attempted suicide in adult psychiatric inpatient services published in English between 1960-2010.<br><u>Population:</u> Included adults, older adults, adolescents and CYP. Excluded people with a history of self-harm who did not self-harm during their inpatient stay.<br><u>Settings:</u> Included a range of inpatient mental health services (e.g., acute, forensic, PICU, rehabilitation wards). Excluded studies conducted in older adult, adolescent or CYP mental health services.<br><u>Outcomes:</u> Self-harm and attempted suicide<br><u>Interventions:</u> NA | Zero suicide contracts | self-harm using a wide range of interventions. Although there is some evidence to suggest that intermittent observation is effective in reducing rates of self-harm and attempted suicide (Bowers et al. 2008, 2011), on the whole, there has been very little research into the effectiveness of these self-harm management strategies. |
| Helleman et al. (2014) (68)<br><u>Title:</u> Evidence base and components of Brief Admission as an intervention for patients with borderline personality disorder: a review of the literature<br>Systematic review | To identify the key components of Brief Admission as a crisis intervention for patients with a BPD and the evidence base for the components of Brief Admission. | Inpatient | <u>Searched:</u> Academic databases<br><u>Designs:</u> Included quantitative studies, qualitative studies, reviews and practice reports. Excluded articles published before 1985.<br><u>Populations:</u> Patients with a BPD diagnosis<br><u>Settings:</u> Any inpatient setting where brief admission was described as being used<br><u>Outcomes:</u> Any | Green cards<br>Brief Admission crisis intervention program | Only two of the included studies examined self-harm or suicide as outcomes. The review authors concluded that Brief Admission can be effectively used to reduce self-harm and suicide in patients with a diagnosis of BPD. |
| (10 included papers) |  |  | <u>Interventions:</u> Brief admissions for people with BPD. Excluded articles that did not describe the components of Brief Admission. |  |  |
| McCabe et al. (2018) (69)<br><u>Title:</u> Effectiveness of brief psychological interventions for suicidal presentations: a systematic review<br>Systematic review<br>(4 included papers) | Systematically review the effectiveness of brief psychological interventions in addressing suicidal thoughts and behaviour in healthcare settings. | ED | <u>Searched:</u> Academic databases<br><u>Designs:</u> Included published controlled studies (cluster randomised controlled trials, randomised controlled trials, controlled before-and-after studies and controlled pre-test/post-test designs). Excluded non-controlled studies.<br><u>Population:</u> Participants of any age and gender at risk of suicide. Excluded assisted suicide and self-harm without intent to die.<br><u>Settings:</u> Any healthcare setting (all results were from emergency departments)<br><u>Outcomes:</u> Primary outcome was suicidal ideation, using any measure. Other outcomes included: identification of suicide risk, suicide attempts, suicide, hope, patient distress and depression.<br><u>Intervention:</u> Involve interactions between professionals/ paraprofessionals (e.g., lay mental health workers, nursing assistants, educators, volunteers) and patients addressing suicidal thoughts and plans. Two-way communication (i.e. not one-way communication in the form of letters/postcards/text messages or exclusively self-guided questionnaires/instruments) between at least one professional/ | Brief intervention and contact<br>The Attempted Suicide Short Intervention Program<br>Teen Options for Change<br>Safety Assessment and Follow-up Telephone Intervention<br>Crisis intervention program | Although there are relatively few studies to-date, brief psychological interventions appear to be effective in reducing suicide and suicide attempts. It is unclear to what extent the effect is due to specific psychological components or to more frequent contacts. All studies to-date have been conducted in the ED. They involve providing information, developing understanding about the suicidal crisis, developing safety plans for potential future crises, and follow-up contact to monitor and support patients. The interventions tested did not appear to reduce suicidal ideation. This suggests that patients may still experience significant distress, and that the interventions affect behaviours (i.e. suicides, suicide attempts). Engaging patients early on and providing brief therapeutic interventions grounded in psychological theories of suicidal behaviour, combined |
|  |  |  | paraprofessional and one patient (other people can be present). The focus must be on suicidal thoughts and plans rather than diagnostic conditions e.g. depression, anxiety, BPD. Focus on routine clinical encounters. Brief interventions (defined as up to three sessions delivered in/soon after presenting episode) which can be supplemented by further follow-up contact. |  | with ongoing follow-up, could be beneficial. |
| Newton et al. (2010) (53)<br><u>Title:</u> Pediatric suicide-related presentations: a systematic review of mental health care in the emergency department<br>Systematic review<br>(10 included studies) | Evaluate the effectiveness of interventions for paediatric patients with suicide-related emergency department visits. | ED | <u>Searched:</u> Academic databases, clinicaltrials.gov and contacted authors for unpublished research<br><u>Designs:</u> Included experimental and quasi-experimental studies. No restrictions placed on comparison groups.<br><u>Population:</u> CYP (aged < 18 years) or only partially including this age range, or parents or emergency department staff<br><u>Settings:</u> Interventions initiated in the emergency department or immediately after<br><u>Outcomes:</u> At least one clinically relevant primary outcome needed. Could be health-related (rates of self-injurious behaviour, death by suicide, suicidal ideation), parent-related (reporting of means restriction) or care-related (service-delivery, consultation, documentation)<br><u>Interventions:</u> Mental-health based, suicide-prevention focused intervention initiated in the emergency department or immediately after emergency department discharge<br>through direct referral/enrolment | Interventions started after discharge from the ED<br>Interventions starting in the ED and continuing post-ED | Robust research into psychosocial management of paediatric suicide-related ED presentations is emerging but limited. Based on the available evidence, transition interventions that start in the ED and continue post-ED appear most promising for reducing suicide-related outcomes and improving post-ED treatment adherence. Findings should be interpreted with caution, however, given the limitations of the included studies. There is a lack of consistency in methods and outcomes measured, and overall limited quality of the studies available. |
| Virk et al. (2022) (73)<br><u>Title:</u> To synthesise evidence on interventions that can be implemented in the paediatric emergency department for children and adolescents presenting with suicidal ideation.<br>Rapid review<br>(6 included papers) | To synthesise evidence on interventions that can be implemented in the paediatric emergency department for children and adolescents presenting with suicidal ideation. | ED | <u>Searched:</u> Academic databases<br><u>Designs:</u> Included RCTs with any comparator published after January 2010. Excluded non-randomised controlled trials.<br><u>Population:</u> CYP aged 6-19 years old. At least 25% needed to be recruited from a paediatric emergency department.<br><u>Settings:</u> Paediatric emergency departments.<br><u>Outcomes:</u> Suicidal ideation, engagement with outpatient services, incidence of depressive symptoms, hopelessness, family empowerment, hospital admission and feasibility of interventions.<br><u>Interventions:</u> Any psychological/ psychosocial/ non-pharmacological intervention used with children or young people in the paediatric emergency department. Excluded interventions employed outside the clinical setting. | Family-based interventions | Overall, findings suggest that family-based interventions for young people presenting with suicidal ideation to EDs are associated with a reduction in suicidal ideation, whereas evidence for the benefit of motivational interviewing is more mixed. Overall, there is a lack of high-quality evidence, and therefore conclusions should be drawn with caution. Further high-quality RCTs with larger sample sizes are needed, which could consider relevant subpopulations e.g. looked after children, young people at very high risk of suicide, young people with experiences of abuse or domestic violence. |
|  |  |  |  | Motivational interviewing |  |
| Broadway-Horner et al. (2022) (59)<br><u>Title:</u> Psychological therapies and non- | To recognize and assess the results from all studies including randomized control trials (RCTs) that have | ED | <u>Searched:</u> Academic databases and Google Scholar<br><u>Designs:</u> Only included RCTs. Excluded studies included in Hawton et al. (1998). | Manual-assisted cognitive therapy (MACT) | At present, evidence is lacking to indicate the most effective forms of treatment for non-suicidal self-injury overdosing in LGBTIQ and non-binary |
|  |  |  |  | Brief psychodynamic interpersonal therapy |  |
| suicidal self-injury in LGBTIQ in accident and emergency departments in the UK: a scoping review<br>Scoping review<br>(7 included papers) | studied the efficiency of psychiatric and psychological assessment of people who have depression that undergo non-suicidal self-injury (NSSI) by self-poisoning, presenting to UK Accident and Emergency Departments. |  | <u>Population:</u> LGBTIQ and non-binary study participants aged 18 years and over who have engaged in non-suicidal self-injury by overdose shortly before entry to the study.<br><u>Settings:</u> Included A&E departments in the UK. Excluded studies with no A&E involvement.<br><u>Outcomes:</u> Included repetition of self-harm behaviour. Excluded studies focusing on suicide.<br><u>Interventions:</u> Psychiatric and psychological therapy treatments | Crisis cards | populations. It appears that psychodynamic interpersonal therapy is one of the most promising approaches for reducing repeated self-harm by overdosing after ED presentation. There was no evidence for manual-assisted cognitive therapy having a significant impact on repeat self-harm. |
| Luxton et al. (2013) (78)<br><u>Title:</u> Can post discharge follow-up contacts prevent suicide and suicidal behavior? A review of the evidence<br>Non-systematic review<br>(11 included papers) | Evaluate the evidence for the effectiveness of suicide prevention interventions that involve follow-up contacts with patients | Inpatient and ED | <u>Searched:</u> Academic databases<br><u>Designs:</u> Included published articles<br><u>Populations:</u> Included inpatient psychiatric patients or emergency room patients being discharged home<br><u>Settings:</u> Inpatient mental health services or emergency departments<br><u>Outcomes:</u> Had to include measurement of suicidal behaviours (suicide, suicide attempts or suicidal ideation).<br><u>Interventions:</u> Follow-up interventions with at least one form of follow-up contact with patients (e.g. letters, postcards, phone calls, in-person visits, electronic contact). The contacts had to be pre-planned, systematic, directed specifically to the patient and<br>initiated by the care providers, but not part of a larger psychotherapy or pharmacotherapy intervention. | Caring letters<br>Postcard follow-up contacts<br>Telephone follow-up contacts | In general, repeated follow-up contacts may help prevent suicidal behaviours. 3/11 studies showed a statistically significant reduction in repeat suicide attempts, and two studies showed a reduction in deaths by suicide. Four studies reported mixed or on-conclusive results but did show trends towards preventative effects. Two studies did not show preventative effects for follow-up interventions. Overall, the evidence is a bit mixed but indicates that this type of post-discharge interventions can be effective in reducing suicide deaths, attempts and ideation. |
| Huber et al. (2023) (54)<br><u>Title:</u> The effectiveness of brief non-pharmacological interventions in emergency departments and psychiatric inpatient units for people in crisis: A systematic review and narrative synthesis<br>Systematic review<br>(39 included studies) | To create a taxonomy of brief non-pharmacological interventions, and review their evaluation methods and effectiveness | Inpatient and ED | <u>Searched:</u> Academic databases, and government health websites for references, plus key non-government organisation crisis resources<br><u>Designs:</u> Included RCTs, non-RCTs, cohort and case-control studies, case series and case reports, surveys and qualitative studies were included. Excluded all evidence syntheses, expert opinion and descriptive studies<br><u>Populations:</u> Included people in crisis presenting to emergency departments with any complaint related to mental or behavioural health, or an inpatient on a psychiatric ward experiencing self-harm thoughts/behaviours or agitation/aggression. Excluded people with solely drug and/or alcohol presentations.<br><u>Settings:</u> Emergency departments and psychiatric wards (included treatments initiated in emergency departments and continued in inpatient settings). Excluded interventions started in the emergency | Special observation | The evidence base for many non-pharmacological interventions in EDs and inpatient services for people in crisis is severely limited, if not entirely absent. Even when there is evidence, inconsistency in outcomes across studies makes it hard to draw conclusions, though some interventions show promise. |
|  |  |  |  | No suicide contracts/safety plans |  |
|  |  |  |  | Short admissions |  |
|  |  |  |  | Specialised suicide-specific therapies in the ED (including post-admission cognitive therapy, Successful Negotiation Acting Positively therapy, family-based crisis intervention) |  |
|  |  |  | department and continued in outpatient settings, and interventions in general medical wards, aged care facilities, group homes, jails, and other non-hospital settings).<br><u>Outcomes:</u> Any<br><u>Interventions:</u> Included all primarily brief non-pharmacological interventions aimed at addressing psychiatric complaints. Incidental medication use was not an exclusion criterion. Interventions that were used during crisis admissions, even if they were not used on a crisis unit, were included. Only included clinical interventions, not processes of care pathways. Excluded interventions if medications were identified as a component of the intervention, and interventions lasting longer than a week. |  |  |
| Mullen et al. (2022)<br>(74)<br><u>Title:</u> Safewards: An integrative review of the literature within inpatient and forensic mental health units<br>Integrative review<br>(19 included studies) | Synthesize the current knowledge and understanding about the implementation, effectiveness, acceptability of Safewards and how it meets the needs of consumers within inpatient and forensic mental health units. | Inpatient | <u>Searched:</u> Academic databases<br><u>Designs:</u> Included all peer-reviewed articles<br><u>Populations:</u> Inpatients in mental health settings<br><u>Settings:</u> Mental health inpatient settings (forensic and non-forensic)<br><u>Outcomes:</u> implementation outcomes (including staff acceptability), effectiveness outcomes (conflict and containment), consumer experiences of care<br><u>Interventions:</u> Safewards | Safewards | Safewards can be effective in reducing conflict within forensic and non-forensic inpatient mental health settings. The review also revealed limitation of fidelity measures and the importance of staff involvement in implementation. There is a lack of consumer perspectives on the Safewards model in the literature. |
|  |  |  | <u>Comparators:</u> Stated 'not applicable' |  |  |
| Ward-Stockham et al. (2022) (55)<br><u>Title:</u> Effect of Safewards on reducing conflict and containment and the experiences of staff and consumers: A mixed-methods systematic review<br>Systematic review<br>(14 included studies) | To evaluate the effect of Safewards on conflict and containment events in inpatient units and the perceptions of staff and consumers | Inpatient | <u>Searched:</u> Academic databases, and unpublished and grey literature repositories<br><u>Designs:</u> Quantitative, qualitative or mixed methods studies<br><u>Populations:</u> Healthcare staff and inpatient consumers<br><u>Settings:</u> Any inpatient setting globally<br><u>Outcomes:</u> Rates of conflict, rates of containment, or staff or consumer experience of safety or perspectives of Safewards<br><u>Interventions:</u> Safewards<br><u>Comparators:</u> No restrictions stated | Safewards | Evidence suggests that Safewards mitigates multiple types of conflict at various stages and can prevent triggers that lead to containment. Most staff and consumers reported Safewards improved therapeutic relationships through enhanced communication and promote a sense of calm and safety on inpatient wards. Staff and patients reported improved ward atmosphere, leading to consumer-centred, recovery-oriented care. Safewards improved the experience of safety from the perspective of staff and patients when combined with ongoing training, leadership and time for consolidation. While results are promising, they should be used cautiously until more robust evidence is established. |
| Finch et al. (2021) (56)<br><u>Title:</u> A Systematic Review of the Effectiveness of Safewards: Has Enthusiasm Exceeded Evidence?<br>Systematic review<br>(13 included studies) | Examine whether Safewards is effective in reducing conflict and containment events; and improving ward climate. | Inpatient | <u>Searched:</u> Academic databases, grey literature (including dissertation, conference and white papers) using university search engines and dissertation repositories, Google Scholar<br><u>Designs:</u> Included journal-published quantitative, qualitative and mixed methods studies written in English<br><u>Populations:</u> Not stated<br><u>Settings:</u> Inpatient settings<br><u>Outcomes:</u> Conflict, containment, ward climate. Excluded studies looking at other factors (e.g., staff experiences of training or challenges with implementation)<br><u>Intervention:</u> Safewards<br><u>Comparators:</u> No restrictions stated | Safewards | There is evidence suggesting that Safewards is effective for reducing conflict and containment in general mental health services, but there is insufficient high-quality empirical evidence to support its effectiveness in other settings. Further research using robust methodological designs with larger, more representative samples is required to determine whether Safewards is effective across the range of contexts in which it is currently being applied. |
| NICE; Self-harm: assessment, management and preventing recurrence; [F] Evidence review for assessment in specialist settings (70)<br>Systematic review<br>(4 included studies) | Explore how assessment for people who have self-harmed should be undertaken in specialist settings? | Mixed (specialist MH settings including inpatient, A&E, and community services) | <u>Searched:</u> Academic databases<br><u>Designs:</u> Included systematic reviews of RCTs or non-randomised comparative prospective and retrospective cohort studies; RCTs; non-randomised comparative prospective cohort studies with N≥100 per treatment arm; non-randomised comparative retrospective cohort studies with N≥100 per treatment arm. Excluded conference abstracts<br><u>Populations:</u> Included all people who have self-harmed, including those with a mental health problem, neurodevelopmental disorder or a learning disability, who have presented to<br>specialist mental health services. Excluded people displaying repetitive stereotypical self-injurious behaviour, for example head-banging in people with a significant learning disability<br><u>Settings:</u> Included specialist mental health settings such as community mental health services, A&E (by specialist staff), inpatient mental health settings. Excluded non-specialist settings.<br><u>Outcomes:</u> Critical outcomes: self-harm repetition (for example, self-poisoning or self-cutting); service user satisfaction (dignity, compassion and respect); suicide. Important outcomes: quality of life; initiation of safeguarding procedures; distress; engagement with after-care<br><u>Interventions:</u> Included assessment including principles of active listening; therapeutic assessment; comprehensive biopsychosocial assessment; assessment performed by different professions [e.g., psychiatric nurses], culturally sensitive assessment.<br><u>Comparators:</u> assessment not including principles of active listening, triage assessment, assessment performed by different professions [such as doctors]; uniform assessment (that is, not taking culture into account). | Therapeutic assessment | Few studies were identified comparing different models of self-harm assessment in specialist mental health settings for people who have self-harmed. No studies reported on suicide as an outcome. In summary, no evidence for impact of therapeutic assessment or psychiatrist vs psychiatric nurse assessment in A&E on repeat self-harm or suicide. |
| Austin et al. (2024) (71) | Synthesise the research evidence | ED | <u>Searched:</u> Academic databases | Assertive case management | This review identified strategies to enhance ED care |
| <u>Title:</u> Improving emergency department care for adults presenting with mental illness: a systematic review of strategies and their impact on outcomes, experience, and performance<br>Systematic review<br>(46 included studies) | associated with strategies used to improve ED care delivery outcomes, experience, and performance for adults presenting with mental illness. |  | <u>Designs:</u> Included empirical peer-reviewed research articles. Excluded literature reviews, conference posters or abstracts, grey literature and case reports. Only included articles published in English.<br><u>Population:</u> Adult mental health presentations (e.g., undifferentiated, suicidal, deliberate self-harm, scheduled, substance-related and addictive disorders, depressive and anxiety disorders). Excluded studies involving people aged under 18, or focused on disability or neurodiversity (e.g., autism).<br><u>Setting:</u> Included EDs. Excluded interventions conducted primarily in the pre-hospital, post-hospital or a ward/clinic setting other than the ED.<br><u>Outcomes:</u> Included measures of system performance (e.g., waiting time, length of stay, time to treatment/assessment, admissions, referrals), patient outcomes (e.g., readmission, adverse events, medical errors, missing diagnoses, pain, quality of life), patient experience, or staff experience.<br><u>Intervention:</u> Implemented models of care or system changes. Excluded studies that did not report an intervention, or that screened presentations without intervention in the ED.<br><u>Comparators:</u> Usual care or other form of care. |  | for individuals presenting with mental health difficulties, including self-harm and suicidality. This included assertive case management. However, the heterogeneity in study samples, strategies, and outcomes evaluated makes adopting existing strategies challenging. Effective strategies for providing care in the ED must align ED system goals, patient needs, and staff experiences. |
| Chammas et al. (2022) | Provide an overview of | Inpatient | <u>Searched:</u> One academic database (PubMed) | Anti-suicide contracts | They identified some |
| (79)<br><u>Title:</u> Inpatient suicide in psychiatric settings: Evaluation of current prevention measures<br>Non-systematic review<br>Number of included studies not stated. | the progress that has been made in the field of inpatient suicide prevention in recent years, discuss the problems that remain, and the future potential developments. |  | <u>Designs:</u> No inclusion or exclusion criteria stated.<br><u>Populations:</u> Inpatient mental health populations. No restrictions specified.<br><u>Settings:</u> Inpatient mental health services<br><u>Outcomes:</u> Suicide-related outcomes<br><u>Interventions:</u> Suicide prevention measures in inpatient mental health services<br><u>Comparators:</u> Not stated | Collaborative Assessment and Management of Suicidality (CAMS)<br>Dialectical behaviour therapy (DBT) | approaches with some evidence (e.g., CAMS, some psychotherapeutic approaches), and others which are now outdated (e.g., anti-suicide contracts). |
| Chaudhary et al. (2020) (72)<br><u>Title:</u> Suicide during Transition of Care: a Review of Targeted Interventions<br>Systematic review<br>(40 included studies) | Summarise the evidence for interventions providing care during the first few weeks after discharge from a healthcare facility (when risk of suicide is highest). | Inpatient and ED | <u>Searched:</u> Academic databases<br><u>Designs:</u> Included all original studies, including RCTs and non-randomised trials. They excluded case reports, case series, letters to editors, study protocols, theses, reviews, commentaries, conference papers, abstract-only articles, book chapters and news articles.<br><u>Population:</u> People discharged from a medical facility to the community. No restriction on race, place, sex, age, ethnicity.<br><u>Setting:</u> Not stated.<br><u>Intervention:</u> Interventions targeting suicidal behaviours after discharge from a medical facility.<br><u>Outcomes:</u> Suicide-related outcomes | Green cards<br>Caring letters<br>Postcards<br>Letter and telephone contact<br>Telephone contacts<br>Brief Intervention and Contact (BIC)<br>Family Intervention for Suicide Prevention (FISP)<br>Mobile crisis team intervention | This review highlights that patients are at high risk of suicide when transitioning from medical care facilities to the community. The evidence they identified for outcomes related to suicidal ideation, attempts and self-harm was mixed. Only 4/9 studies indicated improvements in suicidal ideation, while minimal evidence supported a favourable impact on self-harm, with just three studies showing positive results. They note methodological issues, such as missing data, baseline severity differences between groups, and the lack of a reliable definition of self-harm, which complicate these<br>findings. Additionally, the response to interventions differed between individuals presenting with suicidal behaviours for the first time and those with repeat self-harm. They stated that patients with repeat self-harm are more likely to benefit from psychosocial interventions such as DBT or CBT. |
| De Santis et al. (2015) (64)<br><u>Title:</u> Suicide-specific Safety in the Inpatient Psychiatric Unit<br>Non-systematic review<br>Number of included studies not stated | Assist psychiatric mental health nurses in advance practice, education, leadership and administration, to review and update training, policies, and procedures specific to suicide prevention in inpatient units | Inpatient | Search strategy and eligibility criteria not stated. Focus was on suicide-related outcomes. | No-suicide contracts | Preventing self-harm is most effective by enhancing services, reducing access to lethal means, connecting and collaborating with patients, utilising best practices, reducing acute symptoms, promoting healthy coping and problem-solving skills, increasing hopefulness, identifying reasons for living, strengthening interpersonal connections, and treating patients with compassion and respect. There is a gap in the evidence regarding the effectiveness of psychotherapeutic interventions, multicomponent interventions, observation and monitoring, and effectiveness of hospitalisation in reducing suicidality. It is important for nurses to be trained in |
|  |  |  |  | Collaborative Assessment and Management of Suicidality (CAMS) |  |
|  |  |  |  |  | intensive, competency-based, best practice curricula. |
| Falcone et al. (2017) (61)<br><u>Title:</u> Taking care of suicidal patients with new technologies and reaching-out means in the post-discharge period<br>Non-systematic review<br>Number of included studies not stated | To understand the role of new technologies for reducing self-harm, suicide attempt, and death by suicide, while paying particular attention to post-discharge from an ED or psychiatric ward. | Inpatient and ED | <u>Searched:</u> Academic databases and ResearchGate<br><u>Designs:</u> Papers in English between 1977-2016.<br><u>Population:</u> Patients discharged from inpatient psychiatric wards or from an ED<br><u>Setting:</u> Psychiatric wards or EDs<br><u>Intervention:</u> New technologies (e.g., postcards/letters, text messages, crisis cards, telephone contacts, online interventions) in suicide prevention<br><u>Outcomes:</u> Self-harm and suicide attempts post-discharge | Caring letters | Concluded that it is necessary to reach out and contact patients who have attempted suicide after discharge from hospital. New technologies (e.g., letters, green cards, telephone calls, postcards) can be helpful additions to standard care, helping to prevent suicides. Patients experience them as useable, effective, efficient and secure. However, their effects need to be further investigated through well-designed clinical trials. |
|  |  |  |  | Postcards |  |
|  |  |  |  | Telephone contacts |  |
|  |  |  |  | Letters and telephone contacts |  |
|  |  |  |  | Telephone contacts |  |
|  |  |  |  | Brief intervention and contact |  |
| Manna (2010) (62)<br><u>Title:</u> Effectiveness of formal observation in inpatient psychiatry in preventing adverse outcomes: the state of the science | To determine whether research supports the use of formal observation as an effective strategy in preventing potential harm to patients or others; identify any therapeutic benefit; | Inpatient | <u>Searched:</u> Academic databases, American Psychiatric Association, American Psychiatric Nurses Association.<br><u>Designs:</u> No limits on study design stated. Included quantitative, qualitative and mixed methods literature. Included reviews.<br><u>Population:</u> People in psychiatric inpatient | Bradford Refocusing Model | Findings indicate that there are still large gaps in the research regarding observations in inpatient settings. There is a lack of consensus about formal observation definitions and how it should be carried out regardless of effectiveness. |
| Non-systematic review<br>(10 included studies) | and identify gaps in the research. |  | settings<br><u>Setting:</u> Psychiatric inpatient services<br><u>Intervention:</u> Observation in a psychiatric inpatient setting<br><u>Outcomes:</u> Indications for the use of observation, impact on self-harm, suicide, violence, elopements, and it's positive and negative therapeutic merits. Nurses' and patients' perceptions on its usefulness and impact were also included. |  | This review concludes that formal observation is rooted in tradition, as little research exists evaluating its efficacy for reducing harms. |
| Navin et al. (2019) (63)<br><u>Title:</u> Suicide Prevention Strategies for General Hospital and Psychiatric Inpatients: A Narrative Review<br>Non-systematic review<br>(24 included articles) | To provide an overview of various proposed suicide prevention approaches in the general hospital, including psychiatric inpatient settings, and their evidence base. | Inpatient | <u>Searched:</u> Academic databases, Google Scholar<br><u>Designs:</u> Included peer-reviewed articles in English language journals. Excluded conference proceedings.<br><u>Population:</u> Patients in inpatient psychiatric or medical/surgical settings<br><u>Settings:</u> Inpatient psychiatric services or medical/surgical inpatient services<br><u>Interventions:</u> Suicide prevention approaches<br><u>Outcomes:</u> Suicide | Post-Admission Cognitive Therapy (PACT)<br>Collaborative Assessment and Management of Suicidality (CAMS) | There is a lack of original research on inpatient suicide prevention. There is a need for more rigorous research, and for sustainable, cost-effective, ethical and socio-culturally acceptable suicide prevention strategies. |
| Nugent et al. (2024) (60)<br><u>Title:</u> Behavioural mental health | Identify and describe evidence on brief ED-delivered behavioural and care process interventions among | ED | <u>Searched:</u> Academic databases, ClinicalTrials.gov.<br><u>Designs:</u> Included RCTs and observational studies. Limited systematic reviews to those | Cognitive abandonment psychotherapy<br>Cognitive behaviour therapy (CBT) | A wide variety of multicomponent behavioural interventions delivered in ED settings have shown short-term improvements in |
| interventions delivered in the emergency department for suicide, overdose and psychosis: a scoping review<br>Scoping review<br>(40 included studies) | patients presenting with suicide attempt or acute suicidal ideation, substance overdose, or psychosis. |  | published in the last 7 years, but no date limits on primary research.<br><u>Populations:</u> Adults presenting to EDs or urgent care centres with suicidality (attempt or acute ideation), substance overdose, or acute psychotic symptoms (where psychosis was the primary diagnosis).<br><u>Settings:</u> EDs<br><u>Interventions:</u> Included brief mental health interventions, including screening or risk assessment; triage; referral to inpatient, residential or outpatient settings; behavioural interventions; or treatment of agitation related to substance withdrawal. Excluded legal hold interventions, medication comparative effectiveness trials, primary medical interventions and cardiopulmonary stabilisation and crisis care management of use of reversal agents.<br><u>Outcomes:</u> Included studies reporting on engagement in outpatient, residential or inpatient mental healthcare, severity of acute symptoms (e.g., suicidality), ED or urgent care outcomes, patient or staff safety outcomes or adverse events or harms of interventions.<br><u>Comparators:</u> Not specified | Attempted Suicide Short Intervention Program (ASSIP) | outcomes such as reducing suicide attempts. However, the research is constrained by methodological inconsistencies, ethical challenges relating to randomisation, and setting-level barriers to implementation. Future research could investigate differences in effectiveness based on sociodemographic and clinical characteristics, setting characteristics, and intervention features. |
|  |  |  |  | Brief Intervention and Contact (BIC) |  |
|  |  |  |  | Telephone follow-up contacts |  |
|  |  |  |  | Safety Assessment and Follow-up Telephone Intervention (SAFTI) |  |
|  |  |  |  | Case management |  |
|  |  |  |  | Safety Planning Intervention+ (SPI+) |  |
| Reen et al. (2020) (57)<br><u>Title:</u> Systematic review of interventions to improve constant observation on adult inpatient psychiatric wards<br>Systematic review | To describe and categorize all interventions relevant to constant observations and integrate learning from these interventions to improve this widespread practice and to minimize its restrictive use on | Inpatient | <u>Searched:</u> Academic databases and Google Scholar<br><u>Designs:</u> Peer-reviewed studies, in English, published in any year, any country. All study designs could be included provided the other eligibility criteria are met. Studies offering recommendations on best practice of constant observation, or commentary and discussion pieces on specific interventions were excluded. | Bradford Refocusing Model | Constant observation is widely used in adult inpatient psychiatric wards to ensure patient safety. However, it can also be coercive, anti-therapeutic, and detrimental to both patients and staff. Despite its common use, clear guidelines and robust evidence for its efficacy are lacking. Further research is |
| (16 included studies) | psychiatric wards. |  | <u>Populations:</u> Adult psychiatric inpatient populations<br><u>Settings:</u> Inpatient psychiatric wards, including acute, intensive and forensic psychiatric wards. Excluded physical health settings or services other than adult inpatient psychiatric wards.<br><u>Intervention:</u> Interventions designed to impact constant observation on an inpatient psychiatric ward. Constant observation was defined as close monitoring and supervision of patients by at least one member of clinical staff either by keeping them within eyesight or at arm's length. Interventions were even included if they were designed for an inpatient psychiatric population but not actually implemented on an inpatient psychiatric ward. Excluded interventions addressing only general observation practice or intermittent observation.<br><u>Outcomes:</u> None specified<br><u>Comparators:</u> None specified | City nurses | needed to evaluate the effectiveness and potential risks of constant observation. |

A&E = Accident and Emergency; BPD = Borderline Personality Disorder; CBT = Cognitive Behaviour Therapy; CYP = Children and Young People; DBT = Dialectical Behaviour Therapy; ED = Emergency Department; LGBTIQ = Lesbian, Gay, Bisexual, Transgender, Intersex, and Queer or Questioning; NICE = National Institute for Health and Care Excellence; RCT = Randomised Controlled Trial.

