## Supplementary File 1 for "Quantitative evidence for relational care approaches to assessing and managing self-harm and suicide risk in inpatient mental health and emergency department settings: a scoping review"

**Supplementary File 1:** Impact of relational care approaches on self-harm and suicide in inpatient mental health settings

A total of 46 primary papers evaluated the impact of 30 different relational care approaches on self-harm and/or suicide-related outcomes in non-forensic inpatient mental health settings. The characteristics and results of the primary studies of relational care approaches in inpatient mental health settings are summarised narratively and presented in Table S1, below.

**DBT-based approaches**

Study characteristics

Eleven studies investigated the effectiveness of DBT-based relational care approaches in inpatient settings (81–84,137,138,141–144,149). One study had a RCT design (81), three were non-randomised controlled trials (137,142,149), four had pre-post-designs with comparison groups (82–84,144), and three had pre-post designs without a comparison group (138,141,143). Six papers focused on adult inpatient populations (81,137,138,142–144), two included both older adolescents and adults (82,141) and three included only adolescents (83,84,149). Six studies specifically recruited people with a diagnosis of borderline personality disorder (BPD) (82,137,141–144) and one recruited people with ‘personality disorder’ more generally, with a BPD sub-sample (81).

Interventions

The format of DBT-based approaches varied between studies in terms of the components involved (e.g., group skills sessions, individual therapy sessions, DBT consultation), duration and session frequency. In some studies, the intervention adhered more closely to the standard DBT model, whereas in others it was adapted more significantly. For example, Barley et al. (1993) introduced DBT components into their psychodynamic approach (82).

Effects on self-harm

All 11 primary studies included self-harm as an outcome. 9/11 reported that DBT-based approaches significantly reduced self-harm rates, including compared to controls (82,84,137,138,141–143,149,149). One pilot study reported no significant difference in self-harm rates (144) and one RCT reported significantly higher rates of self-harm in the DBT group compared to a wellness group control but did not report the significance of within-group effects (81). Bloom et al. (2012) suggested that this may be because in the intervention group reported by Springer et al. (1996) (68), in-depth discussion of self-harm was encouraged during group sessions – a practice not consistent with standard DBT (67).

Effects on suicide-related outcomes

5/11 primary studies reported on suicidality as an outcome. Two trials found significant reductions in suicidal ideation in the DBT group, but no significant difference compared to active controls (81,149). One pre-post study reported a significant reduction in suicide attempts in the DBT group compared to historical controls (84), although another found no significant change (143). Another pre-post study with no control group reported significant reductions in self-harm (suicidal or non-suicidal) following an inpatient DBT skills programme (138). No studies statistically tested differences in suicide rates.

**CBT-based approaches**

Study characteristics

Seven studies investigated CBT-based relational care approaches in inpatient settings (85–89,115,164). This included six RCTs (85–89,115), and one pre-post study without a comparison group (136). Six studies recruited adults (85–88,115,164), one did not report the age of the participants (89). In two studies, suicidal military personnel were recruited from the same military inpatient unit in overlapping time periods (86,87). The papers do not clearly state whether there is overlap in their samples of participants.

Interventions

A variety of CBT-based interventions were investigated in the included studies, which varied in content and format. They included: ‘Unified Protocol for the Transdiagnostic Treatment of Emotional Disorders’ (85), post-admission cognitive therapy (PACT) (86,87), problem-solving therapy (PST) (89), cognitive restructuring (89), cognitive behavioural suicide prevention therapy (CBSP) (115), behavioural therapy (88), and Systems Training for Emotional Predictability and Problem Solving (STEPPS) therapy (164).

Effects on self-harm

Only one study reported the effect of a CBT-based approach on self-harm: a pre-post study with no comparison groups which found a significant decrease in the number of hospital admissions for self-harm after participants engaged in STEPPS (164).

Effects on suicide-related outcomes

All seven studies examined suicide-related outcomes, most reporting no significant differences in measures of suicidal ideation (85,87,89,115), self-monitoring of suicidal intention (89), suicide attempts (86,87), ‘suicidal behaviours’ or future suicide probability (115). 2/7 studies reported a significant positive impact of CBT interventions on suicide-related outcomes. This included a RCT reporting significant reductions in suicidal threats and acts, suicidal ideation and plans in a behavioural therapy group over time and compared to the insight-oriented therapy control group (88) and a pre-post study reporting significant reductions in suicide attempts after engaging in STEPPS therapy (164). One RCT reported significantly higher suicidal ideation amongst people receiving PACT compared to enhanced usual care controls, but no significant between-group difference in the proportion of participants meeting criteria for clinically significant change in suicidal ideation (86). No studies statistically analysed differences in rates of completed suicides.

**Other psychological approaches**

Study characteristics

Eight studies investigated other psychological approaches (88,90–93,144,149,152). These included two RCTs (88,90), a non-randomised trial (149), a controlled pilot study (144), a naturalistic non-randomised controlled comparison trial (92), an uncontrolled open, case-focused trial (91), a controlled cohort study (152) and uncontrolled pre-post study (93). Two studies recruited adolescents (93,149), whilst the others recruited adults. One study specifically recruited women diagnosed with BPD (144), one recruited adults experiencing a current episode of major depression (90) and the remaining studies recruited participants presenting with suicide risk more generally.

Interventions

A variety of different psychological interventions were investigated, including: combined DBT and mentalisation-based therapy (MBT) groups (144); phone-based positive psychology (90), cognition-focused intervention (90), insight-oriented psychotherapy (88), psychodynamic-oriented crisis assessment and treatment (149), Collaborative Assessment and Management of Suicidality (CAMS) (91,92), Steps to Enhance Positivity (STEPs) therapy (93), and a brief admission crisis intervention program consisting of a 5-day individualised psychotherapy intervention (152).

Effects on self-harm

4/8 studies investigated the impact of other psychological approaches on self-harm (90,144,149,152). One RCT reported no significant difference in self-harm between participants receiving a phone-based positive psychology intervention versus a cognition-focused intervention (90). However, it did not state whether there was a significant change in self-harm over time within each group (90). A non-randomised trial found that psychodynamic-oriented crisis assessment and treatment significantly reduced self-harm, and that there was no significant difference in self-harm compared to participants receiving adapted inpatient DBT (149). A pilot study reported that self-harm rates significantly reduced for people receiving combined DBT and MBT groups, but there was no significant difference in self-harm rates compared participants receiving DBT only (144). A pre-post study reported lower rates of self-harm in a brief admission crisis intervention program group compared to treatment-as-usual controls (TAU) but did not statistically test the significance of this (152).

Effects on suicide-related outcomes

7/8 studies examined the impact of these other psychological interventions on suicide-related outcomes (88,90–93,149,152), generally indicating favourable outcomes. A cohort study found that a brief crisis intervention program was associated with lower rates of suicide attempts and significantly longer mean suicide attempt day survival compared to TAU controls (152). Two studies found CAMs was associated with significant reductions in suicidal ideation, suicidal cognition and suicide drivers, including compared to TAU matched controls (91,92). An uncontrolled pre-post study found STEPs significantly reduced suicidal ideation (93). A non-randomised trial reported that psychodynamic-oriented crisis assessment and treatment was associated with significant reductions in suicidal ideation, and that there was no significant difference compared to the adapted inpatient DBT comparison group (149).

One RCT reported significant reductions in suicidal threats and acts, suicidal ideation and suicidal plans in the group receiving insight-oriented therapy, but this was a significantly smaller reduction compared to a behavioural therapy comparison group (88). Another RCT found that phone-based positive psychology was associated with significantly higher suicidal ideation than those receiving a cognition-focused intervention (90). It did not report whether the reductions in suicidal ideation over time were statistically significant within each group (90).

**Staff training approaches**

Study characteristics

Three studies examined the effects of staff training on self-harm or suicide-related outcomes in inpatient settings (94,116,134). One was a non-randomised trial (134), one used a before-and-after trial design (116), whist the other employed a quantitative, comparative, quasi-experimental design (94). Only Bowers et al. (2008) had a control group comprising five TAU acute admission wards at the same hospitals as the intervention wards (134). Ercole-Fricke et al.’s (2016) study was conducted on an adolescent inpatient psychiatric ward (94), whereas Bowers et al. (2006) implemented city nurses on adult acute psychiatric wards (116). Bowers et al. (2008) did not specify the age of the acute admission inpatient wards studied (134).

Interventions

Two studies evaluated a ‘city nurse’ intervention, which involved employing additional nurses (one per ward) with clinical expertise to work with ward staff three days per week to move towards a low-conflict, low containment therapy-based model of nursing (116,134). Ercole-Fricke et al. (2016) evaluated ‘collaborative problem-solving training for nurses’ (94).

Effects on self-harm

All three studies evaluated the impact of the staff training interventions on self-harm. Ercole-Fricke et al. (2016) reported that ‘collaborative problem-solving training for nurses’ was associated with significant decreases in rates of self-harm incidents (94). Bowers et al. (2006) also reported a decrease in the official reporting of self-harm incidents after introducing ‘city nurses’ (116). However, Bowers et al. (2008) found no significant difference in ‘conflict events’ (including suicidal and non-suicidal self-harm, among other incidents) between intervention wards implementing ‘city nurses’ and TAU wards (134).

Effects on suicide-related outcomes

Two studies evaluated the impact of ‘city nurses’ on suicide-related outcomes (116,134). Bowers et al. (2006) reported no significant difference in suicide attempts per shift after implementing ‘city nurses’ (116). Bowers et al. (2008) found no significant difference in ‘conflict events’ (including suicidal and non-suicidal self-harm, among other incidents) between intervention wards implementing ‘city nurses’ and TAU wards (134).

**Observations**

Study characteristics

Six studies investigated the impact of special observations on self-harm or suicide-related outcomes (117–122). All had cross-sectional study designs, except one which had an uncontrolled longitudinal analysis design (117). All investigated acute adult psychiatric wards, except Bowers et al. (2011) (121) who analysed data from patients aged 17-77 from a variety of inpatient mental health ward types (including adult mental health wards, psychiatric intensive care units, forensic wards, mental health rehabilitation wards).

Interventions

4/6 studies reported on ‘constant observation’ of patients by staff (117–119,122). 3/6 studies reported on ‘intermittent observations’ (intermittent checks on patients by staff at short regular intervals) (119–121).

Effects on self-harm

5/6 studies examined self-harm as an outcome (117–120,122). They found that constant observation was either not significantly associated with self-harm (117,119) or with significantly higher rates of self-harm (118). Stewart, Bowers & Ross (2011) noted that a small number of self-harm incidents did occur during constant observation but did not statistically analyse this association (122). In two studies, intermittent observations were associated with significantly reduced self-harm (119,120).

Effects on suicide-related outcomes

2/6 studies on observations examined suicide-related outcomes (121,122). In their analysis of one year of nationally reported suicide attempts on inpatient psychiatric wards in England and Wales, Bowers et al. (2011) concluded that patients are discovered attempting suicide by staff checks (including intermittent observations) and by staff being “caringly vigilant and inquisitive”. They noted that most suicide attempts occurred in evenings or at night, peaking at times of nursing shift handovers, when supervision is at a reduced level (121). Stewart, Bowers & Ross (2011) in their cross-sectional analysis of data from 84 acute psychiatric inpatient wards found that attempted suicides during constant observation only occurred when the constant observation was initiated post-admission, not on admission. They concluded that this suggests that placing patients with a known self-harm or suicide risk under constant observations upon admission may have a 'modest protective effect' for preventing suicide attempts (122). However, they did not conduct any statistical significance testing.

**Ward and organisation-level approaches**

Study characteristics

Seven studies investigated ward- and organisation-level approaches in inpatient settings (80,123–127,156). These included one pragmatic cluster RCT (123), a controlled pre-post study (124), an uncontrolled interrupted time series study (80), and four pre-post studies with no comparison groups (125–127,156). Six were based in adult inpatient services (123–127,156) and one was based at a child and adolescent inpatient service (80).

Interventions

The studies examined a range of ward- and organisation-level approaches. Two evaluated Safewards, an explanatory model which emphasises the need to understand patients’ perspectives and view them as active agents on wards (123,156). The Safewards model incorporates ten interventions for improving relationships between patients and staff, fostering a safe ward atmosphere, and to enable staff to respond calmly to patients’ distress (123,156).

Reen et al. (2021) investigated the effect of adding twilight nursing shifts (3-11pm) and introducing a structured evening activity programme (80).

Three studies evaluated the Tidal model, a recovery model which focuses on patients’ narratives, values their expertise, promotes staff curiosity and validation, and encourages collaborative recovery with a primary goal of providing a safe environment for repair and recuperation (124–126).

One study evaluated the Bradford Refocusing model, which aims to reduce formal observations, replacing ‘control’ interventions with ‘care interventions’, directed and reviewed primarily by nursing staff (127).

Effects on self-harm

All seven studies included self-harm as an outcome, generally reporting reductions in self-harm measures. The two Safewards studies (123,156) measured ‘conflict incidents’ which included self-harm amongst other types of incidents (e.g., suicide attempts, aggression, verbal abuse, substance/alcohol use). In their pragmatic cluster RCT, Bowers et al. (2015) reported that for shifts with conflict or containment incidents, the Safewards conditions significantly reduced the rate of conflict events by 15% relative to the physical health intervention control (123). This was consistent with Dickens et al.’s (2020) uncontrolled longitudinal pre-post study reporting significant decreases in conflict rates after implementing Safewards (156).

Reen et al.’s (2020) interrupted time series study reported that although there was no significant change in the rate of self-harm on the ward, the introduction of twilight nursing shifts and a structured evening activities programme significantly reduced the proportion of adolescents self-harming on the ward, and this reduction was significantly larger in the evenings (80).

Reductions in self-harm rates were reported in pre-post studies evaluating the Tidal model (124,125), and the Bradford Refocusing model (127) but none of these studies conducted statistical significance testing. In one pre-post study, there were no self-harm incidents in the 6 months before or after implementation of the Tidal model (126).

Effects on suicide-related outcomes

4/7 studies investigated the impact of ward- and organisation-level approaches on suicide-related outcomes (123,126,127,156). As mentioned above, Bower et al.’s (2015) pragmatic cluster RCT and Dicken et al.’s (2020) pre-post study of Safewards measured ‘conflict’ incidents, which included suicide attempts amongst other incidents (e.g., self-harm, aggression, verbal aggression, substance/alcohol use). Both studies reported that Safewards was associated with significant reductions in conflict rates (123,156), including compared to a staff physical health control intervention (123). Stevenson, Barker & Fletcher (2002) reported no suicides in the 6-months before or after implementation of the Tidal model (126). Dodds & Bowles (2002) reported suicide rates before and after implementing the Bradford Refocusing model but did not conduct any statistical significance testing (127).

**Mixed interventions**

Study characteristics

Two studies evaluated ‘mixed interventions’ which involved a mix of different components (95,155). One study was a RCT (95), and the other had a quantitative descriptive study design (155). Berntsen et al.’s (2011) study was conducted at a child and adolescent inpatient unit (155). Pfeiffer et al.’s (2019) study was conducted in two adult inpatient psychiatric units (95).

Interventions

Pfeiffer et al. (2019) evaluated an intervention combining DBT strategies and peer support, aimed at improving patients’ hope and sense of belonging. Peer specialists were trained in DBT techniques for managing acute suicidal risk (such as acceptance, relaxation, mindfulness) and met with patients on the unit and for up to 12-months post-discharge to provide support (95). Berntsen et al. (2011) implemented a combination of interventions on a child and adolescent inpatient unit, including: staff training in DBT and seclusion and restraint, implementing a programme to reward patient behaviour, and introducing five patient exercise sessions per week (155).

Effects on self-harm

Self-harm was only measured in one study; Bernsten et al. (2011) reported a reduction in total self-harm incidents, and that there was an apparent increase in incident rates when the DBT program was temporarily stopped. However, they did not conduct any statistical significance testing, and noted the changes may have been attributable to a number of factors since a number of changes were introduced on the ward at once (155).

Effects on suicide-related outcomes

Pfeiffer et al. (2019) measured suicide attempt rates but were not able to statistically test changes over time due to limited power (95).

**Other approaches**

Study characteristics

Six papers examined other approaches (81,96–99,135) including two RCTs, reported in three papers (81,96,97), a correlational design with retrospective chart review (98), an uncontrolled pre-post study (99) and a pilot study without controls (135). All studies recruited adult inpatient populations.

Interventions

Three papers, from two studies, reported on a ‘caring letters’ intervention, which involved brief typed caring letters sent to patients regularly after inpatient discharge for five years (96,97) or one year (135). The letters express concern that the patient is managing okay and invite a response (96,97). The remaining studies investigated a wellness and lifestyle discussion group (81), no-suicide contracts (98), and a safety agreement tool/contract (99). No-suicide contracts and safety agreement tools are verbal or written agreements between staff and patients, indicating that the patient agrees not to kill or harm themselves and that they will seek help when experiencing thoughts or urges to harm themselves.

Effects on self-harm

Four studies investigated the impact of ‘other approaches’ on self-harm (81,98,99,135). Potter et al. (2005) reported no significant difference in self-harm rates (which included non-suicidal self-harm, attempted and completed suicides) following the introduction of safety agreement tool/contract (99). Drew et al. (2001) reported significantly higher (7x higher) rates of self-harm for people with no-suicide contracts than those without (98). The authors speculated that this may be because people who were likely to self-harm were more likely to have a no-suicide contract than those without, rather than it being an adverse effect of the contract. In Springer et al.’s (1996) RCT, significantly fewer participants in the wellness and lifestyle discussion group ‘acted out’ (displayed behaviours including self-harm, harm to others, threats of self-injury or violence, attempts to abscond, etc.) compared to the adapted inpatient DBT comparison group (81). However, it did not report the statistical significance of any within-group changes in ‘acting out behaviours’. Bennewith et al. (2014) reported that 12/80 participants receiving ‘caring letters’ presented to an ED for treatment following self-harm within 12-months post-discharge but conducted no statistical comparisons (135).

Effects on suicide-related outcomes

All five papers reported on measures of suicide-related outcomes. A RCT found that suicide rates were significantly lower for people who received ‘caring letters’ compared to no-contact controls for the first two years post-discharge (96), but not at 5-year follow-up (97). Bennewith et al. (2014) reported that 1/80 patients receiving ‘caring letters’ died by suicide within 12-months of discharge from inpatient care, but conducted no statistical significance testing (135).

Another RCT reported that suicidal ideation significantly reduced in participants attending a wellness and lifestyle discussion group, and that there was no significant difference compared to participants receiving adapted inpatient DBT (81).

Drew (2001) found that patients with no-suicide contracts were significantly more likely (5x more likely) to engage in suicidal behaviour than patients without contracts (98). The authors speculated that this is because people with high-risk of suicidal behaviour are more likely to be placed on no-suicide contracts. Potter et al. (2005) found no significant difference in self-harm rates (‘self-harm’ in this study including self-harm, suicide attempts and completed suicides) after introducing a safety agreement tool/contract (99).

**Table S1. Summary of findings from primary studies included in the reviews, by relational care approach, in inpatient mental health settings.** Green shading = evidence the approach had a significant positive impact on the outcome. Yellow shading = evidence the intervention had no significant effect on the outcome. Red shading = evidence the intervention had a significant negative effect on the outcome. Grey shading = outcome not measured, or outcome measured but no significance testing reported.

| **Study** | **Study design** | | **Intervention** | **Intervention description** | **Control group** | **Setting and participants** | **Self-harm** | **Suicide** | **Reviews study was included in** |
| --- | --- | --- | --- | --- | --- | --- | --- | --- | --- |
| **DBT-based approaches** | | | | | | | | | |
| Springer et al. (1996) (81)  Country: USA | RCT | | Adapted inpatient DBT | Daily 45-minute ‘Creative Coping’ DBT group sessions administered over 10 days by experienced nurses. | Existing wellness and lifestyle group on the ward (same number, frequency and duration of sessions) | Adults with ‘personality disorders’ admitted to a general inpatient psychiatric unit.  DBT group n = 16  Control n = 15 | The study measured “acting out behaviours” which included self-harm, harm to others, verbal threats of self-injury or violence, or attempts to ‘undermine’ their treatment (e.g., attempts at absconding). Significantly more of the DBT group participants "acted out" compared to wellness group controls (p < 0.05). The significance of any within-group changes in ‘acting out behaviours’ was not reported. | Significant reduction in suicidal ideation in both DBT and wellness group controls (p < 0.01) between baseline and discharge. No significant between-group difference (p > 0.05). | Bloom et al. (2012) Nawaz et al. (2021)  Yiu et al. (2021) |
| Gibson et al. (2014) (137)  Country: Ireland | Single-centre, non-randomised clinical trial | | Adapted inpatient DBT | Short-term inpatient DBT skills-focused group programme called ‘Living Through Distress’. Delivered as four 1-hour sessions per week over a 6-week period by experienced clinical psychologists. | TAU | Adults presenting with self-harm or BPD to an independent, not-for-profit psychiatric hospital  DBT group: n = 70  Control n = 21 | Significant decrease in self-harm post-intervention in the DBT group compared to TAU controls (p = 0.04), maintained at 3-months follow-up (p = 0.01). | Not measured. | Nawaz et al. (2021) Thibaut et al. (2019) Timberlake et al. (2020) |
| Bohus et al. (2000) (141)  Country: Germany | Prospective pilot | | Adapted inpatient DBT | Standard DBT administered over three months by trained staff. | No control | 24 female inpatients (aged 17-44) at a psychiatric hospital who had BPD and had self-harmed at least twice and/or attempted suicide at least once within the last two years | Significant reduction in self-harm frequency one-month post-DBT compared to pre-intervention (p = 0.004). | Not measured. | Bloom et al. (2012) Nawaz et al. (2021) |
| Bohus et al. (2004) (142)  Country: Germany | Non-randomised trial | | Adapted inpatient DBT | Standard DBT administered over three months by trained staff. | Wait list control (received TAU in the community) | 50 adult female inpatients meeting criteria for BPD.  DBT group: n = 31  Wait list control: n = 19 | Significant reduction in self-harm acts at post-assessment in the DBT group, compared to wait list controls (p = 0.039). | Not measured. | Bloom et al. (2012) Nawaz et al. (2021) |
| Barley et al. (1993) (82)  Country: USA | Pre-post with comparison group | | Adapted inpatient DBT | Adapted psychodynamic approach to DBT (mean length of stay was 106 days). | General adult psychiatry unit with no DBT treatment components | 130 adults and CYP (aged 16-57) with a diagnosis of BPD on a psychiatric inpatient unit implementing the adapted psychodynamic approach to DBT | Significant reduction in self-harm on the adapted DBT unit compared to the non-DBT control unit (p = 0.007). Within the adapted DBT unit, rates of self-harm during and after implementation of DBT were lower than before DBT was introduced (p < 0.05). | Not measured. | Bloom et al. (2012)  Nawaz et al. (2021) |
| Katz et al. (2004) (149)  Country: Canada | Non-randomised trial | | Adapted inpatient DBT | Two-week DBT intervention involving 10 daily, manualised training sessions and twice-weekly individual DBT therapy, and a DBT milieu. | TAU (traditional, psychodynamic-oriented crisis assessment and treatment model involving daily psychodynamic psychotherapy groups, individual psychodynamic therapy once/week and a psychodynamic-oriented milieu) | 62 adolescents (aged 14-17) with suicide attempts or suicidal ideation admitted to one of two psychiatric inpatient units (one using a DBT protocol and one TAU)  DBT group: n = 32  Control group: n = 30 | Significant reduction in self-harm (medium effect size) compared to baseline in both the DBT group (d = 0.63) and controls (d = 0.73), but no significant between-group differences. Improvements continued at follow-up but were not statistically significant. | There were no completed suicides in either group. Suicidal ideation significantly reduced in both groups post-intervention and at one-year follow-up, however there was no significant between-group difference at either time point. | Bloom et al. (2012) Griffiths et al. (2022) Nawaz et al. (2021) |
| Edel et al. (2017) (144)  Country: Germany | Pilot study | | Adapted inpatient DBT | Adapted inpatient DBT treatment lasting four weeks, involving a DBT skills-training group twice a week and two sessions a week of mindfulness group training. Patients also receive one individual session of psychotherapy per week and attend groups with occupational, sports, and (optional) music and dance therapy. | Combined DBT and mentalisation-based group therapies | 73 female inpatients diagnosed with BPD (aged 18-50) at the inpatient treatment unit for patients with BPD at a local university hospital | Self-harm rates significantly reduced in the combined DBT and MBT group at post-intervention (p = 0.008), but not in the DBT only control group (p = 0.216). However, there was no significant between-group difference (p = 0.263). | Not measured. | Timberlake et al. (2020) |
| McDonell et al. (2010) (83)  Country:USA | Pre-post study with historical control | | Adapted inpatient DBT | Adapted DBT for adolescents in a long-term inpatient facility 2000-2005. The intervention lasted 12 months and was offered at three levels of intensity (full DBT, group DBT, milieu DBT) depending upon each individual’s age and type of disorder | Historical controls 1995-1999 | Setting: Adolescent long-term inpatient facility  Intervention: n = 106, aged 12-17  Controls: n = 104; aged 12-15, of similar gender and length of stay to intervention group | Young people with three or more episodes of self-harm (28% of sample) were compared. They had significantly lower rates of self-harm across 12 months of hospitalisation, compared to historical controls (p < 0.05). | Not measured. | Bloom et al. (2012) Griffiths et al. (2022) Nawaz et al. (2021) |
| Tebbett-Mock et al. (2020) (84)  Country: USA | Pre-post study with historical controls | | Adapted inpatient DBT | Adapted DBT for adolescents, which included all aspects of the DBT model. | Historical controls receiving TAU (all patients who were hospitalised on the same unit during the same seasonal span of 8 months the year before DBT implementation). | Settings: Adolescents (aged 12-17) on an adolescent acute psychiatric inpatient unit  Intervention group: n = 425  Historical controls: n = 376 | Significant reduction in self-harm incidents in the 8-month period after implementing DBT compared to historical controls receiving TAU (p < 0.01). | Significant reduction in suicide attempts in the 8-month period after implementing DBT compared to historical controls receiving TAU (p < 0.01). | Chammas (2022) Griffiths et al. (2022) Nawaz et al. (2021) |
| Kleindienst et al. (2008) (143)  Country:Germany | Naturalistic follow-up | | Adapted inpatient DBT | Standard DBT administered over three months by trained staff as reported by Bohus et al. 2004. | No control | 31 adult female patients meeting criteria for BPD (aged 18-44) who completed inpatient DBT in Bohus trial were followed up for a further 20 months post- discharge. | Significant reductions in self-harm rates reported at 1-month follow-up after inpatient DBT (Bohus et al., 2004) persisted over an additional follow-up period of 20 months (p = 0.002). The mean number of self-harm incidents in the four-weeks prior to the follow-up were also significantly lower than in the four weeks prior to receiving the inpatient DBT intervention (p = 0.002). | There was no significant change in rates of suicide attempts over the course of the 21-month follow-up post-intervention (p = 0.18). There were no recorded completed suicides during the entire study period of two years. | Bloom et al. (2012) |
| Booth et al. (2014) (138)  Country:Ireland | Pre-post | | Adapted inpatient DBT | Six-week ‘Living through distress' skills programme based on the group skills training component of DBT. | No control | 114 adult inpatients of a psychiatric hospital who had a history of self-harm | Significant reductions in participants’ reports of self-harm in the six weeks post-DBT compared to the six weeks pre-DBT (p = 0.02) which was maintained at 3-month follow-up (p = 0.01). Note: no distinction was made between suicidal and non-suicidal self-harm. Suicide rates were not measured. | | Thibaut et al. (2019) |
| **CBT-based approaches** | | | | | | | | | |
| Bentley et al. (2017) (85)  Country:USA | Proof of concept RCT | | Unified Protocol for the Transdiagnostic Treatment of Emotional Disorders | A cognitive-behavioural intervention addressing psychological disorders with aversive reaction to frequent negative emotion. Unified protocol (5 sessions) was modified to address suicidal thoughts and behaviours in an inpatient setting. | TAU | 12 adults reporting a recent suicide attempt or active suicidal ideation in an inpatient psychiatric unit  Intervention group: n = 6  Control group: n = 6 | Not measured. | No observable between-group differences in clinician-rated suicidal thoughts and behaviour frequency at 1-month or 6-month follow-up. Unable to test the statistical significance due to small sample size. No statistically significant difference in self-reported suicidal ideation between the intervention and TAU control groups.  This study was included in Yiu et al.'s (2021) meta-analysis. This meta-analysis included 10 studies covering a range of psychosocial interventions (CBT, DBT, gratitude journalling) and concluded that overall, the psychosocial interventions did not significantly reduce suicidal ideation or suicide attempts compared to controls. | Nawaz et al. (2021) Yiu et al. (2021) |
| LaCroix et al. (2018) (86)  Country: USA | Pilot RCT | | Post-admission cognitive therapy (PACT) | An inpatient CBT programme based on the cognitive model of depression and suicide. 60-90 min CBT sessions over 3 days. The intervention is the same as described in Ghahramanlou-Holloway et al. (2020). | Enhanced usual care | 36 military personnel admitted to an inpatient psychiatric unit following a recent suicide attempt, all met criteria for acute stress disorder or PTSD. Note that participants were recruited from the same service as in Ghahramanlou-Holloway et al. (2018) with overlapping recruitment periods.  Intervention group: n = 18  Control group: n = 18 | Not measured. | There were no significant differences in suicide reattempt status at 3-month follow-up (p = 0.579). Three participants randomized to the PACT intervention and two participants randomized to the control group made at least one subsequent suicide attempt following psychiatric discharge. No participants in the intervention group, and one participant in the enhanced usual care control group, died by suicide during the follow-up period.  Suicidal ideation was higher in the PACT intervention group compared to enhanced usual care controls over time (p = 0.044), but the proportion of PACT participants who met the criteria for clinically significant change on the suicidal ideation measure was not higher than enhanced usual care control participants.  This study was included in Yiu et al.'s (2021) meta-analysis. This meta-analysis included 10 studies covering a range of psychosocial interventions (CBT, DBT, gratitude journalling) and concluded that overall, the psychosocial interventions did not significantly reduce suicidal ideation or suicide attempts compared to controls. | Navin et al. (2019) Nawaz et al. (2021) Yiu et al. (2021) |
| Ghahramanlou-Holloway et al. (2020) (87)  Country:USA | Pilot RCT | | Post-admission cognitive therapy (PACT) | Six 60-90 min sessions of individual psychotherapy over three days. Includes a focus on engagement, safety planning, CBT formulation of suicide attempt behaviour, teaching and practising CBT skills (e.g. problem-solving, emotional regulation) and relapse prevention. Same intervention as described in LaCroix et al. (2018). | Enhanced usual care = usual care + assessment services provided by the study. Usual care received by patients varied but could include individual and group therapy, art therapy, recreation therapy, medication management, visitors, and access to other healthcare. | 24 adults admitted to a military inpatient unit due to a recent suicidal crisis. Note that participants were recruited from the same service as in LaCroix et al. (2018) with overlapping recruitment periods.  Intervention group: n = 12  Control group: n = 12 | Not measured. | In three-month follow up period, there were no significant differences in suicide re-attempt status or suicidal ideation between participants in the PACT and enhanced usual care control groups. No participants died by suicide during the study period.  This study was included in Yiu et al.'s (2021) meta-analysis. This meta-analysis included 10 studies covering a range of psychosocial interventions (CBT, DBT, gratitude journalling) and concluded that overall, the psychosocial interventions did not significantly reduce suicidal ideation or suicide attempts compared to controls. | Huber et al. (2024)  Yiu et al (2021) |
| Haddock et al. (2019) (115)  Country: England | RCT | | Cognitive-behavioural suicide prevention therapy (CBSP) | Up to 20 individual CBSP sessions, each lasting 1 hour, over 6 months continuing in the community following discharge. CBSP is a psychological therapy aiming to develop a detailed understanding of an individual’s experiences of suicidality and to change their thinking processes using CBT approaches. It was carried out by clinical psychologists. | TAU | Adult patients on acute psychiatric wards from one NHS Trust who had experienced suicidal thoughts or behaviours in the 3 months prior to admission  Intervention group: n = 24  Control group: n = 27 | Not measured. | No significant differences in suicidal behaviours, suicidal ideation or future suicide probability between the CBSP intervention group and TAU control group at 6-weeks or 6-months post-baseline.  This study was included in Yiu et al.'s (2021) systematic review and meta-analysis. The results of this individual study were not reported in the review. This meta-analysis included 10 studies covering a range of psychosocial interventions (CBT, DBT, gratitude journalling) and concluded that overall, the psychosocial interventions did not significantly reduce suicidal ideation or suicide attempts compared to controls. | Yiu et al. (2021) |
| Liberman & Eckman (1981) (88)  Country: USA | RCT | | Behaviour therapy | Four hours of therapy per day over an 8-day period. The primary element was 17 hours of social skills training. It also included anxiety management training, and family negotiation and contingency contracting. Provided by an experienced psychologist assisted by two assistants. | Insight-oriented psychotherapy (four hours of therapy per day over an 8-day period, comprising individual therapy, psychodrama and group therapy and family therapy) | 24 repeat suicide attempters aged 18-47 admitted to an inpatient psychiatric unit  Intervention group: n = 12  Control group: n = 12 | Not measured | There was a significantly greater reduction in suicidal threats and acts, suicidal ideation, and suicidal plans in the behaviour therapy group compared to the insight-oriented therapy control group at 2-weeks (p < 0.05), 12-weeks (p < 0.05) and 24-weeks (p < 0.01). The behaviour therapy group showed less frequent suicidal thought at the 24-week (p < 0.05) and 36-week follow-ups (p < 0.025).  This study was included in Yiu et al.'s (2021) systematic review and meta-analysis. The results of this individual study were not reported in the review. This meta-analysis included 10 studies covering a range of psychosocial interventions (CBT, DBT, gratitude journalling) and concluded that overall, the psychosocial interventions did not significantly reduce suicidal ideation or suicide attempts compared to controls. | Yiu et al. (2021) |
| Patsiokas & Clum (1985) (89)  Country: USA | RCT | | Cognitive restructuring and problem solving | Cognitive restructuring: This intervention focused on suicide ideation and used strategies e.g. identifying cognitions, cognitive distortions and discussing the validity of basic assumptions, beliefs and attitudes related to the suicidal behaviour.  Problem-solving: This intervention focused on training patients in problem-solving skills. | Non-directive control treatment | 15 hospitalised suicide attempters in psychiatric inpatient wards at three hospitals (age not stated)  Cognitive restructuring group: n = 5  Problem-solving group: n = 5  Nondirective control: n = 5 | Not measured. | No significant differences were found between the groups in self-monitoring of suicidal intention or ideation.  This study was included in Yiu et al.'s (2021) systematic review and meta-analysis. The results of this individual study were not reported in the review. This meta-analysis included 10 studies covering a range of psychosocial interventions (CBT, DBT, gratitude journalling) and concluded that overall, the psychosocial interventions did not significantly reduce suicidal ideation or suicide attempts compared to controls. | Yiu et al. (2021) |
| Alesiani et al. (2014) (164)  Country: Italy | Pre-post | | Systems Training for Emotional Predictability and Problem Solving (STEPPS) therapy | A two-part affect programme designed to decrease recurrent suicidal behaviour. It is a manualised, cognitive-behavioural, skills-based group treatment programme, originally developed for people with BPD and people with difficulties in emotion and behaviour regulation. Delivered as a 20-week programme for patients with BPD. It combines psychoeducation and skills training with a systems component. | No control | 32 adult patients recruited from an inpatient psychiatric unit | Significant reduction in number of hospital admissions for self-harm at 12-month follow-up for people who completed the program. | Significant decrease in suicidal attempts during and after STEPPS, and at 12-month follow-up. | Nawaz et al. (2021) |
| **Other psychological approaches** | | | | | | | | | |
| Celano et al. (2017) (90)  Country: USA | RCT | | Phone-based positive psychology | Phone-based positive psychology, weekly one-on-one telephone sessions over 6 weeks. Each session focused on a different exercise including: gratitude for positive events, identifying and using personal strengths, gratitude letter, enjoyable and meaningful activities, leveraging past success and acts of kindness. | Cognition-focused control intervention | 65 adults (aged 19-84) with a current major depressive episode reporting suicidal ideation or a recent suicide attempt in inpatient psychiatric units  Positive psychology group: n = 32  Cognition-focused control group: n = 33 | No significant difference between the groups in non-suicidal self-injurious behaviour at 12-week follow-up. Significance of changes in self-injurious behaviour over time within each group was not reported. | Suicidal ideation was significantly higher in the phone-based positive psychology intervention group compared to the cognition-focused intervention control group at 6-weeks and 12-weeks. Significance of changes in suicidal ideation over time within each group was not reported. One person in the positive psychology group reported a suicide attempt, none in the cognition-focused intervention control group. | Nawaz et al. (2021) |
| Liberman & Eckman (1981) (88)  Country: USA | RCT | | Insight-oriented psychotherapy | Four hours of therapy per day over an 8-day period, comprising individual therapy, psychodrama, group therapy and family therapy. | Behaviour therapy | 24 repeat suicide attempters aged 18-47 admitted to an inpatient psychiatric unit  Intervention group: n = 12  Control group: n = 12 | Not measured | There was a significantly greater reduction in suicidal threats and acts, suicidal ideation, and suicidal plans in the behaviour therapy group compared to the insight-oriented therapy control group at 2-weeks (p < 0.05), 12-weeks (p < 0.05) and 24-weeks (p < 0.01). However, it did significantly decrease in both groups. The behaviour therapy group showed less frequent suicidal thought at the 24-week (p < 0.05) and 36-week follow-ups (p < 0.025).  This study was included in Yiu et al.'s (2021) systematic review and meta-analysis. The results of this individual study were not reported in the review. This meta-analysis included 10 studies covering a range of psychosocial interventions (CBT, DBT, gratitude journalling) and concluded that overall, the psychosocial interventions did not significantly reduce suicidal ideation or suicide attempts compared to controls. | Yiu et al. (2021) |
| Yen et al. (2019) (93)  Country: USA | Pre-post | | Steps to enhance positivity (STEPs) therapy | Modified skills to enhance positivity (STEPs) therapy. It included an in-person phase during the inpatient admission focusing on delivering psychoeducation and positive affect skills and strategies, and a remote delivery phase for four-weeks post-discharge involving weekly phone calls and daily text messages. Participants picked their own topics to cover. | No control | 20 adolescent patients (aged 12-18) at a psychiatric inpatient unit admitted due to concerns of suicide risk | Not measured. | Large effect sizes were observed for significant reductions in suicidal ideation from baseline to post-treatment (d = 1.07), and from baseline to 3-month follow-up (d = 2.97). p-values were not reported. In the year after discharge, one participant had a suicide attempt, and five were readmitted for suicidality in the following six months. | Nawaz et al. (2021) |
| Katz et al. (2004) (149)  Country: Canada | Non-randomised trial | Psychodynamic-oriented crisis assessment and treatment | | Inpatient treatment model including daily psychodynamic psychotherapy groups, individual psychodynamic therapy at least once/week, and a psychodynamic-oriented milieu | Adapted inpatient DBT | 62 adolescents (aged 14-17) with suicide attempts or suicidal ideation admitted to one of two psychiatric inpatient units (one using a DBT protocol and one TAU)  DBT group: n = 32  Control group: n = 30 | Significant reduction in self-harm (medium effect size) compared to baseline in both the DBT group (d = 0.63) and psychodynamic-oriented treatment groups (d = 0.73), but no significant between-group differences. Improvements continued at follow-up but were not statistically significant. | There were no completed suicides in either group. Suicidal ideation significantly reduced in both groups post-intervention and at one-year follow-up, however there was no significant between-group difference at either time point. | Bloom et al. (2012) Griffiths et al. (2022) Nawaz et al. (2021) |
| Edel et al. (2017) (144)  Country: Germany | Pilot study | | Combined DBT and mentalisation-based group therapies | Combined DBT and mentalisation-based group therapies.  Four weeks standard DBT procedure (as controls) plus mentalization-based treatment consisting of 2 modules of MBT per week for four weeks (the first four modules psychoeducational followed by four psychotherapeutic modules). | Four weeks standard DBT procedure involving a DBT skills-training group twice a week and two sessions a week of mindfulness group training. Patients also receive one individual session of psychotherapy per week and attend groups with occupational, sports, and (optional) music and dance therapy. | 73 female inpatients diagnosed with BPD (aged 18-50) at the inpatient treatment unit for patients with BPD at a local university hospital | Self-harm rates significantly reduced in the combined DBT and MBT group at post-intervention (p = 0.008), but not in the DBT only control group (p = 0.216). However, there was no significant between-group difference (p = 0.263). Note: this is considered to still be a significantly positive impact on self-harm given that the control group is an active control (DBT). | Not measured. | Timberlake et al. (2020) |
| Berrino et al. (2011) (152)  Country: Switzerland | Cohort study | | Brief admission crisis intervention program | A 5-day individualised psychotherapy intervention provided by experienced supervised nurses in an inpatient psychiatric crisis unit at the general hospital. It includes interpersonal intervention with the family and other close friends, especially partners, to clarify communication process and decrease conflicts, as well as teaching adaptive coping behaviours. | TAU | 200 adults hospitalised in the internal medicine unit of an emergency centre for self-intoxication.  Intervention group: n = 100  Control group: n = 100 | Lower rates of repeated self-harm at 3-month follow-up after a brief 5-day admission compared to TAU, but this was not significance tested. | Significantly lower rates of suicidal attempts (p = 0.05) and longer mean suicide attempt day survival (p = 0.05) in the brief admission intervention group compared to the TAU control group at 3-month follow-up. | Helleman et al. (2014) |
| Ellis et al. (2012) (91)  Country: USA | Pilot study of feasibility and effectiveness – open trial, case-focused design | | Collaborative Assessment and Management of Suicidality (CAMS) | CAMS is a clinical framework for maintaining a collaborative focus on reducing suicidal ideation and behaviour as a means of coping. CAMS-based problem-solving interventions may include targeting and treating hopelessness, emotional dysregulation, interpersonal isolation, impulsivity, symptoms of PTSD, or difficulties planning for the future. It is provided via two 50-minute individual therapy sessions per week, starting with completion of the Suicide Status Form. | No control | 20 adult patients hospitalised with recent histories of suicidal ideation and behaviour at a psychiatric hospital (aged 21-55, 16 female, average inpatient stay of 51 days) | Not measured. | There were significant decreases in suicidal ideation (p = 0.001), suicidal cognitions (p = 0.001), and each of the suicide drivers (pain, stress, agitation, hopelessness, self-hate, suicide probability; all p < 0.001) from the Suicide Status Form from baseline to post-intervention. | Chammas (2022) De Santis et al. (2015) Thibaut et al. (2019) |
| Ellis et al. (2015) (92)  Country: USA | Naturalistic controlled-comparison trial | | Collaborative Assessment and Management of Suicidality (CAMS) | CAMS is a structured, collaborative approach to risk assessment, treatment planning, alliance-building and risk reduction. It is a framework for treatment regardless of the therapeutic orientation placing special emphasis on developing a shared understanding of the suicidal episode, planning for safety in hospital and post-discharge, and addressing vulnerabilities such as hopelessness and self-hatred. All patients received an intensive treatment programme (general medical care, pharmacotherapy, physical activities, 2 x week individual and group psychotherapy, daily psychoeducational groups, family work, social/recreational activities). | The TAU group (matched for age and gender by propensity score) received individual therapy from a therapist who had not been CAMS trained. | Convenience sample of 52 adults admitted to a psychiatric hospital (aged 18-68;  69.2% female, 92.3% White, average length of stay 58.8 days) who reported attempted suicide or suicidal ideation within 2 months prior to admission.  Intervention group: 26  Control group: n = 26 | Not measured. | Significantly greater decreases in suicidal ideation (p < 0.05) and suicide cognitions (p < 0.05) in the CAMS group compared to TAU matched controls between admission and discharge. | Navin et al. (2019) Thibaut et al. (2019) |
| **Staff training approaches** | | | | | | | | | |
| Bowers et al. (2006) (116)  Country: England | Before-and-after trial | | City nurses | Involves employing additional nurses (one per ward) with clinical expertise to work with ward staff three days per week to move towards low-conflict, low-containment therapy-based nursing. | No control | Two acute adult admission inpatient psychiatric wards. Age not reported. | Significant decrease in self-harm incidents per shift in the 12-month period after implementing the city nurses, compared to the 3-month pre-intervention period (p = 0.004). There was a decrease in the official reporting of self-harm incidences. | No difference in suicide attempts per shift in the 12-months follow-up period after implementing the city nurses, compared to the 3-month intervention period (p = 0.9). | Cox et al. (2010) Nawaz et al. (2021) Reen et al. (2020) |
| Bowers et al. (2008) (134)  Country:England | Non-randomised controlled trial | | City nurses | Involves employing additional nurses (one per ward) with clinical expertise to work with ward staff three days per week to move towards low-conflict, low-containment therapy-based nursing. | Five TAU wards at the same hospitals | Eight acute admission wards in East London. Three were intervention wards, and five were control wards at the same hospitals. Age range not stated.  Intervention wards: 3  Control wards: 5 | There was no significant decrease in conflict events (which included both suicidal and non-suicidal self-injury) between the intervention wards implementing city nurses and the TAU control wards, controlling for patient occupancy and clustering of result by ward. | | Reen et al. (2020) |
| Ercole-Fricke et al. (2016) (94)  Country: USA | Quasi-experimental | | Collaborative problem-solving training for nurses | Training for nurses to teach them to diffuse situations for example by distracting or engaging patients in respectful conversation. | No control | An inpatient adolescent psychiatric unit | Significant reduction in self-harm incidents post-training compared to pre-training (p = 0.005). | Not measured. | Nawaz et al. (2021) Timberlake et al. (2020) |
| **Observations** | | | | | | | | | |
| Bowers et al. (2003) (118)  Country: England | Cross-sectional survey | | Constant observation | Special observation includes constant observation (the constant presence of the observer with the patient) or intermittent observation (checks at short time intervals e.g., every 15 minutes). | No control | Patients admitted for at least 2 weeks on 12 acute adult psychiatric wards in London | Self-harm was significantly associated with constant observation (p < 0.01). | No associations reported. | James et al. (2012) |
| Bowers et al. (2008) (119)  Country: England | Cross-sectional | | Special observations | Special observation includes constant observation (the constant presence of the observer with the patient) or intermittent observation (checks at short time intervals e.g., every 15 minutes). | No control | 136 acute adult mental health wards in London | Constant observation was not associated with self-harm rates, but intermittent observation was associated with significantly reduced self-harm, as were levels of qualified nursing staff and more intense programmes of patient activities. | Not measured. | James et al. (2012) |
| Bowers et al. (2011) (121)  Countries: England and Wales | Cross-sectional | | Special observations | Special observation includes constant observation (the constant presence of the observer with the patient) or intermittent observation (checks at short time intervals e.g., every 15 minutes). | No control | Data was extracted from the National Patient Safety Agency’s Reporting and Learning System regarding all attempted suicides between 1^st^ January 2009 and 31^st^ December 2009 in mental health inpatient intensive care units, secure units or wards classified as adult mental health, forensic mental health, mental health rehab or older adult mental health in England and Wales. Included patients aged 17-77 years old. | Not measured. | The analysis found that patients are discovered attempting suicide by staff checks (including intermittent observation, medication rounds, meals, routine activities) and by staff being “caringly vigilant and inquisitive” (noticing the absence of patients, their psychological distress, physical state, responding to unusual noises, etc.) It was noted that most suicide attempts occurred in evenings or at night, peaking at times of nursing shift handovers when supervision is at a reduced level. “The use of intermittent observation and other patient checks should be increased, and particularly directed to private areas of the ward. All staff should act on any sense of unease or feeling that something about a patient, their behaviour, or noises on the ward, are not right.” | Evans et al. (2022) |
| Stewart et al. (2009) (117)  Country: England | Longitudinal analysis | | Special observations | Special observation includes constant observation (the constant presence of the observer with the patient) or intermittent observation (checks at short time intervals e.g., every 15 minutes). | No control | Officially collected data covering a 2.5-year period from 16 acute adult wards at three hospitals in London. | No significant association between constant observation and self-harm rates in a longitudinal analysis of officially collected data covering a 2.5-year period. | Not measured. | Evans et al. (2022) Huber et al. (2024)  James et al. (2012) |
| Stewart & Bowers (2012) (120)  Country: England | Cross-sectional | | Special observations | Special observation includes constant observation (the constant presence of the observer with the patient, with or without engagement) or intermittent observation (checks at short time intervals e.g., every 15 minutes). | No control | Data from end-of-shift reports completed by nurses on 136 acute adult mental health wards in 67 English hospitals during 2004 and 2005 | Negative correlation between intermittent observations and incidents of self-harm. | Not measured. | Evans et al. (2022) |
| Stewart, Bowers & Ross (2012) (122)  Country: England | Cross-sectional, retrospective, case note study | | Constant observation | Constant special observation (CSO): the constant presence of a designated staff member within arm’s length or within eyesight of the patient. | No control | 84 acute adult psychiatric wards in 31 hospital locations in London and surrounding areas during 2009-2010. Data collected for the first two weeks of admission from 522 patients’ case notes. | A small number of self-harm incidents did occur during constant observation. Constant observation cannot be viewed in isolation from other forms of containment. A range of interventions are frequently used both before and during constant observation. No significance testing reported. | Attempted suicides were limited to constant observation episodes initiated post-admission only (not on admission). This provides some evidence supporting the idea that placing patients with a known self-harm or suicide risk under constant observation when they are admitted may have a modest protective effect of preventing patients attempting suicide, at least while they are under observation. No significance testing reported. | Evans et al. (2022) |
| **Ward- and organisation-level approaches** | | | | | | | | | |
| Bowers et al. (2015) (123)  Country: England | Pragmatic cluster RCT | | Safewards | Consists of an explanatory model that highlights the need to understand the patient perspective and see’s patients as active agents on wards. From the model, ten interventions are prescribed to improve relationships between patients and staff, foster a safe atmosphere on the ward and respond calmly to signs of agitation or distress. Staff are, for instance, encouraged to offer reassurance when needed, assist patients dealing with bad news and soften some of the natural power barriers. A concerted attempt is also made to highlight positive patient contributions, and capture advice and messages of support about the ward from patients who are successfully returning to the community. | Implemented a package of interventions directed at improving staff physical health | Staff and patients in 31 randomly chosen adult wards at 15 randomly chosen hospitals within 100km of central London  Safewards intervention group: n = 16 wards  Physical health intervention control group: n = 15 wards | This study measured ‘conflict’ incidents which included  e.g., self-harm, suicide attempts, aggression, substance/alcohol use, verbal abuse, etc.  For shifts with conflict or containment incidents, the Safewards condition significantly reduced the rate of conflict events by 15%, relative to the physical health intervention control. | | Mullen et al. (2022) Finch et al. (2021)  Nawaz et al. (2021) Timberlake et al. (2020) Ward-Stockham et al. (2022) |
| Dickens et al. (2020) (156)  Country: Australia | Longitudinal pre-post | | Safewards | Safewards, as described in Bowers et al. (2015). Interventions were contextualised to each ward in this study. Implementation was conducted over a 24-week period (4-week preparation, 16-week implementation, and 4-week outcome phases). The order of implementation of interventions was decided by participating wards. | No control | 8 adult inpatient mental health units in Australia | In this study, ‘conflict’ incidents were measured (which included e.g., self-harm, suicide attempts, aggression, substance/alcohol use, verbal abuse, etc.).  The mean conflict rate significantly decreased from baseline to implementation phase (p = 0.002), and from baseline to outcome phase (p = 0.001) but did not fall further from implementation phase to outcome phase. There was no significant difference in conflict-free event days across the three study phases. | | Finch et al. (2021)  Mullen et al. (2022) Ward-Stockham et al. (2022) |
| Reen et al. (2020) (80)  Country: UK | Interrupted time series | | Twilight shifts and evening activities programme | Adds a regular twilight shift (3-11pm) for nursing staff and introduces a structured evening activity programme on the ward (e.g. including drama, games, pet groups, podcasting). | No control | One child and adolescent psychiatric ward in the UK | The proportion of adolescents self-harming on the ward significantly decreased post-intervention (p = 0.001), and this reduction was significantly larger in the evenings (p = 0.004) compared to other times of day (p = 0.09). There was no significant change in the rate of self-harm per 100 bed days (p = 0.415). | Not measured. | Griffiths et al. (2022)  Nawaz et al. (2021) |
| Fletcher & Stevenson (2001) (198)  Country: England | Pre-post | | Tidal model | A recovery model which emphasises individuals’ narratives and perceptions of their own illness, moving away from a symptom-focused approach. It values patients’ expertise about their lives, promoting a curious and validating stance from staff. Collaborative contact is encouraged through regular assessments, documenting their needs and problems verbatim, and empowering them to lead their recovery. The Tidal model assumes the primary aim of acute care is to provide a safe haven within which support is provided to enable repair and recuperation work to be undertaken. | No control | One adult acute psychiatric ward in Newcastle, UK. | Rates of self-harm were 6% lower in the 6-month period after implementing the Tidal model compared to the 6-month comparison period pre-implementation. Statistical significance testing not conducted. | Not measured. | Evans et al. (2022) |
| Stevenson, Barker & Fletcher (2002) (126)  Country: England | Pre-post | | Tidal model | A recovery model which emphasises individuals’ narratives and perceptions of their own illness, moving away from a symptom-focused approach. It values patients’ expertise about their lives, promoting a curious and validating stance from staff. Collaborative contact is encouraged through regular assessments, documenting their needs and problems verbatim, and empowering them to lead their recovery. The Tidal model assumes the primary aim of acute care is to provide a safe haven within which support is provided to enable repair and recuperation work to be undertaken. | No control | Adults admitted to the inpatient psyciatric ward across a 6-month period prior to the implementation of the tidal model (n = 69), and patients (n = 81) during an equivalent period post-implementation | There were no self-harm episodes during the 6-month period pre-Tidal model implementation, or the 6-month comparison period post-implementation. | There were no episodes of suicide during the 6-month period pre-Tidal model implementation, or the 6-month comparison period post-implementation. | Identified through study cited in Evans et al. (2022) |
| Gordon et al. (2004) (124)  Country: England | Pre-post study with comparison groups | | Tidal model | A recovery model which emphasises individuals’ narratives and perceptions of their own illness, moving away from a symptom-focused approach. It values patients’ expertise about their lives, promoting a curious and validating stance from staff. Collaborative contact is encouraged through regular assessments, documenting their needs and problems verbatim, and empowering them to lead their recovery. The Tidal model assumes the primary aim of acute care is to provide a safe haven within which support is provided to enable repair and recuperation work to be undertaken. | Three adult TAU acute inpatient psychiatric wards | One adult acute inpatient psychiatric ward in Birmingham | Reported a 55% reduction in the number of intended or actual self-harm incidents in the year after implementing the Tidal model compared to the year before. Significance testing not conducted. | Not measured. | Evans et al. (2022) |
| Dodds & Bowles (2001) (127)  Country: England | Pre-post | | Bradford Refocusing Model | Model aiming to give nursing staff control over observation decisions, reduce formal observations, and replace ‘control’ interventions with ‘care’ interventions. | No control | One inner city adult male admission inpatient psychiatric ward | In the year post-implementation of the Bradford Refocusing model, there was a 67.1% reduction in self-harm incidents. Statistical significance testing not conducted. | In the 18-month period post-implementation of the Bradford Refocusing Model, there were no suicides on the ward. However, in this period, two patients died by suicide whilst away from the ward on leave, close to discharge. In the previous year, one patient died by suicide on the ward. No statistical significance testing conducted. | Cox et al. (2010) Manna (2010) Nawaz et al. (2021) Reen et al. (2020) |
| **Mixed interventions** | | | | | | | | | |
| Berntsen et al. (2011) (155)  Country: Australia | Retrospective study quantitative descriptive study | | Staff training in DBT and seclusion and restraint, programme to reward patient behaviour, five patient exercise sessions per week | Changes included: staff training in use of seclusion, restraint and DBT, a behavioural program which rewarded patients for safe and appropriate behaviour with more freedom and access to activities, and an activity program involving five structured exercise sessions as week. | No control | Psychiatric inpatient unit for children and adolescents (aged 6-16) (n = 294 children admitted during the study period) | Total self-harm incidents reduced from 60 in 2006 to 20 in 2008. Statistical significance testing was not reported. This was thought to be due to a combination of factors. An apparent increase in incident rates was seen when the DBT program was temporarily stopped. | Not measured | Griffiths et al. (2022) |
| Pfeiffer et al. (2019) (95)  Country: USA | RCT | | Peer support and DBT strategies | Intervention combining DBT strategies and peer support and aimed at improving hope and belongingness through fundamentals of peer support, for example, supportive listening and sharing of the peer supporter’s own experience. Relaxation and mindfulness techniques focused on self-acceptance were also introduced by the peer interventionist to manage acute suicidal risk. The peer specialist met with patients on the inpatient unit to establish rapport, and then continued to provide support post-discharge over the following 12 weeks. | TAU | 70 adults at high risk of suicide at two inpatient psychiatric units  Intervention group: n = 34  Control group: n = 36 | Not measured | Did not formally test for differences in suicide attempt rates across time or groups due to limited power.  However, this study was included in Yiu et al.'s (2021) meta-analysis. This meta-analysis included 10 studies covering a range of psychosocial interventions (CBT, DBT, gratitude journalling) and concluded that overall, the psychosocial interventions did not significantly reduce suicidal ideation or suicide attempts compared to controls. | Thibaut et al. (2019)  Yiu et al. (2021) |
| **Other approaches** | | | | | | | | | |
| Motto, 1976 (96); Motto & Bostrom (2001) (97)  Country: USA | RCT | | Caring letters | Brief typed caring letters sent to patients regularly after inpatient discharge for five years. The letters included an expression of concern that the person was managing alright and invited a response if they wished to send one. The letters are sent monthly for four months, then every two months for eight months, then every three months for four years. | No contact | 843 people admitted to nine psychiatric inpatient facilities in San Francisco due to a depressive or suicidal state. Mean age: 33.9 (range not provided)  Intervention group: n = 389  Control group: n = 454 | Not measured. | Suicide rates were significantly lower in the caring letters group than the no contact controls for the first 2 years (p = 0.043), but not at 5-year follow-up. | Chaudhary et al. (2020) Falcone et al. (2017)  Luxton et al. (2013) |
| Bennewith et al. (2014) (135)  Country:England | Pilot study | | Caring letters | Supportive letters sent by psychiatrists to high-risk patients over the 12-month period following discharge from inpatient psychiatric wards. | No control | 102 patients on three general adult psychiatric inpatient wards | 12/80 patients receiving the intervention on two of the wards presented to an ED for treatment following a self-harm episode in the 12-months post-discharge. 2/80 self-harmed within two weeks of discharge, a further eight within six months, and two 6-12 months after discharge. No comparisons or significance testing. | One person who had not attended the ED for self-harm, died by suicide. No comparisons or significance testing. | Chaudhary et al. (2020) |
| Springer et al. (1996) (81)  Country: USA | RCT | | Wellness and lifestyle group | The wellness and lifestyle group involving five session topics: recreation, health and fitness, families, hobbies and current events, led by a nurse. The group met for 45 minutes each weekday. | Adapted inpatient DBT (same number, frequency and duration of sessions) | Adults with ‘personality disorders’ admitted to a general inpatient psychiatric unit.  DBT group n = 16  Control n = 15 | The study measured “acting out behaviours” which included self-harm, harm to others, verbal threats of self-injury or violence, or attempts to ‘undermine’ their treatment (e.g., attempts at absconding). Significantly more of the DBT group participants "acted out" compared to wellness group controls (p < 0.05). | Significant reduction in suicidal ideation in both DBT and wellness group controls (p < 0.01) between baseline and discharge. No significant between-group difference (p > 0.05). | Bloom et al. (2012) Nawaz et al. (2021)  Yiu et al. (2021) |
| Drew (2001) (98)  Country: USA | Correlational design with retrospective chart review | | No-suicide contracts | A verbal or written agreement between staff and a patient, indicating that the patient agrees not to kill or harm themselves and that they will seek help when suicidal thoughts reach an extreme level | Inpatients discharged from a psychiatric inpatient unit that did not routinely use no-suicide contracts | 577 medical records of inpatients with a primary discharge diagnosis of a major mood disorder, schizoaffective disorder or schizophrenia, with an inpatient stay duration of five days or more, discharged from January 1996- mid-July 1997 from the adult inpatient psychiatric units of two general hospitals (one where no-suicide contracts were used routinely, and one where they were not) | Self-harm rates were significantly (seven times) higher for people with no-suicide contracts than patients without contracts. They speculated that people who were likely to self-harm were more likely to be placed on a no-suicide contract, rather than this being an adverse effect of the contract. | Patients with no-suicide contracts were significantly (five times) more likely to engage in suicidal behaviour than patients without contracts. They speculated that people who were likely to engage in suicide behaviour were more likely to be placed on a no-suicide contract, rather than this being an adverse effect of the contract. | Chammas (2022) De Santis et al. (2015) Huber et al. (2024)  James et al. (2012) |
| Potter et al. (2005) (99)  Country: USA | Pre-post | | Safety agreement tool /contract | A safety agreement tool consisting of four questions for the patient with multiple choice answers regarding thoughts of self-harm, strategies, and instructions for nursing staff. | No control | An adult acute psychiatric facility | No significant difference in the rate of self-harm (including completed suicides, attempted suicides and other self-harming behaviours) following the introduction of the safety agreement tool/contract on an inpatient ward (p = 0.49). | | Huber et al. (2024) |

A&E = Accident and Emergency; BPD = Borderline Personality Disorder; CBT = Cognitive Behaviour Therapy; CYP = Children and Young People; DBT = Dialectical Behaviour Therapy; ED = Emergency Department; LGBTIQ = Lesbian, Gay, Bisexual, Transgender, Intersex, and Queer or Questioning; NICE = National Institute for Health and Care Excellence; RCT = Randomised Controlled Trial.; TAU = Treatment As Usual.
