## Supplementary File 2 for "Quantitative evidence for relational care approaches to assessing and managing self-harm and suicide risk in inpatient mental health and emergency department settings: a scoping review"

**Supplementary File 2:** Summary of evidence for impact of relational care approaches on self-harm and suicide in emergency department settings

41 primary papers were identified evaluating the impact of 32 different relational care approaches on self-harm and/or suicide in ED settings. The characteristics and results of each primary study in ED settings are summarised narratively below, and presented in Table S2, below.

**Approaches based only in the emergency department**

**Relational approaches to risk assessments**

Study characteristics

One study investigated relational approaches to risk assessments in EDs (128); this RCT recruited adolescents (128).

Interventions

Ougrin et al.’s (2013) RCT compared therapeutic assessment with TAU (128). Therapeutic assessment is a brief, semi-structured and collaborative psychological intervention. It involves a basic psychosocial assessment and a 30-minute therapeutic intervention in the ED. It aims to help people gain insight into their problems and make changes, using elements such as joint formulation, enhancing motivation to change, identifying ways of breaking vicious cycles and creating an ‘understanding letter’, involving family members at all stages where possible (128).

Effects on self-harm and suicide-related outcomes

Ougrin et al. (2013) included both suicidal and non-suicidal self-harm in their definition of ‘self-harm’. Their RCT reported no significant difference in ED presentations with suicidal and non-suicidal self-harm, or total number of suicidal and non-suicidal self-harm episodes, between adolescents receiving therapeutic assessment versus TAU (128). There were no completed suicides in either group at 2-year follow-up (128).

**Interventions based solely in the emergency department**

Study characteristics

One study evaluated a relational care intervention based solely in the ED (100). It recruited adolescents presenting to the ED with suicidality.

Intervention

Wharff et al. (2019) compared family-based crisis intervention (FBCI) to TAU. FBCI involved a single session in the ED, delivered by trained psychiatric social workers, which focused on creating a joint crisis narrative, teaching cognitive behavioural skills, providing psychoeducation about depression, and safety planning (100).

Effects on self-harm

Wharff et al. (2019) did not evaluate FBCI’s impact on self-harm (100).

Effects on suicide-related outcomes

Patient-rated suicidality significantly reduced in both the FBCI and enhanced usual care control groups, but there was no significant difference between the groups up to 1-month follow-up. There were no completed suicides during the study period in either group (100).

**Approaches initiated in the emergency department and continued post-discharge**

**Study characteristics**

Twelve papers investigated relational care approaches initiated in the ED and continued post-ED discharge (101–107,150,158,159,165,166). These included six RCTs reported on in seven papers (103,106,107,158,159,165,166), one non-randomised trial (150), a prospective quasi-experimental study with non-random allocation of subjects (104,105), one cohort comparison study (102) and an interrupted time series study with historical controls (101). Three studies recruited adults only (101,102,159), five studies recruited adolescents only (103–107,150), and one study recruited both adults and children (165,166). The age of participants in one study was not stated (158).

**Interventions**

Nine different relational care approaches were investigated which were initiated in the ED and continued post-discharge. Most involved an initial face-to-face session with the patient, and in some cases their family, followed-up by in-person or telephone visits to check-in.

Psychoeducation/information-based ED session with follow-up

Four studies, reported on in five papers, evaluated psychoeducation/information-based ED sessions with post-discharge follow-up (101,102,158,165,166). Three papers reported on two RCTs of ‘Brief Intervention and Contact’ (BIC), which comprised a single psychoeducational session about risk, protective factors, alternative coping behaviours and referral options, followed by follow-up telephone calls or in-person visits over 18-months post-ED discharge (158,165,166).

Miller et al. (2017) evaluated ‘Safety Assessment and Follow-up Telephone Intervention’ (SAFTI) which similarly involved suicide risk screening and provision of discharge resources in the ED by a nurse, followed by post-discharge telephone calls focusing on reducing suicide risk to the patient and a significant other (101).

Stanley et al. (2018) investigated a ‘Safety Planning Intervention’ with telephone follow-up (SPI+), a brief ED-based clinical intervention combining evidence-based strategies to reduce suicidal behaviour through a prioritised list of coping skills and strategies, and at least two telephone follow-up contacts to monitor suicide risk, review and revise the safety planning intervention, and support treatment engagement (102).

CBT-based ED session with follow-up

Two studies, reported in three papers, evaluated brief CBT interventions with post-discharge follow-up (103–105). Two papers evaluated ‘Successful Negotation Acting Positively’ (SNAP) therapy, a brief, standardised cognitive behavioural treatment program for adolescent suicide attempters and their families involving psychiatric evaluation and a crisis therapy session in the ED, followed-up by six outpatient therapy sessions (104,105). Asarnow et al. (2011) evaluated ‘Family Intervention for Suicide Prevention’ (FISP), which involved a family-based CBT session designed to increase motivation for follow-up treatment and safety, delivered in the ED, and followed-up by post-discharge telephone contacts (103).

Motivational interviewing-based ED session with follow-up

Two studies evaluated motivational interviewing-based brief interventions with post-discharge follow-up (106,107). This included ‘Teen Options for Change’ which involved an adapted motivation interview being conducted with the suicidal patient, and then the family, in the ED to develop a personalised action plan, followed-up by a post-discharge letter and telephone call to facilitate implementation of the action plan (107). Grupp-Phelan et al. (2019) evaluated ‘Suicidal Teens Accessing Treatment after an Emergency Department Visit’ (STAT-ED), a brief motivational interviewing intervention for suicidal adolescents presenting to the ED which targets family engagement, problem-solving, referral assistance and involves follow-up case management telephone calls to support families with accessing mental health care (106).

Other approaches

Greenfield et al. (2002) evaluated a rapid response outpatient team, where individuals were contacted by a psychiatric nurse and psychiatrist immediately after ED-discharge to schedule a follow-up appointment and were supported by a team until longer-term care arrangements were made in the community (150). Inui-Yukawa et al. (2021) investigated assertive case management initiated in the ED and continued post-discharge	(159).

**Effects on self-harm**

Only 1/12 papers examined self-harm as an outcome. Inui-Yukawa et al. (2021) found significantly lower self-harm rates in the assertive case management group compared to enhanced usual care controls (159).

**Effects on suicide-related outcomes**

All twelve papers included suicide-related outcomes, reporting mixed results.

Psychoeducation/informational session and follow-up

Fleishmann et al.’s (2008) RCT found significantly lower suicide rates in the BIC group compared to TAU controls (165), but Bertolote et al. (2010) reported no significant difference in repeated suicide attempts in the same RCT (166). In their RCT, Amadéo et al. (2015) found no significant difference in frequency of suicidal behaviour or completed suicides between participants receiving BIC versus TAU controls at 18-month follow-up (158).

Miller et al.’s (2017) interrupted time series study reported significantly lower risk of suicide attempts, total number of suicide attempts, and scores on a ‘suicide composite’ measure (which took into account death by suicide, suicide attempts, interrupted or aborted suicide attempts and suicide preparatory acts) for participants receiving SAFTI compared to historical controls receiving TAU (101). Similarly, Stanley et al. (2018) reported that patients receiving SPI+ were significantly less likely to engage in suicidal behaviour than those receiving usual care during the 6-month follow-up period (102).

Cognitive behaviour therapy-based ED session with follow-up

No significant differences were reported in suicidal ideation between patients receiving SNAP compared to TAU in a non-randomised quasi-experimental study (104,105), or in suicidality between FISP and enhanced usual care controls in a RCT (103). Rotheram-Borus et al. (1996; 2000) reported that the suicide attempt base rates in their study were too low to be statistically analysed (104,105).

Motivational-interviewing based intervention session and follow-up

Two RCTs evaluating motivational-interviewing based interventions reported no significant difference in suicidal ideation between those in the intervention groups and enhanced usual care controls (106,107).

Other approaches

Greenfield et al.’s (2002) non-randomised trial found significantly reduced suicide-related hospitalisations at 6-months follow-up for adolescents receiving the rapid response outpatient model compared to TAU controls (150). There were no significant differences in levels of suicidality between the groups. No patients had died in either group at 6-month follow-up. Inui-Yukawa et al. (2021) reported significantly lower total numbers of suicide reattempts, and significantly lower incidence rates of recurrent suicides within 1-month, 3-months and 6-months, compared to enhanced usual care controls (159).

**Approaches starting after emergency department discharge**

Study characteristics

Twenty-seven papers, reporting on twenty-five studies, investigated relational care approaches started after ED discharge (108–114,129–133,136,139,140,145–148,151,153,154,157,160–163). 17/25 studies were RCTs (108,109,113,114,129–133,136,139,140,145,148,153,154,157,160), and there were two with controlled quasi-experimental designs (110,151), one non-randomised trial (111), one pre-post study with historical controls (146), two prospective studies without control groups (112,147), one case-control study (162,163) and one cross-sectional study (161). Twelve studies recruited adults only (113,114,130,131,136,139,145,146,148,154,160,161), four recruited adolescents only (108–111), six recruited both adults and adolescents (112,129,140,147,157,162,163), and the age range of participants was not stated in three studies (132,133,151,153).

**Interventions**

Psychological interventions

Ten studies investigated twelve different psychological interventions started after ED discharge (108,109,114,129,130,139,140,153,154,160). They varied in content and format, and included: CBT (114), skills-based CBT (108), non-directive supportive relationship treatment (108), manual-assisted CBT (MACT) (129), CBT with case management (160), interpersonal problem-solving skills training (IPSST) (140), a brief problem-oriented approach (140), brief psychodynamic interpersonal therapy (130), abandonment psychotherapy (154), attachment-based family therapy (ABFT) (109), problem-solving therapy (PST) (139) and the Attempted Suicide Short Intervention Program (ASSIP) (153).

On-demand access to crisis support

Morgan et al. (1993) investigated ‘green cards’ offered to patients presenting with self-harm for the first time. The green card provided them with access to an on-call trainee psychiatrist if needed in the future. They were encouraged to visit or phone the ED before self-harming and had the option of an on-demand crisis admission if deemed necessary (131). Two papers reported on one RCT evaluating ‘crisis cards’ offering patients 24-hour crisis telephone consultation with an on-call psychiatrists for up to six months after their first presentation to hospital for self-harm (132,133).

Follow-up contacts only

Beautrais et al. (2010) evaluated the impact of caring postcards sent regularly to patients for 12 months following discharge from an index ED attendance for self-harm (157). Six studies investigated telephone follow-up contacts after ED discharge (111,112,145,146,151,162,163). Two studies investigated telephone and letter follow-up contacts (136,147), and one study investigated a crisis card combined with telephone follow-up contacts (148). Frequency and duration of contacts varied between studies.

Other approaches

Deykin et al. (1986) investigated a ‘specialised direct service for youths’, a community-based outreach program providing support, a liaison with the hospital and advocacy with relevant agencies (110). Currier et al. (2010) evaluated a mobile crisis team which assessed patients within 48 hours of ED discharge and could refer to other forms of support (113). Shin et al. (2019) investigated case management, providing patients with four, weekly face-to-face or telephone follow-up contacts post-ED discharge and then referrals to community mental health centres (161).

**Effects on self-harm**

Eight of the papers investigated the impact of relational care approaches starting after ED discharge on self-harm (129–133,136,139,140,157), most reporting no significant impact of psychological interventions initiated post-ED discharge on self-harm.

Psychological interventions

Tyrer et al.’s (2004) RCT reported no significant difference in the likelihood of repeated episodes of self-harm between the group receiving MACT compared to TAU controls (129). Guthrie et al.’s (2001) RCT found significantly fewer patients receiving brief psychodynamic interpersonal therapy self-harmed by 6-month follow-up compared to TAU controls (130). McAuliffe et al.’s (2014) RCT found no significant difference in rates of repeat self-harm between participants receiving PST compared to TAU controls (139). McLeavey et al. (1994) did not test the statistical significance of changes in self-poisoning rates for people receiving IPSST or a brief problem-oriented approach due to its low base rate (140). Impact on self-harm was not measured in the remaining studies of psychological interventions started post-ED discharge.

On-demand access to crisis support

Morgan et al.’s (1993) RCT found no significant difference in rates of self-harm between the group receiving ‘green cards’ compared to TAU controls (131). Likewise, in another RCT there was no significant difference in self-harm repetition between a ‘crisis card’ intervention group compared to TAU controls at 6-month or 12-month follow-up (132,133).

Follow-up contacts only

A RCT reported no significant differences in the proportion of participants re-presenting with self-harm or total number of re-presentations for self-harm between a group receiving postcard follow-up contacts versus TAU, after adjusting for prior self-harm (157). Kapur et al.’s (2013) RCT did not distinguish between suicidal and non-suicidal self-harm; it reported that participants receiving letter and telephone follow-up contacts had a significantly higher total number of episodes of repeat-self harm, and higher 12-month self-harm repeat rates per individual, compared to TAU controls (136).

**Effects on suicide-related outcomes**

Twenty-two studies included suicide-related outcomes (108–114,130,131,136,139,140,145–148,151,153,154,160–163), providing some evidence that some psychological interventions and telephone follow-up contacts started post-ED discharge can improve suicide-related outcomes.

Psychological interventions

Brown et al.’s (2005) RCT reported significantly lower suicide reattempt rate and significantly fewer participants making at least one subsequent suicide attempt in the CBT group compared to enhanced usual care controls, though there was no significant between-group difference in suicidal ideation (114). Donaldson et al.’s (2005) RCT reported that both skills-based CBT treatment and non-directive supportive relationship treatment significantly reduced suicidal ideation, with no significant between-group differences in suicidal ideation or suicide attempts (108). Lin et al.’s (2020) RCT reported no significant difference in proportion of participants with a suicide attempt, or number of subsequent suicide attempts, between patients receiving CBT plus case management compared to those receiving TAU at 6- or 12-month follow-up (160).

Guthrie et al.’s (2001) RCT reported a significantly greater reduction in suicidal ideation in the brief psychodynamic interpersonal therapy group compared to TAU controls, and no completed suicides in either group (130). Andreoli et al.’s (2016) RCT reported that participants receiving abandonment psychotherapy had significantly fewer suicidal relapses and significantly improved suicidal ideation compared to intensive community TAU controls at 3-months follow-up, but no significant differences in repeat suicide attempts (154).

Diamond et al.’s (2010) RCT reported significantly greater reductions in self-reported and clinician-rated suicidal ideation, and the proportion of participants meeting criteria for clinical recovery on suicidal ideation, in adolescents receiving ABFT compared to enhanced usual care controls (109). Gysin-Maillart et al.’s (2016) RCT found no significant effect of ASSIP on suicidal ideation but did find a significant reduction in risk of attempting suicide compared to the TAU control group (Gysin-Maillart et al., 2016). This remained significant even when individuals with a diagnosis of BPD were excluded from the analysis (153).

McLeavey et al.’s (1994) RCT reported reductions in self-poisoning rates amongst people receiving IIPST compared to those receiving a problem-oriented approach, though they could not conduct statistical significance testing due to their sample size being too small (140). The paper did not specify whether the self-poisoning included suicidal and/or non-suicidal self-poisoning. McAuliffe et al. (2014) reported that in their RCT, one participant in the PST group, and two participants in the TAU group, died by suicide during the 12-month follow-up period but no statistical significance testing was conducted (139).

On-demand access to crisis support

One RCT reported that there were no suicides in either the ‘green card’ intervention group or the TAU control group during the study period (Morgan et al., 1993). Evans et al. (1999; 2005) did not measure any suicide-related outcomes (132,133).

Follow-up contacts only

4/9 studies investigating the impact of follow-up contacts on suicide-related outcomes reported significant improvements (145,146,151,162). Vaiva et al.’s (2006) RCT found significantly lower suicide reattempts in the group receiving a telephone follow-up contact one-month post-ED discharge, but not the group contacted 3-months post discharge, compared to no contact controls (145). Similarly, a quasi-experimental study reported significant reductions in the rate of repeat suicide attempts in a group of patients receiving telephone contacts over a three-month follow-up period (151). Exbrayat et al.’s (2017) pre-post study with historical controls reported significantly reduced repeat suicide attempts in the group receiving telephone follow-up contacts compared to TAU controls (146). Cebrià et al. (2013; 2015) in their case-control study reported significantly lower rates of patients who attempted suicide, and significantly longer time to next suicide attempt in the intervention group receiving telephone follow-up contacts compared to TAU controls at 1-year follow-up (162), but not 5-year follow-up (163).

1/9 studies investigating the impact of follow-up contacts on suicide-related outcomes did not distinguish between suicidal and non-suicidal self-injury (136). This RCT reported that participants receiving letter and telephone follow-up contacts had a significantly higher total number of episodes of repeat-self harm, and higher 12-month self-harm repeat rates per individual, compared to TAU controls (136).

1/9 studies reported no significant difference in number of suicide attempts or proportion of patients reattempting suicide at 12-months follow-up between patients receiving a crisis card and telephone follow-up contact intervention compared to TAU controls in an RCT (148). The remaining 3/9 studies investigating the impact of follow-up contacts on suicide-related outcomes did not conduct any statistical significance testing (111,112,147).

Other approaches

A controlled quasi-experimental study found no significant difference in repeat suicide attempts between adolescents receiving the specialised direct service for youths compared to TAU controls (110). A RCT found no significant difference in suicidal ideation at 3-months between patients in the mobile crisis team intervention group compared to TAU controls (113). A cross-sectional study found that participants who completed a post-ED discharge case management intervention were significantly more likely to have reduced suicidal risk than non-completers (161).

**Table S2. Summary of findings from primary studies included in the reviews, by relational care approach, in emergency department settings.** Green shading = evidence the approach significantly reduced the outcome. Yellow shading = evidence the intervention had no significant impact on the outcome (for controlled studies, no significant difference compared to controls). Red shading = evidence the intervention significantly increased the outcome. Grey shading = outcome not measured, or outcome measured but no significance testing reported.

| **Study** | **Study design** | **Intervention** | **Intervention description** | **Control group** | **Setting and participants** | **Self-harm** | **Suicide** | **Reviews study was included in** |
| --- | --- | --- | --- | --- | --- | --- | --- | --- |
| **Approaches based only in the emergency department** | | | | | | | | |
| *Relational approaches to risk assessments* | | | | | | | | |
| Ougrin et al. (2013) (128)  Country: England | RCT | Therapeutic assessment | A manualised procedure including a basic psychosocial assessment (approx. 1 hour) and a 30-minute therapeutic intervention which included: joint construction of a diagram involving reciprocal roles, core pain and maladaptive procedures; identifying a target problem; considering and enhancing motivation for change; exploring potential ways of breaking the vicious cycles identified; and describing the diagram and exits in an ‘understanding letter’. Family members are involved in all stages where possible. | TAU (standard psychosocial assessment) | 69 adolescents aged 12-18 presenting to EDs with self-harm referred for psychosocial assessment.  Intervention group: n = 35  Control group: n = 34 | No significant differences in ED presentations with self-harm (p = 0.53) or total number of self-harm episodes (p = 0.09) between adolescents receiving therapeutic assessment vs TAU at 24-months follow-up. There were no significant differences in frequency of self-harm in year 1 and year 2 of the follow-up (p = 0.30). There were no completed suicides in either arm at 24-month follow-up. Note: this study included both suicidal and non-suicidal self-harm as ‘self-harm’. | | NICE NG225 Evidence Review F |
| *Interventions based solely in the emergency department* | | | | | | | | |
| Wharff et al. (2019) (100)  Country: USA | RCT | Family-based crisis intervention (FBCI) + TAU | Involves a single 60–90-minute session involving creating a joint crisis narrative, cognitive behavioural skill-building, therapeutic readiness, psychoeducation about depression and safety planning. The clinical team makes recommendations for treatment with input from the patient and family. It was delivered by trained psychiatric social workers. | TAU (standard psychiatric evaluation and clinical/discharge recommendations) | 139 adolescents (aged 13-18) presenting to a paediatric ED with suicidality  Intervention group: n = 68  Control group: n = 71 | Not measured. | Suicidality significantly reduced in both the FBCI and enhanced usual care control group over time (p < 0.001), although there were no significant between group differences post-treatment or at 1-month follow-up.  There were no completed suicides reported during the study period in either condition. | Virk et al. (2022) Huber et al. (2024) |
| **Approaches initiated in the emergency department and continued post-discharge** | | | | | | | | |
| *Psychoeducation/information-based emergency department session with follow-up* | | | | | | | | |
| Fleischmann et al. (2008) (165); Bertolote et al. (2010) (166)  Countries: Brazil, India, Sri Lanka, Iran, China | RCT | Brief intervention and contact (BIC) | Single session providing psychoeducation on risk and protective factors, basic epidemiology, alternative behaviours and referral options. It is followed up by 9 phone calls or in-person visits over 18 months by someone with clinical experience. | TAU | 1867 suicide attempters aged 10-85 discharged from emergency care at collaborating hospitals in five culturally different sites (Brazil, India, Sri Lanka, Iran, China)  Intervention group: n = 922  Control group: n = 945 | Not measured. | Significantly fewer people died by suicide in the BIC group than the TAU control group (p < 0.001) during the 18-month follow-up period (165).  There were no significant differences in repeated suicide attempts between the BIC and TAU control groups during the 18-month follow-up period (p = 0.909) (166). | Chaudhary et al. (2020) Falcone et al. (2017) McCabe et al. (2018) Newton et al. (2010) Nugent et al. (2024) Luxton et al. (2013) |
| Amadéo et al. (2015) (158)  Country: French Polynesia | RCT | Brief Intervention and Contact (BIC) | Involved a one-hour information session, as close to discharge as possible and nine follow-up phone calls over 18 months by a person with clinical experience. | TAU | 515 people admitted to an ED for intentional self-harm (suicidal or non-suicidal). Age not stated.  Intervention group: n = 100  Control group: n = 100 | Not measured. | There was no significant difference in frequency of suicidal behaviour (p = 0.36) or completed suicides (p = 0.5) between patients in the BIC group compared to TAU controls at 18-month follow-up. There were also no significant differences when considering specific subgroups, including people with personality disorders (p = 0.4), a history of sexual abuse (p = 0.563) or past history of suicide attempt (p = 0.870). | Chaudhary et al. (2020) Falcone et al. (2017) |
| Miller et al. (2017) (101)  Country: USA | Interrupted time series | Safety Assessment and Follow-Up Telephone Intervention (SAFTI) | An intervention which included secondary suicide risk screening by nurses, provision of discharge resources and up to seven post-ED discharge brief telephone calls to the patient, and up to four calls to a significant other, focused on reducing suicide risk. | Historical controls receiving TAU or universal screening only | 1376 adults (aged 26-57) presenting with a suicide attempt or ideation within the week prior to a visit to one of eight EDs in the USA  Phase 1 (TAU): n = 497  Phase 2 (universal screening): n = 377  Phase 3 (universal screening + intervention): n = 502 | Not measured. | Participants in the intervention phase had a significant reduction in risk of suicide attempts compared to participants in the TAU phase (p = 0.03) and fewer total suicide attempts (p = 0.04) over a 52-week period.  There was also a significant reduction in a 'suicide composite' measure in the intervention phase compared to TAU. This composite measured five different types of suicidal behaviour, including: death by suicide, suicide attempt, interrupted or aborted attempts, and suicide preparatory acts. | McCabe et al. (2018) Nugent et al. (2024) |
| Stanley et al. (2018) (102)  Country: USA | Cohort comparison study | Safety Planning Intervention with Follow-up (SPI+) | SPI+ is a brief, structured intervention designed to mitigate future risk by providing suicidal individuals with a written, personalised safety plan to be used in the event of a suicidal crisis. It includes: i) identifying warning signs; ii) identifying internal coping strategies; iii) identifying family and friends and social places to help distract; iv) identifying individuals who can provide support during a suicidal crisis; v) listing mental health professionals and urgent care services to contact during a crisis; vi) lethal means counselling for making the environment safer. This session in the ED is followed-up at least two brief telephone calls to assess risk, review and revise the safety plan, and support treatment engagement. They continue on a weekly basis until the patient begins treatment or withdraws. | TAU  Four TAU Veterans Affairs ED comparison sites were identified, matched on  geographic location, approximate number of ED evaluations per year, and presence of an inpatient unit. Electronic health record data was obtained from them over the same time period for comparison. | 1640 adult patients presenting to nine Veterans Affairs EDs (including five Veterans Affairs ED intervention sites where SPI+ was implemented and four Veterans Affairs ED comparison TAU sites without SPI+)  Intervention group: n = 1186  Control group: n = 454 | Not measured. | Patients who visited the ED for suicidal related concerns and received SPI+ were half as likely to exhibit suicidal behaviour and compared to patients who received TAU during their ED visit (p = 0.05). This remained significant when controlling for whether the patient had a history of suicidal behaviour in the six months pre-intervention (p = 0.03). | Nugent et al. (2024) |
| *CBT-based ED session with follow-up* | | | | | | | | |
| Rotheram-Borus et al. (1996; 2000) (104,105)  Country: USA | Prospective quasi-experimental study with non-random allocation of subjects | 'Successful Negotiation Acting Positively' (SNAP) therapy | In the ED, individuals and families are provided with psychiatric evaluation, a crisis therapy session, and referral to SNAP therapy, which is then delivered over six outpatient sessions. SNAP is a brief, standardised cognitive behavioural treatment program for adolescent suicide attempters and their families. It includes a focus on role-playing, problem-solving and negotiation. | TAU | 140 adolescent female suicide attempters (aged 12-18) presenting to an ED  Intervention group: n = 65  TAU control group: n = 75 | Not measured. | The base rate of suicide attempts in both conditions was too low to statistically compare them. There were no significant differences in suicidal ideation between SNAP and control groups at 3- or 18-month follow-up. | Newton et al. (2010) Huber et al. (2024) |
| Asarnow et al. (2011) (103)  Country: USA | RCT | Family Intervention for Suicide Prevention (FISP) | A family-based cognitive behaviour therapy session in the ED designed to increase motivation for follow-up treatment and safety, followed-up by telephone contacts after discharge. | Usual ED care enhanced by provider education | 181 adolescents (aged 10-18) presenting to two EDs in Los Angeles for suicide attempts and/or ideation  Intervention group: n = 89  Control group: n = 92 | Not measured. | No significant effect of FISP on suicidality compared to enhanced usual care controls on suicidality at 2-month follow-up. | Chaudhary et al. (2020) Virk et al. (2022) |
| *Motivational interviewing-based ED session with follow-up* | | | | | | | | |
| Grupp-Phelan et al. (2019) (106)  Country: USA | RCT | Suicidal Teens Accessing Treatment After an Emergency Department Visit (STAT-ED) | Brief motivational interviewing-based intervention for suicidal adolescents presenting to EDs. It includes motivational interviewing to target family engagement, problem-solving, referral assistance, and limited case management. This case management consisted of 1-2 follow-up telephone calls within two days of ED discharge, during which the social worker talked with the parent and assisted with any problems accessing mental health treatment. | Enhanced usual care (brief mental health care consultation and referral) | 168 adolescents aged 12-17 who screened positive on the Ask Suicide Screening Questions during a non-psychiatric ED visit at two academic paediatric EDs between April 2013-July 2015  Intervention group: n = 79  Control group: n = 80 | Not measured. | No significant between-group differences in suicidal ideation in adolescents in the motivational interviewing group compared to enhanced usual care controls during the 6-month follow-up period (p = 0.72). | Virk et al. (2022) |
| King et al. (2015) (107)  Country: USA | RCT | Teen Options for Change | An adapted motivational interview is conducted with the teen individually, then with the parent/guardian to help develop a personalised action plan and provide materials, and then a follow-up letter and telephone call is provided 2-5 days after the ED visit to support and facilitate action plan implementation. | Enhanced treatment as usual.  Adolescents randomized to this group were given a crisis card with suicide emergency phone numbers in addition to written information about depression, suicide risk, firearm safety and local mental health services. | 49 adolescents (aged 14-19) presenting to an ED for a non-psychiatric emergency primary complaint, who had a positive suicide risk screen, defined as suicidal ideation, a recent suicide attempt or positive screens for both depression and alcohol or drug abuse  Intervention group: n = 27  Control group: n = 22 | Not measured. | Both the Teen Options for Change intervention and enhanced usual care control groups showed a significant reduction in suicidal ideation at 2-month follow-up (p < 0.01). However, there was no significant between-group differences. | McCabe et al. (2018) Virk et al. (2022) |
| *Other approaches* | | | | | | | | |
| Greenfield et al. (2002) (150)  Country: Canada | Non-randomised trial | Rapid response outpatient team | Involves a part-time psychiatrist and psychiatric nurse who contacts the patient immediately after assessment in the ED to schedule a follow-up appointment, and a team then provides care until long-term arrangements can be made in the community. | TAU | 286 suicidal adolescents (aged 12-17) presenting to the ED of a paediatric hospital  Intervention group: n = 158  Control group: n = 128 | Not measured. | The rapid response outpatient model significantly reduced suicide-related hospitalisations for adolescents at 6-month follow-up compared to TAU controls (p < 0.001). There were no significant between-group differences in levels of suicidality over the follow-up period, and none of the patients had died at 6 months. | Newton et al. (2010) |
| Inui-Yukawa et al. (2021) (159)  Country: Japan | RCT | Assertive case management | Assertive, continuous case management initiated at the ED and continued post-discharge. Main components included: planning regular interviews, assessment, psychoeducation, encouragement of engagement with psychiatric treatment, coordinating appointments with psychiatrists and primary care clinicians, and use of social resources. | Enhanced usual care | 592 adult (aged 20 and over) self-poisoning patients with clear suicidal intent admitted to EDs with a primary psychiatric diagnosis  Intervention group: n = 297  Control group: n = 295 | The number of non-suicidal self-harm episodes was significantly lower in the assertive case management group compared to enhanced usual care controls (p = 0.007). | Incidence of a first recurrent suicide attempt within 1-month (p = 0.02), 3-months (p = 0.0006) and 6-months (p = 0.004) was significantly lower in the assertive case management group compared to enhanced usual care controls, as was the total number of suicide attempts (p = 0.03). | Austin et al. (2024) |
| **Approaches starting after emergency department discharge** | | | | | | | | |
| *Psychological interventions* | | | | | | | | |
| Brown et al. (2005)  Country: USA | RCT | Cognitive Behaviour Therapy (CBT) | Ten outpatient sessions of CBT on a weekly or biweekly basis. The intervention was specifically developed for preventing suicide attempts. It involved identifying thoughts, images and core beliefs activated prior to the suicide attempt, learning adaptive coping strategies, addressing specific vulnerability factors, and relapse prevention. Tracking and referral services were also provided by case managers. | Enhanced usual care with tracking and referral services | 120 adults who attempted suicide and were evaluated at an ED within 48 hours of the attempt  Intervention group: n = 60  Control group: n = 60 | Not measured. | At 18-month follow-up, significantly fewer participants in the CBT group compared to enhanced TAU controls made at least one subsequent suicide attempt (p = 0.049). Participants in the CBT group also had a significantly lower suicide reattempt rate compared to enhanced TAU controls (p = 0.049). There were no significant between-group differences in suicidal ideation rates. | Nugent et al. (2024) |
| Lin et al. (2020) (160)  Country: Taiwan | RCT | Cognitive Behaviour Therapy (CBT) plus case management | Six CBT sessions delivered over four months by a case manager. It included addressing vulnerability factors, enhancing problem-solving and coping strategies, and increasing social support by addressing barriers to treatment adherence and accessing professional support. | TAU (standard case management) | 147 adult patients with suicidal behaviour admitted to an ED  Intervention group: n = 72  Control group: n = 75 | Not measured. | There was no significant between-group difference in proportion of participants with a repeat suicide attempt (p = 0.076), or number of subsequent suicide attempts (p = 0.09) at 6-month follow-up. There were also no significant between-group differences at 12-month follow-up. By 6-month follow up, two participants in the intervention group and one in the control group died by suicide (statistical significance not tested). | Nugent et al. (2024) |
| Donaldson et al. (2005) (108)  Country: USA | Pilot RCT | Skills-based treatment | A brief skills-based, cognitive behavioural intervention involving problem-solving and affect management skills. Each session included an assessment of suicidality, skill education and skill practice (including in-session and homework practice). Consists of 9 individual sessions, 1-3 family sessions, and 2 optional crisis sessions. | Non-directive, supportive relationship treatment | 39 adolescents (aged 12-17) presenting to a general paediatric ED or inpatient unit of a child psychiatric hospital after a suicide attempt  Intervention group: n = 15  Supportive relationship treatment control group: n = 16 | Not measured. | Suicidal ideation significantly reduced at 3- and 6-month follow-ups in both groups, but there were no significant differences in suicidal ideation between the skills-based treatment intervention group and supportive relationship treatment group controls. There were six suicide reattempts in the study’s follow-up period, but no significant difference in suicide reattempts between the two conditions. | Newton et al. (2010) |
| Tyrer et al. (2004) (129)  Countries: England and Scotland | RCT | Manual-assisted cognitive behaviour therapy (MACT) | Patients were sent a 70-page MACT manual and were offered up to seven face-to-face sessions with a therapist trained in MACT methods. MACT is a brief form of cognitive therapy combined with some aspects of DBT approaches. | TAU | 480 patients (aged 16-65) presenting to hospitals liked to five major centres with a self-harm episode  MACT intervention group: n = 239  TAU control: n = 241 | No significant difference in the likelihood of repeated episodes of self-harm in the MACT group compared to TAU controls at 1-year follow-up (p = 0.20). | Not measured. | Newton et al. (2022) Broadway-Horner et al. (2022) |
| McLeavey et al. (1994) (140)  Country: Ireland | RCT | Interpersonal problem-solving skills training (IPSST) | Involves five weekly sessions, starting withing two weeks of ED discharge, with training in five general stages of problem-solving, with a supplementary manual and homework assignments. | Brief problem-oriented approach | 39 intentional self-poisoning patients (aged 15-45) admitted to the ED of a regional hospital  IIPST intervention group: n = 19  Brief problem-oriented approach control group: n = 20 | Repeated self-poisoning rates were lower in the IPSST group than the control group at 1-year follow-up. However, the authors stated that the low base rate for repetition of self-poisoning meant that the sample size was too small to statistically test for differences in repeated self-poisoning.  Note: This study did not appear to distinguish between suicidal and non-suicidal self-poisoning. | | Newton et al. (2010) |
| Guthrie et al. (2001) (130)  Country: England | RCT | Brief psychodynamic interpersonal therapy | Four sessions of therapy delivered weekly by nurse therapists to people who have presented to the ED after harming themselves. The therapy focuses of identifying and helping to resolve interpersonal difficulties which contribute to distress. It is delivered to people at their home. | TAU | 119 adults who had deliberately poisoned themselves and represented to the ED of a teaching hospital  Psychodynamic interpersonal therapy group: n = 58  Control group: n = 61 | Significantly fewer patients in the psychodynamic interpersonal therapy intervention group had self-harmed compared to TAU controls at 6-month follow-up (p = 0.009). | Significantly greater reduction in suicidal ideation in the brief psychodynamic interpersonal therapy group compared to TAU controls at 6-month follow-up. There were no successful suicide attempts in either group during the follow-up period. | Broadway-Horner et al. (2022) |
| Andreoli et al. (2016) (154)  Country: Switzerland | RCT | Abandonment psychotherapy | Abandonment psychotherapy in combination with an antidepressant medication protocol and risk management program. Abandonment psychotherapy involves a manualised cognitive/psychodynamic intervention delivered over three months, with two sessions each week. It specifically targets abandonment fears and experiences, and difficulties in romantic relationships. It was delivered either by psychotherapists (AP-P) or nurses (AP-N). | Intensive community treatment as usual (psychiatric crisis intervention unit with nurse visits, medication adjustment, group therapy, social worker support and hospitalisation services) | 170 adults presenting to an ED in Switzerland with deliberate self-harm with major depressive disorder and borderline personality disorder. Self-harm had to be severe enough to require inpatient medical/surgical treatment  AP-P group: n = 70  AP-N group: n = 70  Control group: n = 30 | Not measured. | Participants receiving either form of abandonment psychotherapy had significantly fewer suicidal relapses and significantly improved suicidal ideation compared to intensive community TAU at 3-month follow-up. There were no significant differences in suicidality measures for those receiving abandonment psychotherapy delivered by psychotherapists vs nurses. There were no significant between-group differences in repeat suicide attempts at 3-month follow-up. One participant in the TAU group died by suicide. | Nugent et al. (2024) |
| Diamond et al. (2010) (109)  Country: USA | RCT | Attachment-based family therapy (ABFT) | This intervention focuses on strengthening parent-adolescent bonds through 6-8 face-to-face sessions delivered by trained therapists. They involved completion of tasks that promoted family connectedness and adolescent autonomy. It encouraged open communication about problems, including core family conflicts linked to suicide, between family members. | Enhanced usual care | 66 suicidal adolescents (aged 12-17) identified in primary care and EDs  ABFT intervention group: n = 35  Enhanced usual care control: n = 31 | Not measured. | Significantly greater rate of reduction in self-reported suicidal ideation in the ABFT group compared to enhanced usual care controls post-intervention, and benefits were maintained at 6-month follow-up. Similar between-group differences were found on clinician ratings of adolescents’ suicidal ideation.  Significantly more patients in ABFT met criteria for clinical recovery on suicidal ideation post-treatment than enhanced usual care controls, maintained at 6-month follow-up. | Virk et al. (2022) |
| Gysin-Maillart et al. (2016) (153)  Country: Switzerland | RCT | The Attempted Suicide Short Intervention Program (ASSIP) | A brief therapy based on a patient-centred model of suicidal behaviour, with an emphasis on early therapeutic alliance. It incorporates psychoeducation, a cognitive case conceptualisation and safety planning in the first three sessions, and continued long-term outreach contact via letter for 24 months.  Patients were contacted via letter for 24 months – every 3 months in the first year and every 6 months in the second year. | TAU | 120 patients who had recently attempted suicide and been admitted to the ED of a general hospital in Switzerland. Included adults but age range not stated.  Intervention group: n = 60  Control group: n = 60 | Not measured | No significant difference in suicidal ideation, but there was a significant reduction in risk of attempting suicide during the 24-month follow-up period in the ASSIP group compared to the TAU control group (p < 0.001). This remained significant when individuals with a diagnosis of borderline personality disorder were excluded from the analysis. | McCabe et al. (2018) Nugent et al. (2024) |
| McAuliffe et al. (2014) (139)  Country: Ireland | RCT | Problem-solving therapy (PST) | Group problem-solving intervention lasting 6 weeks. Consists of six 2-hour group sessions, held weekly, of structured, manualised interpersonal problem-solving skills training, facilitated by a trained therapist and co-therapist. | TAU | Patients aged 18-64 who have self-harmed during the previous three days, recruited from EDs at two sites and two acute inpatient psychiatric units  Intervention group: n = 222  Control group: n = 211 | No significant differences in rates of repeated self-harm at 6-week, 6-month or 12-month follow up between the PST intervention and TAU control groups. | One participant in the PST group, and two participants in the TAU control group died by suicide during the 12-month follow-up period. No statistical significance testing conducted. | Nawaz et al. (2021) |
| *On demand access to crisis support* | | | | | | | | |
| Morgan et al. (1993) (131)  Country: England | RCT | Green cards | Offering patients who have harmed themselves for the first time rapid, easy access to on-call trainee psychiatrists in the event of further difficulties and encouraging patients to seek help at an early stage, before self-harming, by attending the ED or by telephone. It included an on-demand crisis admission should this be judged as necessary. | TAU | 212 patients admitted to an ED after self-harming for the first time. Included adults, but age range not provided.  Intervention group: n = 101  Control group: n = 111 | There was no significant between-group difference in rates of actual self-harm between those receiving the green cards intervention versus TAU controls. | No suicides occurred in either the intervention or control group. No significance testing. | Chaudhary et al. (2020) Helleman et al. (2014) |
| Evans et al. (1999; 2005) (132,133)  Country: England | RCT | Crisis cards | Patients receive a card offering 24-hour crisis telephone consultation with an on-call psychiatrist for up to 6 months after the index presentation to hospital for self-harm. | TAU | 827 patients admitted to hospital following self-harm between Nov 1994 and July 1996. Age not stated.  Intervention group: n = 417  Control group: n = 410 | No significant difference in self-harm repetition at 6-months or 12-months follow-up between the crisis card group and controls. | Not measured. | Broadway-Horner et al. (2022) |
| *Follow-up contacts only* | | | | | | | | |
| Beautrais et al. (2010) (157)  Country: New Zealand | RCT | Postcard follow-up contacts | Caring postcards are sent regularly to patients after discharge from an index ED attendance for self-harm. Six postcards are sent over 12 months. | TAU | 327 patients aged 16+ treated for deliberate self-harm or a suicide attempt at a psychiatric emergency service at a hospital in New Zealand  Intervention group: n = 153  Control group: n = 174 | After adjustment for prior self-harm, there were no significant differences between the postcard intervention and TAU control groups in the proportion of participants re-presenting with self-harm or in the total number of re-presentations for self-harm. | Not measured. | Chaudhary et al. (2020) Falcone et al. (2017) Luxton et al. (2013) |
| Vaiva et al. (2006) (145)  Country: France | RCT | Telephone follow-up contacts | Participants were randomised to a group that received telephone follow-up contact at either 1 month post-ED discharge or 3 months post-ED discharge. | TAU (no telephone follow-up contacts) | 605 adults (aged 18-65) discharged from 13 EDs in France after attempted suicide by deliberate self-poisoning  1-month telephone follow-up group: n = 147  3-month telephone contact group: n = 146  No telephone contact TAU: n = 312 | Not measured. | Suicide re-attempts were significantly lower in the one-month telephone follow-up group compared to TAU controls (n = 0.03) over the first six months after follow-up contact.  For participants contacted at three months post-ED discharge, the number who reattempted suicide was not significantly lower than controls (p = 0.27). | Chaudhary et al. (2020) Falcone et al. (2017) Luxton et al. (2013) Nugent et al. (2024) |
| Termansen & Bywater (1975) (151)  Country: Canada | Quasi-experimental four group cohort study (follow-up contact groups 1 and 2, assessment only groups 3 and 4) | Telephone follow-up contacts | Groups 1 and 2 received initial assessment in the emergency ward, follow-up contacts for 3 months, and re-assessment at 3 months. The follow-up contacts involved a minimum of daily contact for week 1 post-discharge, every two days for week 2, twice a week for weeks 3 and 4, once a week for weeks 5-8, and every two weeks for weeks 9-12. | Groups 3 and 4 had no follow-up contacts.  Group 3 received initial assessment in the emergency ward, no follow-ups but assessment at 3-months.  Group 4 was identified by admission records only, did not receive follow-ups and was assessed at 3-months. | 202 suicide attempters discharged from emergency care. Age not stated.  Group 1: n = 57  Group 2: n = 57  Group 3: n = 50  Group 4: n = 38 | Not measured. | There was a significantly lower rate of repeat suicide attempts in group 1 (who received initial assessment + follow-up contacts) over a 3-month follow-up period (p < 0.05). Significance of changes in suicide reattempt rates in the other groups not reported. | Luxton et al. (2013) |
| Donaldson et al. (1997) (111)  Country: USA | Non-randomised trial | Telephone follow-up contacts | Three phone interviews were scheduled with patients and parents at 1-, 2- and 6-weeks post-ED discharge. These focused on treatment expectations, outpatient services, problem, concerns, and resistance to attending outpatient psychotherapy sessions. | TAU | 101 adolescents (aged 12-18 years) presenting to an ED following a suicide attempt  Intervention group: n = 23  Control group: n = 78 | Not measured. | None of the participants in the telephone follow-up contact intervention group attempted suicide at 3-month follow-up, whereas 9% in the TAU control group did. No significance testing. |  |
| Exbrayat et al. (2017) (146)  Country: France | Pre-post study with control | Telephone follow-up contacts | Telephone follow-up contacts were made to patients by a nurse at 8-, 30- and 60-days after treatment for attempted suicide. They assessed their suicide risk and treatment adherence during these calls. | Historical controls receiving TAU | 823 adult patients admitted to an ED for suicide attempt between 1^st^ Jan-31^st^ Dec 2010, who had no history of psychiatric hospitalisation exceeding 72 hours  Intervention group: n = 436  Control group: n = 387 | Not measured. | Repeat suicide attempts were significantly lower in the intervention group receiving telephone follow-up contacts compared to historical controls receiving TAU (p = 0.037) at one-year follow-up. | Chaudhary et al. (2020) Nugent et al. (2024) |
| Normand et al. (2018) (147)  Country: France | Cohort study | Telephone and letter follow-up contacts | Telephone follow-up calls post-ED discharge, followed by a standardised letter in cases where telephone contact attempts were unsuccessful. Group A was contacted at 1 week and 1, 6- and 12-months post-discharge. | Group B were contacted at 1 week, 1 month and 6 months. | 173 people aged 16-21 years old admitted to an ED for suicide attempt  Group A: n = 93  Group B: n = 80 | Not measured. | At one-year follow-up, 23/93 successfully contacted young people had reattempted suicide at least once. No statistical significance testing. | Chaudhary et al. (2020) |
| Cebrià et al. (2013; 2015) (162,163)  Country: Spain | Case-control | Telephone follow-up contacts | Patients received a systematic, one-year telephone follow-up programme. They were contacted by telephone at 1 week and 1-, 3-, 6-, 9-, and 12-months post-ED discharge. The phone calls involved assessing risk, treatment adherence, and intervention for crisis situations. When increased suicide risk was identified, an urgent ED visit was arranged. Patients under 18 years old received thorough assessment by a clinical psychologist, specific psychotherapeutic intervention with the family, and 12-month telephone follow-up. | TAU | 991 patients of all ages discharged from the EDs of two hospitals after a suicide attempt  Intervention group: n = 604  Control group: n = 387 | Not measured. | Time to next suicide attempt was significantly longer in the intervention group compared to usual care controls at 1-year follow-up (p < 0.0005) but not at 5-year follow-up (p = 0.294). Rate of patients who attempted suicide was also significantly lower in the intervention group at 1-year follow-up (p = 0.005) but not 5-year follow-up (p = 0.401) | Chaudhary et al. (2020) Falcone et al. (2017) Nugent et al. (2024) |
| Catanach et al. (2019) (112)  Country: USA | Prospective pilot | Telephone follow-up contacts | Minimum of five weekly telephone follow-up contacts post-ED discharge. The goal of each call was to: assess risk, review and revise the ED discharge plan, identify obstacles to treatment, identify additional needed resources, and provide continued caring contact until the patient attended appropriate outpatient appointments. If indicated, safety planning was done during the calls. | No control | 2737 visits to the ED by patients (of any age) evaluated for suicidal behaviour in, and discharged home from, 15 EDs across Colorado | Not measured. | When asked, only 7/1924 participants reported a suicide attempt during their engagement with the telephone follow-up intervention. However, 93 patients were referred more than once. No statistical significance testing conducted. | Nugent et al. (2024) |
| Kapur et al. (2013) (136)  Country: England | Pilot RCT | Letter and telephone follow-up contacts | Patients were provided with a leaflet listing local and national sources of help by post. They were given two telephone calls within the first two-weeks post-ED contact, and then a series of letters over a 12-month period. The purpose of the calls was to make contact and facilitate access to support. The letters included a general statement of concern, modified where appropriate to tailor them to individuals’ circumstances. | TAU | 66 adults presenting to two EDs with self-harm (regardless of suicidal intent) between November 2010 – May 2011 | The total number of episodes of repeat self-harm (p = 0.016), and 12-month repeat rate per individual (p = 0.046) was significantly higher in the intervention group compared to usual care controls. Note: this study did not distinguish between suicidal and non-suicidal self-harm. | | Chaudhary et al. (2020) Falcone et al. (2017) |
| Mouaffak et al. (2015) (148)  Country: France | RCT | Crisis card and telephone follow-up contacts | Patients were given a card providing the telephone number of a senior psychiatrist available 24/7 at the ED. Patients were called 2-weeks, 1-month and 3-month post-discharge by a trained nurse, psychologist or psychiatrist. Calls involved: brief assessment, evaluation of adherence to treatment, and discussing the patient’s current situation and any changes. When the risk for suicide was detected, an urgent ED visit was arranged. | TAU | 320 adults admitted to an ED for a suicide attempt  Intervention group: n = 160  Control group: n = 160 | Not measured. | No significant between-group difference in proportion of patients reattempting suicide (p = 0.98), or number of suicide attempts, at 12-months (p = 0.98). Response to the intervention did not differ according to past history of suicide attempts. | Chaudhary et al. (2020)  Falcone et al. (2017) |
| *Other approaches* | | | | | | | | |
| Deykin et al. (1986) (110)  Country: USA | Quasi-experimental | Specialised direct service for youths | A specialised direct service which is a community-based outreach program providing support, a liaison with the hospital, and advocacy with relevant agencies. Number of contacts was unreported. | TAU | 319 suicidal adolescents (aged 13-17) presenting to EDs at two hospital  Intervention group: n = 172  Control group: n = 147 | Not measured. | There was no significant difference in repeat suicide attempts between the specialist direct service intervention group and TAU controls. | Newton et al. (2010) |
| Currier et al. (2010) (113)  Country:USA | RCT | Mobile crisis team | Community-based clinical assessment conducted by a mobile crisis team within 48 hours of discharge at the patient’s choice of location. Clinicians could refer to other forms of support as needed. | TAU | 120 adult patients presenting voluntarily or brought by police to the ED with suicidal thoughts, plans or behaviours  Intervention group: n = 56  Control group: n = 64 | Not measured. | There was no significant difference in suicidal ideation over the 3-month follow-up between the mobile crisis team intervention group and TAU controls (p = 0.7416). | Chaudhary et al. (2020) |
| Shin et al. (2019) (161)  Country: South Korea | Cross-sectional | Case management | Case management linking patients to psychiatric services and rehabilitation centres. Includes four, weekly follow-up face-to-face or telephone sessions. Case managers include social workers, nurses and clinical counsellors. Following these follow-up sessions, patients are referred to community mental health centres. | No control | 439 suicide attempters visiting an ED in South Korea between October 2013 – December 2017. Age range not stated but included adults.  Case management completers: n = 277  Case management non-completers: n = 162 | Not measured. | Compared with the incomplete group, the group of patients who completed case management were significantly more likely to have reduced suicide risk. | Nugent et al. (2024) |

A&E = Accident and Emergency; BPD = Borderline Personality Disorder; CBT = Cognitive Behaviour Therapy; CYP = Children and Young People; DBT = Dialectical Behaviour Therapy; ED = Emergency Department; LGBTIQ = Lesbian, Gay, Bisexual, Transgender, Intersex, and Queer or Questioning; NICE = National Institute for Health and Care Excellence; RCT = Randomised Controlled Trial.; TAU = Treatment As Usual.
